# Nanopore sequencing of 1000 Genomes Project samples to build a comprehensive catalog of human genetic variation

**DOI:** 10.1101/2024.03.05.24303792

**Authors:** Jonas A. Gustafson, Sophia B. Gibson, Nikhita Damaraju, Miranda PG Zalusky, Kendra Hoekzema, David Twesigomwe, Lei Yang, Anthony A. Snead, Phillip A. Richmond, Wouter De Coster, Nathan D. Olson, Andrea Guarracino, Qiuhui Li, Angela L. Miller, Joy Goffena, Zachery Anderson, Sophie HR Storz, Sydney A. Ward, Maisha Sinha, Claudia Gonzaga-Jauregui, Wayne E. Clarke, Anna O. Basile, André Corvelo, Catherine Reeves, Adrienne Helland, Rajeeva Lochan Musunuri, Mahler Revsine, Karynne E. Patterson, Cate R. Paschal, Christina Zakarian, Sara Goodwin, Tanner D. Jensen, Esther Robb, The 1000 Genomes ONT Sequencing Consortium, University of Washington Center for Rare Disease Research (UW-CRDR), Genomics Research to Elucidate the Genetics of Rare Diseases (GREGoR) Consortium, W. Richard McCombie, Fritz J. Sedlazeck, Justin M. Zook, Stephen B. Montgomery, Erik Garrison, Mikhail Kolmogorov, Michael C. Schatz, Richard N. McLaughlin, Harriet Dashnow, Michael C. Zody, Matt Loose, Miten Jain, Evan E. Eichler, Danny E. Miller

## Abstract

Less than half of individuals with a suspected Mendelian condition receive a precise molecular diagnosis after comprehensive clinical genetic testing. Improvements in data quality and costs have heightened interest in using long-read sequencing (LRS) to streamline clinical genomic testing, but the absence of control datasets for variant filtering and prioritization has made tertiary analysis of LRS data challenging. To address this, the 1000 Genomes Project ONT Sequencing Consortium aims to generate LRS data from at least 800 of the 1000 Genomes Project samples. Our goal is to use LRS to identify a broader spectrum of variation so we may improve our understanding of normal patterns of human variation. Here, we present data from analysis of the first 100 samples, representing all 5 superpopulations and 19 subpopulations. These samples, sequenced to an average depth of coverage of 37x and sequence read N50 of 54 kbp, have high concordance with previous studies for identifying single nucleotide and indel variants outside of homopolymer regions. Using multiple structural variant (SV) callers, we identify an average of 24,543 high-confidence SVs per genome, including shared and private SVs likely to disrupt gene function as well as pathogenic expansions within disease-associated repeats that were not detected using short reads. Evaluation of methylation signatures revealed expected patterns at known imprinted loci, samples with skewed X-inactivation patterns, and novel differentially methylated regions. All raw sequencing data, processed data, and summary statistics are publicly available, providing a valuable resource for the clinical genetics community to discover pathogenic SVs.

## INTRODUCTION

As an initiative to sequence a large set of healthy reference genomes from globally diverse ancestries, the 1000 Genomes Project (1KGP) marked a significant milestone in genomic research, yielding the first sequencing-based map of normal patterns of human genetic variation that could be used to filter and prioritize candidate disease-causing variants (International HapMap Consortium 2005; Byrska-Bishop et al. 2022; 1000 Genomes Project Consortium et al. 2015). The impact of 1KGP on our understanding of human genetic diversity has been enormous, and the flagship papers have been cited more than 10,000 times in clinical and basic research studies. In addition to the profound insights presented in the 1KGP papers, the success of the project has been amplified by the use of high-quality, open-access, and diverse datasets. Today, databases such as gnomAD (Koenig et al. 2023) and DECIPHER (Firth et al. 2009) have built on the 1KGP principles for determining the population allele frequency of variants, which can aid in variant interpretation. While these databases have benefited from pooling data from large projects, such datasets are not readily available for long-read sequencing (LRS) data, which are better able to resolve structural variants (SVs) and identify variants in complex regions of the genome (Ebert et al. 2021; Liao et al. 2023; Chaisson et al. 2019).

Structural variants, defined as insertions, deletions, duplications, inversions, repeat expansions, and translocations at least 50 bp in size, are major contributors to genetic diversity and disease susceptibility and are more likely to have a larger effect size than single nucleotide variants (SNVs) (Eichler 2019). SV calling using short-read sequencing approaches can be challenging because they detect fewer than half of the ∼25,000 SVs present in an individual, are incapable of fully resolving the complex structure of many SVs, and have low concordance between callers (Chaisson et al. 2019; Zhao et al. 2021; Cameron et al. 2019). These challenges extend into clinical testing where commonly used approaches, such as exome sequencing, have low sensitivity for SV detection, meaning individuals with disease-causing SVs may remain undiagnosed even after comprehensive clinical genetic testing (Hiatt et al. 2021; Cohen et al. 2022; AlAbdi et al. 2023; Miller et al. 2021). Therefore, there is broad interest in using new technologies like LRS to develop comprehensive catalogs of common human SVs to facilitate improved detection of disease-associated variants (Wojcik et al. 2023).

There are now numerous examples of LRS-based methods being used to identify and comprehensively resolve SVs missed by prior clinical testing methods, leading to increased interest in the use of LRS as a single data source in the clinical environment. Historical concerns about cost, error rates, and computational tools available for both commercially available LRS technologies (Pacific Biosciences, PacBio and Oxford Nanopore Technologies, ONT) have also largely been resolved (Logsdon et al. 2020; Wang et al. 2021; Kolmogorov et al. 2023). The combination of falling costs, improving error rates, simplified sample preparation requirements, and standardization of bioinformatics tools has created a clear path toward the clinical use of LRS as a single test in the near future (Wojcik et al. 2023; Damaraju et al. 2024).

While analyses of 1KGP to date have made profound contributions using arrays or short-read sequencing technology, these approaches are inherently limited in their ability to capture certain types of genetic differences, such as SVs, repeat expansions, and epigenetic markers like methylation status. Building on the landmark effort of the 1KGP, the 1000 Genomes Project ONT Sequencing Consortium (1KGP-ONT) is leveraging ONT LRS with the goal of generating high-coverage, high-quality sequencing data for over 800 samples from the 1KGP sample set. This international effort aims to: 1) assess both assembly-based and alignment-based approaches to LRS data analysis; 2) evaluate variants in difficult-to-analyze regions of the genome; and 3) facilitate the identification of SVs that are uncharacterized or difficult to detect by short-read approaches. This effort is complementary to work from other groups performing LRS of 1KGP samples, such as the Human Pangenome Reference Consortium (HPRC) (Wang et al. 2022), Human Genome Structural Variant Consortium (HGSVC) (Ebert et al. 2021), and the recent release of low-coverage ONT sequencing data of nearly 900 1KGP samples (Noyvert et al. 2023), and it demonstrates the increasing likelihood that the entire collection will ultimately be sequenced using both LRS sequencing platforms commonly used today. Following 1KGP principles, all data generated through the 1KGP-ONT consortium are publicly released for immediate incorporation into clinical and basic research efforts.

Here, we present results from analysis of the first 100 samples sequenced by the 1KGP-ONT consortium. Using these data, we describe variation that would be difficult or impossible to detect or fully resolve using short-read technology, including repeat expansions associated with disease in *RFC1* and *ATXN10*, skewed patterns of X-chromosome inactivation in 46,XX samples, and differentially methylated regions (DMRs) unique to single samples. Because the major goal of this effort is to develop a catalog of common human SVs for filtering and prioritizing disease-associated SVs, we demonstrate how SVs from a modest number of individuals can be used to filter variants in unsolved cases and identify high-priority regions for follow-up analysis.

## RESULTS

Approximately 3,200 cell lines or DNA samples from the 1KGP are available at the National Human Genome Research Institute (NHGRI) Sample Repository for Human Genetic Research housed at the Coriell Institute for Medical Research repositories (Coriell) (International HapMap Consortium 2005; 1000 Genomes Project Consortium et al. 2015). These anonymized samples, which are not associated with medical or phenotypic data, are from individuals who self-reported ancestry, sex, and good health at the time of sample collection. We selected 100 samples from all 5 superpopulations based on their absence from other large-scale sequencing efforts (Liao et al. 2023; Ebert et al. 2021; Noyvert et al. 2023); we did not attempt to balance subpopulations within these samples, and four of the first 100 samples represent two parent–child pairs **(Figure 1A, Table S1)**.

**Figure 1.**
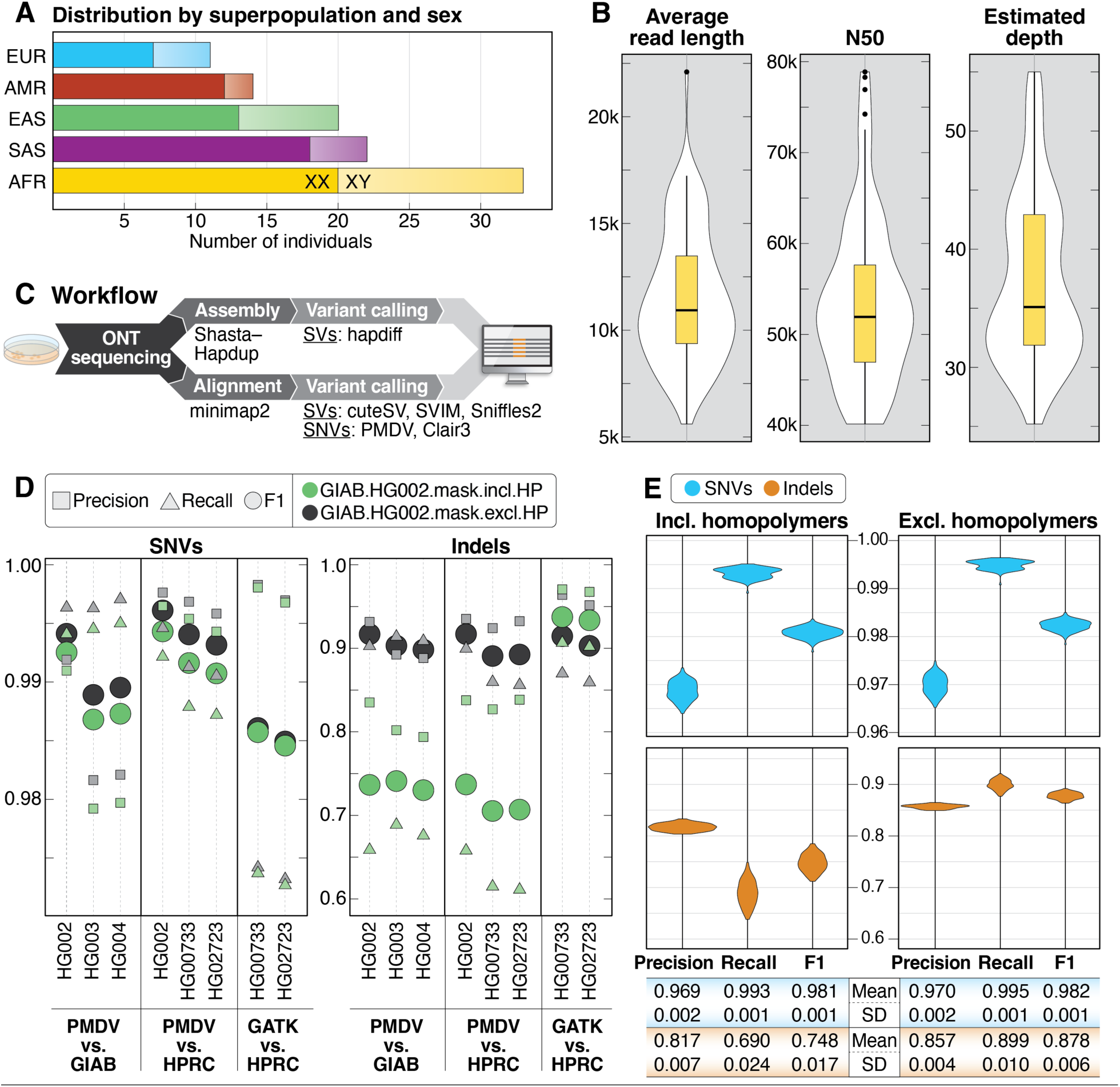
Summary statistics of samples, sequencing and small variant detection. **A:** Samples selected for sequencing are shown by superpopulation and sex. **B:** Violin plots showing average read length, read N50, and average depth of coverage for all 100 samples. **C:** DNA was extracted from cells grown from aliquots received from Coriell and sequenced using the R9.4.1 pore. Data was analyzed using both alignment– and assembly-based approaches. **D:** Comparison of precision, recall, and F1 scores for SNVs and indels called from ONT data (PMDV) or Illumina data (GATK) compared to GIAB or HPRC calls for 5 high-confidence samples genome-wide in GIAB high-confidence regions only (GIAB.HG002.mask.incl.HP) and when excluding homopolymers in the GIAB high-confidence regions (GIAB.HG002.mask.excl.HP). Homopolymers were defined as any sequence of four identical nucleotides or more, including one bp flanking each side of the sequence. **E:** Precision, recall, and F1 scores for SNVs and indels from chromosomes 1–22 called with PMDV in GIAB high-confidence regions (including homopolymers) and GIAB high-confidence regions when excluding homopolymers.

### Sequencing pipeline

To create a more comprehensive and accurate catalog of SVs and variants within difficult-to-sequence or repetitive regions of the genome, we isolated high molecular weight (HMW) DNA using an optimized protocol from lymphoblastoid cell lines (LCLs) that were obtained from Coriell and cultured in the lab. After sequencing using the R9.4.1 pore and base calling, we obtained an average depth of coverage of 37.4x and read N50 of 53.8 kbp for these 100 samples **(Figure 1B, Table S2)**.

All samples were processed using two separate pipelines **(Figure 1C)**. First, an in-house alignment-only pipeline was developed using minimap2 for alignment, Clair3 for small variant calling, and Sniffles2, CuteSV, and SVIM for SV calling (Heller and Vingron 2019; Jiang et al. 2020; Li 2018; Zheng et al. 2022; Smolka et al. 2024). Single nucleotide variant calls from this pipeline were used to ensure sample integrity by comparison with short-read-based variant calls from previous studies (Byrska-Bishop et al. 2022). Second, samples were processed using the Napu pipeline, which generates assembly-based SV calls using Hapdiff after generating a phased *de novo* assembly using Shasta-Hapdup, minimap2 alignment-based small variant calls using Pepper-Margin-DeepVariant (PMDV), and minimap2 alignment-based SV calls using Sniffles2 (Kolmogorov et al. 2023; Shafin et al. 2021; Smolka et al. 2024). Outputs from both pipelines have been made publicly available.

### Small variant accuracy

We evaluated the performance of our variant-calling pipelines by comparing small variant calls (SNVs and indels <50 bp) to those generated by prior studies and using orthogonal short– and long-read sequencing technologies. To do this, original ONT sequencing data was obtained for five samples sequenced using the R9.4.1 pore that were also previously sequenced on the Illumina and PacBio platforms (HG002/NA24385, HG003/NA24149, HG004/NA24143, HG00733, and HG02723) (Shafin et al. 2020). The ONT data for these five samples was downsampled to similar depth of coverage and representative read N50 values as the 100 samples reported here **(Table S3)**. Each sample was then processed using the in-house minimap2 (Clair3) and Napu (PMDV) pipelines.

Using published SNV and indel calls from Genome in a Bottle (GIAB; HG002, HG003, and HG004) and the HPRC(HG002, HG00733, and HG02723) as benchmark sets, we calculated precision, recall, and F1 scores: 1) genome-wide; 2) only for GIAB HG002 v4.2.1 high-confidence regions (Wagner et al. 2022); and 3) for GIAB HG002 v4.2.1 high-confidence regions excluding homopolymers (defined as 4 or more consecutive identical nucleotides +/–1 bp on each side) **(Figure 1D, Figure S1)**. The same statistics were calculated for the published small variant calls from Illumina short-read data for two samples (HG00733 and HG02723).

Similar to prior studies, we observed slightly higher precision, recall and F1 scores for SNVs called from ONT data within GIAB high-confidence regions both including and excluding homopolymers; we observed lower scores for indels when comparing ONT to Illumina in GIAB high-confidence regions only, which markedly improved when homopolymer regions were excluded (Kolmogorov et al. 2023; Harvey et al. 2023).

We performed similar recall, precision, and F1 calculations using small variant calls from chromosomes 1–22 for the 100 samples presented here **(Figure 1E, Figure S2)**. Because high-quality “truth set” assemblies do not exist for these 100 samples, we used the previously published Illumina short-read data as a truth set. Within HG002 high-confidence regions as above, the average SNV precision and recall were 0.969 and 0.993, respectively, while indel precision and recall were 0.817 and 0.690 **(Figure 1E)**. When homopolymers were excluded, SNV precision and recall improved to 0.970 and 0.995. While indel precision improved slightly to 0.857, homopolymer masking had a more substantial effect on indel recall, which increased to 0.899. Overall, these results validated that both of our variant-calling approaches (Clair3 and PMDV) were capable of providing high-quality small variant calls that were concordant with prior studies (Kolmogorov et al. 2023).

### Genome assembly

We performed *de novo* genome assemblies for each of the 100 samples using both the Napu pipeline (which runs Shasta-Hapdup) and Flye (Kolmogorov et al. 2023; Shafin et al. 2020). In general, we found that Flye assemblies had a higher contig NG50 than Shasta-Hapdup assemblies **(Figure 2A),** and results were robust to read N50 differences **(Figure 2B)**. We saw similar contig NG50 patterns when our analysis included the 5 benchmarking genomes with similar average depth of coverage and read N50. The assembled genomes were highly complete, with each assembly covering approximately 93.5% (Flye) and 93.6% (Shasta-Hapdup) of the GRCh38 reference genome **(Figure S3)** and a consensus accuracy similar to previously published studies using the R9 pore **(Figure 2C)** (Kolmogorov et al. 2023).

**Figure 2.**
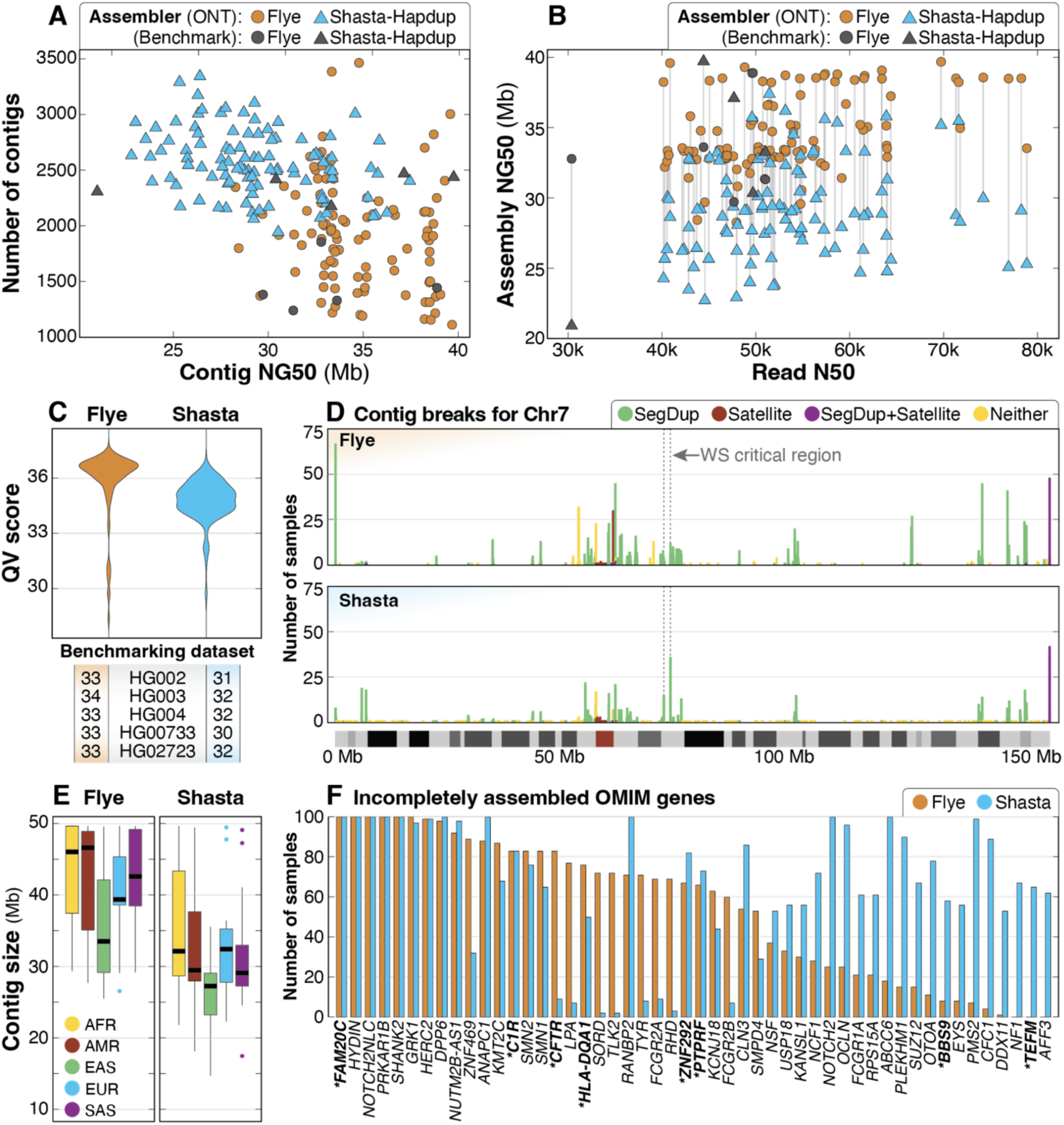
Summary of *de novo* assembly results. **A:** Contig NG50 compared to total number of contigs for both assembly methods shows that the haploid assemblies generated by Flye are longer and have fewer contigs than Shasta-Hapdup, but no contigs generated by Flye exceed 40 Mbp. Assemblies for each benchmarking sample show similar statistics. **B:** Read N50 compared to assembly NG50 shows that assembly NG50 does not significantly improve with higher read N50. **C**: QV scores for both Flye and Shasta-Hapdup assemblies show slightly higher assembly QV scores for the haploid Flye assemblies. Values for the five benchmarking genomes are shown. **D**: Count of contig breaks for all 100 samples on chromosome 7 demonstrate that assembly breaks cluster in similar locations when using both assembly approaches and that there are a large number of single breaks spread across the chromosome. The 1.5–1.8 Mbp Williams-Beuren syndrome critical region is indicated with a dashed box and is flanked by clusters of assembly breaks within segdups (Morris 1993). The position of assembly breaks were categorized as “Satellite” (only satellite repeats), “SegDup+Satellite” (segdups and satellite repeats), “SegDup” (only segdups) or “Neither” (outside segdups and satellite repeat regions). **E**: Contig sizes filtered for contigs longer than 1 Mb for each superpopulation. **F**: OMIM genes incompletely assembled in 50 or more samples using either Flye (orange) or Shasta-Hapdup (blue). For Shasta-Hapdup, if one haplotype was completely assembled in a sample but the other was incomplete, the gene is counted as incompletely assembled. Assembly of 5 genes (*FAM20C*, *HYDIN*, *NOTCH2NLC*, *PRKAR1B*, and *SHANK2*) was incomplete for all 100 samples using both assemblers. Genes that are not in or do not contain a segdup are in bold with an asterisk.

We investigated why many of the Flye assemblies had similar contig NG50 values by plotting the contig breakpoints for both the Shasta-Hapdup and Flye assemblies. Among the 100 Flye assemblies, 97.1% of assembly breaks occurred within regions annotated as segmental duplications (segdups), satellite sequence, or both while 2.9% occurred within nonrepetitive sequence **(Table S4)**. Among the 2.9% of assembly breaks in nonrepetitive sequence, 90% were seen only in a single sample, suggesting stochastic artifacts of the assembly process. Performing a focused analysis of chromosome 7 revealed an increased number of contig breaks in the telomeric and pericentromeric regions for both Flye and Shasta-Hapdup assemblies **(Figure 2D)**, and at positions flanking well-described recurrent copy number changes associated with disease (Morris 1993). Among all contig breaks on chromosome 7, 1.9% of those in Flye assemblies were breaks in nonrepetitive sequence that are not within 10 kbp of a segdup, similar to the genome-wide average of 2.9%. This was surprising, and visual analysis of these regions did not reveal sample-specific differences that would easily explain the break in assembly, such as a duplication, inversion, or increased number of SNVs, suggesting that local sequence variation did not influence the position of assembly breaks in these nonrepetitive regions **(Figure S4)**.

We then evaluated contig size across superpopulation groups and assembly of disease-associated OMIM genes. Calculating the median contig size per sample excluding contigs <1 Mbp **(Figure 2E)** revealed a higher median contig size for African samples compared to all non-African samples, which was expected given the higher genetic diversity of the African samples. We then evaluated how well disease-associated genes were assembled in these samples. Among 4,615 disease-associated OMIM genes (excluding genes on the X and Y chromosomes), we found that 2.7% (123/4,615) and 3.0% (140/4,615) of genes in the Flye or Shasta-Hapdup assemblies, respectively, were incompletely or incorrectly assembled (i.e., they were not spanned by a single, complete contig) in at least 5 samples **(File S1)**. Among the 200 assemblies (100 Flye and 100 Shasta-Hapdup), we found that 5 OMIM genes were incompletely assembled in all 200 assemblies and another 45 OMIM genes were incompletely assembled in at least 50 or more of the 200 assemblies **(Figure 2F)**. We observed more incompletely assembled genes in the Shasta-Hapdup assemblies, partly due to the requirement for a single gene to be entirely spanned by a single contig in both haplotypes for it to be considered fully assembled.

We subsequently applied PGGB to construct chromosome-level pangenome graphs from the 100 Shasta-Hapdup assemblies and generate multisample variant calls including all types of variants (Garrison et al. 2023). To investigate the differences between assembly approaches, we performed principal component analysis (PCA) on a chromosome 20 pangenome graph created by combining the 100 Shasta-Hapdup assemblies with 44 assemblies from the HPRC (Liao et al. 2023). The PCA showed a clear separation between the two pangenomes (**Figure S5A**). However, a PCA based on the euchromatic, noncentromeric fraction of the chromosome 20 graph demonstrates that this difference is primarily due to the improved resolution of highly repetitive sequences by the HiFi-based HPRC assemblies (**Figure S5B**), supporting the high-quality nature of our assemblies.

### Variation within active transposable elements

The largely repetitive and polymorphic nature of active transposable elements, especially full-length *L1* and *ERV*, makes them challenging to fully resolve and characterize using short-read assemblies (Yang et al. 2024). We anticipated that long-read assemblies would allow us to overcome these challenges. Using RepeatMasker (http://www.repeatmasker.org/), we identified interspersed repeats in the 100 Shasta-Hapdup assemblies and found that the fraction of major interspersed repeats differs by no more than 3% compared to that of the T2T-CHM13 assembly (Nurk et al. 2022) **(Table S5).** Furthermore, there was minimal variation among the 100 assemblies in interspersed repeat content.

Among the youngest polymorphic interspersed repeats that are too long to resolve with short reads (Chaisson et al. 2019), LINE-1s (∼6,000 bp) are the only type that are actively expanding in the human genome. We found that the total base pairs of LINE-1 sequence (including young and old LINE-1s) in the 100 assemblies (496 Mbp average) is lower than observed in the CHM13 T2T assembly (512 Mbp), likely due to LINE-1s within unassembled regions. To measure the ability of these ONT-based assemblies to resolve young LINE-1s, we calculated the number of the youngest LINE-1 elements (L1HS) and the number of full-length (≥6 kbp) L1HS elements. Overall, we found similar numbers of L1HS and full-length L1HS sequences compared to HG002 and HG005 from GIAB and the CHM13 T2T assembly **(Figure S6)**. Although HERV-Ks (∼9,000 bp) are unlikely to be actively replicating in modern humans, like LINE-1s, they are known to be polymorphic in the human population (Li et al. 2019; Subramanian et al. 2011). Therefore, we also counted the number of full-length HERV-Ks (HERVK-int) and found that the number per genome is similar among the 100 assemblies and CHM13 T2T, HG002, and HG005. This demonstrates that these assemblies are of sufficient quality to resolve the youngest long interspersed repeats and that there is variation in the number of these insertions among different human populations.

### Structural variant analysis

We called SVs using four alignment-based and one assembly-based method **(see Methods)**. To validate our SV-calling methods, we compared them against a known set of SV calls generated by the HPRC (Liao et al. 2023). We generated SV calls from three of the five genomes used for small variant benchmarking (HG002/NA24385, HG00733, and HG02723) and identified an average of 23,732 SVs across all five callers. This is similar to the average of 22,755 SVs among 15 human genomes assembled by Audano *et al*. (2019) but less than those predicted by the HPRC and HGSVC (Ebert et al. 2021; Liao et al. 2023). The greater number than Audano is expected given that they were called with older PacBio chemistries (RSII CLR) and an approach, SMRT-SV, that excluded SV calls in some pericentromeric regions or regions where variant calls were considered less reliable (Audano et al. 2019). Benchmarking against the HPRC Sniffles2 SV calls (Liao et al. 2023) and restricting calls to regions within the GIAB HG002 SV Tier1 v0.6 benchmarking regions (GIAB Tier1 Regions) (Zook et al. 2020) revealed F1 scores greater than 90% for both methods among all three samples **(Figure 3A)**. When comparing genome-wide SV calls (not restricted to the GIAB Tier1 regions) our F1 score decreased to approximately 70% for all three samples, suggesting difficulty in generating concordant SV calls in low-complexity or repetitive regions of the genome **(Table S6)**.

**Figure 3.**
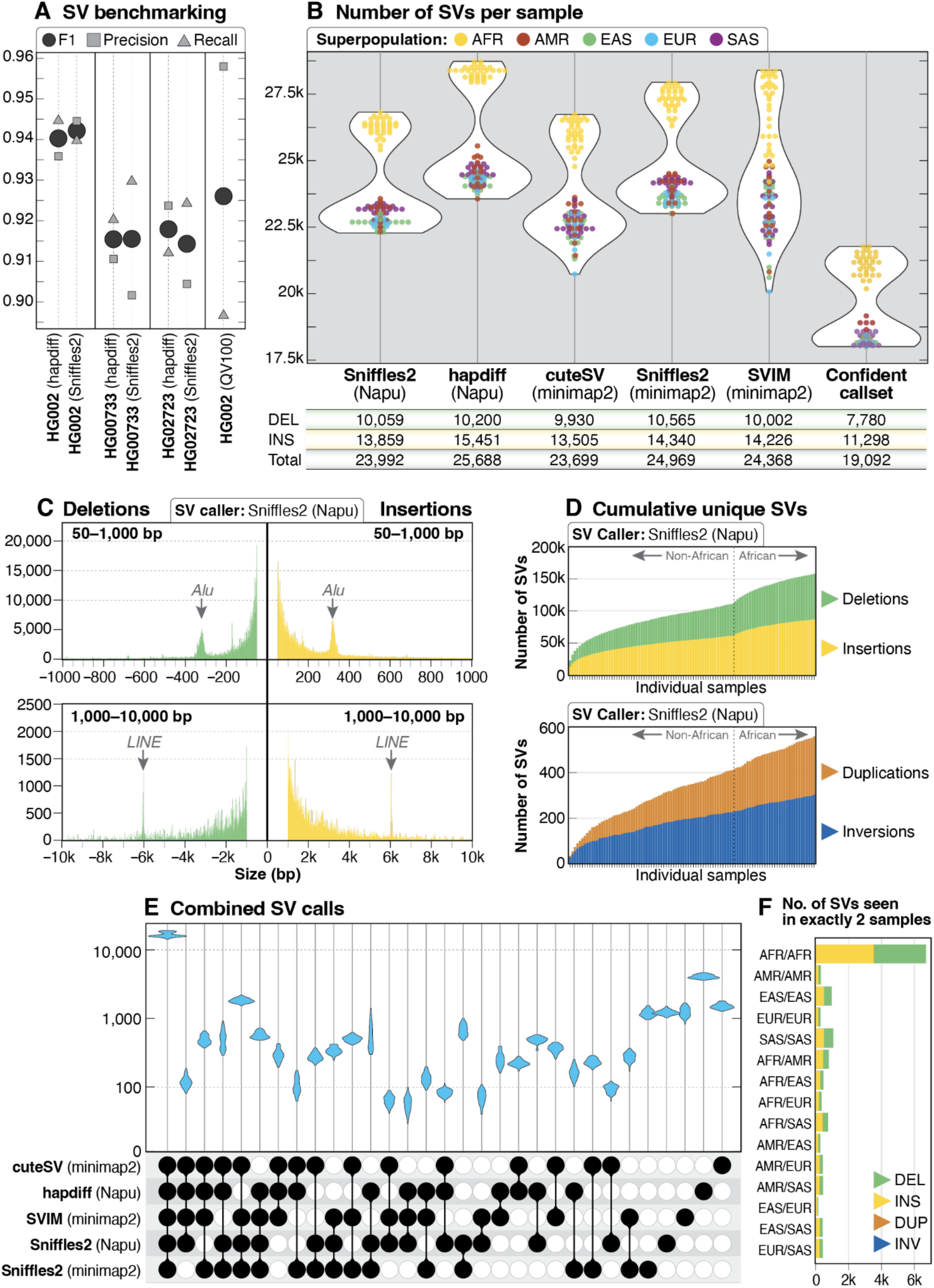
SV call set. **A:** SV calls were benchmarked against HPRC Sniffles2 SV calls within the GIAB HG002 SV Tier1 benchmarking regions. **B**: A similar number of genome-wide SVs were identified by all five callers used in this study, with the highest number of SVs per sample identified by hapdiff. The confident call set is defined as variants called by hapdiff and at least 2 unique alignment-based callers. For each call set the average number of deletions (DEL), insertions (INS) and total SVs (including INV, DUP and BND events) per sample is shown below the plot. **C:** Histogram of insertion and deletion counts stratified by size using Sniffles2 from the Napu pipeline. The peak around 300 bp represents *Alu* insertions or deletions, and the peak around 6 kbp represents LINE insertions or deletions. **D:** Cumulative novel SVs per sample. The frequency of new SVs observed increases when samples from individuals of African ancestry are included. **E:** Upset plot of overlap among SV callers after merging with Jasmine. For each sample, 5 vcf files were merged, demonstrating that the majority of calls in each sample were called by all 5 callers. The next highest violin plots are calls made by all callers except for hapdiff (the only assembly-based caller) and calls made only by one caller. **F:** Among 113,696 SVs from the Jasmine-merged confident call set, 12,432 were found in exactly 2 samples, with 6,181 (50%) of those calls in pairs in which both samples are from the African superpopulation.

We observed high per-caller concordance between the number of SV calls from the three benchmarking genomes and the 100 genomes presented here **(Figure 3B)**. Across the five callers, we identified an average of 24,543 SVs per sample (min: 20,068, max: 28,734), similar to the 23,000–28,000 SVs per sample reported by the HGSVC (Ebert et al. 2021). Consistent with prior work, we observed more total SV calls in samples from the African superpopulation (Ebert et al. 2021; Audano et al. 2019; 1000 Genomes Project Consortium et al. 2015). The distribution of insertions and deletions called in this dataset was also as expected, with peaks around 300 bp for Alu elements and at 6 kbp for LINE elements **(Figure 3C)**. A generally proportional number of SVs per chromosome was observed and, on average, more insertion than deletion events were identified per chromosome for all SV callers **(Figure S7)**. The distribution of total SV events genome-wide was similar to prior work, with an increased number of insertions and deletions near the telomeres and centromeres **(Figure S8)**. We identified an increasing number of novel SVs, excluding breakends (BNDs), for each additional sample sequenced among all SV callers **(Figure 3D)**.

Because a primary goal of our study is to identify and catalog high-quality SVs among the 1KGP samples, we merged the SVs from each of the five SV callers per sample using Jasmine (Kirsche et al. 2023). We observed high concordance between SV callers across all samples **(Figure 3E)**, with an average of 16,722 SVs per sample called by all callers and no individual sample having an SV type that was noticeably higher or lower than other samples within the same superpopulation **(Figure S9A)**. Analysis of SVs called by at least four, three, two, or one callers per sample identified an average of 20,242, 22,685, 25,540, and 34,796 SVs, respectively **(Figure S9B)**.

The SVs per sample called exclusively by hapdiff represent the majority of SVs called exclusively by a single caller. Because hapdiff was the only assembly-based caller in our dataset, we examined whether these calls represented false positives or SVs in regions where alignment may be challenging. Our analysis found that of the 407,779 SVs (excluding BNDs) called only by hapdiff across all 100 samples (i.e., not merged), 151,575 (37.1%) were fully or partially within a segdup or within 1,000 bp of a segdup, suggesting that they may be in complex copy-number polymorphic regions of the genome, and thus potential artifacts because of their proximity to a segdup. Of the SVs that were not fully or partially within a segdup or within 1,000 bp of a segdup, 119,255 (46.5% of the remaining SVs) overlap a VNTR region. Analysis of SVs called only by hapdiff did not reveal any individual sample or population outliers **(Figure S9C)**, and visual analysis of 30 randomly selected SVs from this set found that 28/30 were likely false-positive calls. These false-positive SV calls were either located within centromeric (6/30), telomeric (8/30), or other low-complexity regions (3/30), or they had allelic representations different from those observed with alignment-based methods (10/30) **(Figure S10)**. This suggests that difficult-to-assemble regions are a major source of false-positive assembly-based SV calls.

An SV frequency call set was generated that represented SVs called by all five callers (100,915 total SVs), four or more (119,805 total SVs), three or more (133,766 total SVs), two or more (155,407 total SVs), or at least one caller (252,954 total SVs). Among the 100 samples described here, there were a total of 113,696 shared or unique high-confidence SVs (SVs identified by hapdiff and 2 or more unique callers, excluding BNDs), with 32% found in only one sample (36,096 of 113,696). We found that 12,432 (11%) of these shared SVs were seen in exactly 2 samples, and that approximately half of these shared SVs were in samples only from the African superpopulation **(Figure 3F)**, similar to previous analysis (1000 Genomes Project Consortium et al. 2015). Among 50,458 high-confidence SVs that intersect protein-coding genes, 97% (49,142/50,458) are within or include intronic sequence, 3.3% (1,654 / 50,458) are within or include coding sequence, and 2.0% (992/50,458) are within or include a 5’ or 3’ untranslated region (UTR).

To investigate the functional significance of SVs intersecting protein-coding genes, we performed an SV-eQTL analysis using the merged SV call sets and the recently published MAGE RNAseq dataset, which was derived from the same HapMap samples (Taylor et al. 2023). This revealed a strong overlap with SVs and SV-eQTLs previously found using a collection of 31 diverse LRS-based genomes (Kirsche et al. 2023), including a 1,235-bp deletion associated with *TNFSF13*, a gene implicated in a number of cancers and autoimmune diseases (Chen et al. 2021; Ortiz-Aljaro et al. 2022). This analysis revealed several new associations, including a 96-bp deletion not previously detected by Kirsche *et al*. (2023) that is associated with the *IFFO2* gene, a potential factor in breast cancer susceptibility (Danforth 2016) **(Figure S11)**.

### Structural variation within medically relevant genes

Sequencing of samples from all five superpopulations allowed us to evaluate population-specific SVs intersecting genes associated with an OMIM phenotype (n = 4,866) and revealed 349 high-confidence SVs in or including at least one defined exon **(Figure S12A, Table S7)**. These events ranged in size from 50 bp – the lower limit for a variant to be considered an SV – (deletions in *TNFRSF13C* and *TF* and an insertion in *IMPG2*) to 87,776 bp (a deletion that fully includes *IGHM*). Visual analysis of 30 randomly selected events confirmed that all were likely to be true positives. These 349 SVs are distributed across all chromosomes and impact 335 exons in 236 unique OMIM genes, with 123 of those 335 exons containing ClinVar variants that are annotated as pathogenic or likely pathogenic **(Figure S12B)**. We found that 150/349 (43%) of these SVs are found in only one sample, and no single sample has more than 6 unique SVs (HG01369). Three SVs (a 458-bp insertion in *ABCC11*, a 243-bp insertion in *XYLT1*, and a 118-bp insertion in *MED13L*) are seen in all 100 samples, suggesting the reference genome represents a minor allele at some or all of these positions. Indeed, GRCh38 has been patched to include a similar insertion in *XYLT1*. We then evaluated whether SVs intersecting genes also clustered within specific superpopulations and found that of the 38 SVs observed in only 2 samples, 76% (29/38) were superpopulation-specific with 55% of those (16/29) seen in samples from the African superpopulation.

In this dataset, we observed 4 SVs spanning multiple genes, some of which are known population variants. This includes a 22.8-kbp deletion spanning *HBB*, *HBD*, and *HBG1* **(Figure 4A)** associated with beta thalassemia (Huisman et al. 1972) (MIM: 613985) and two samples with a 19,304-bp deletion including *HBA1* and *HBA2* commonly referred to as the Southeast Asian deletion (Farashi and Harteveld 2018) (MIM: 604131) (**Figure S13)**.

**Figure 4.**
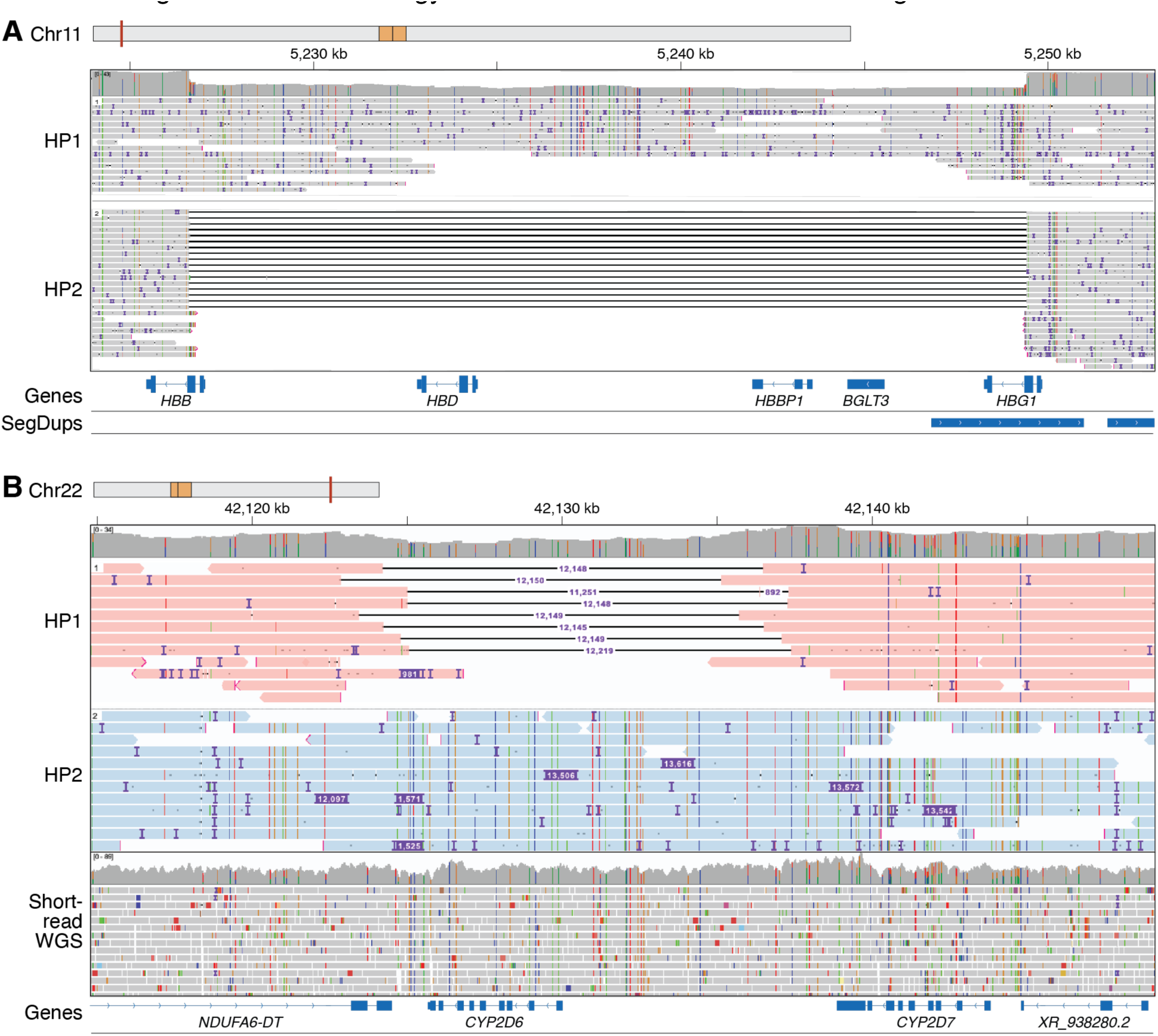
SVs, including multi-exon deletions are found in medically relevant genes. **A.** Phased IGV view of a 22,791-bp deletion in GM19035 that includes all or part of HBB, HBD, HBBP1, BGLT3, and HBG1. Variants in this region are associated with beta thalassemia (MIM: 613985) with this specific deletion known as Hemoglobin Kenya with this individual likely being an asymptomatic carrier (Huisman *et al*. 1972). GM19035 is from an individual from the Luhya population within the African superpopulation. **B.** Phased IGV view from LRS data showing a *CYP2D6* full gene deletion on one haplotype (HP1) and a hybrid tandem arrangement (*\*36+*10*) represented by an insertion on the second haplotype (HP2) in HG02396, compared to short-read whole genome sequence data from the same sample in which the complex nature of this event cannot be resolved.

We did not expect to find rare SVs in X-linked OMIM genes in 46,XY samples, since those events would be more likely to be associated with a disease. However, five SVs were found in our confident set that intersected an X-linked OMIM gene in at least one 46,XY sample. Of these, four are in a 3’UTR and were observed in at least two 46,XX samples. One of the four events, found in only one sample, was an approximately 141-bp insertion in exon 15 of *RPGR* (OMIM: 312610), a gene associated with several X-linked conditions including retinitis pigmentosa, cone-rod dystrophy, and macular degeneration (Fahim et al. 1993). A similar insertion at this position has been reported twice in ClinVar as a variant of uncertain significance (VUS) associated with primary ciliary dyskinesia, once as a 141-bp insertion (ClinVar entry 2121719) and once as a 69-bp insertion (ClinVar entry 1975740). Evaluation of the short-read sequencing data for this sample at this position did not clearly demonstrate the insertion, but the insertion consists of only C– and T-nucleotides, which makes it difficult to align and evaluate using short-read technology **(Figure S14)**. The presence of this insertion in a 46,XY 1KGP sample suggests that this variant may be present at a higher allele frequency than expected, is difficult to reliably call using short-read technology, or could be associated with a later onset of the associated phenotype.

Next, we wondered how many SVs we would find in genes that are often difficult to evaluate using short-read technology and found a substantial number of high-confidence SVs in challenging regions of the genome. For example, 42% (47,315/113,696) of the high-confidence SVs occur fully outside of the GIAB Tier 1 regions, and visual inspection of 30 events confirmed the presence of an SV. We also identified 407 high-confidence SVs within coding regions defined as unreliable for variant identification using short-read sequencing based on analysis of gnomAD data (Hijikata et al. 2024). In both cases, these SVs reside in regions that may be filtered by variant annotation pipelines. Finally, 9,788 of the high-confidence insertions were ≥ 500 bp, which may preclude accurate resolution of these events and limit our understanding of their impact on gene expression or splicing when evaluated using short-read technology.

Cytochrome P450 (CYP) genes are among the gene sets that are challenging to interrogate using short-read technologies and may require separate variant calling approaches to fully evaluate (Lee et al. 2019). There is interest in characterizing star alleles (haplotypes of SNVs and structural variation) in these pharmacogenes, as they are known to impact drug response (Zanger and Schwab 2013). Within this dataset, we observed that LRS enabled better resolution of full gene deletion and duplication SV events in highly polymorphic CYP pharmacogenes such as *CYP2D6*, a major pharmacogene involved in the metabolism of over 20% of clinically prescribed medications (Zanger and Schwab 2013). For example, we identified one individual (HG02396) with a *CYP2D6* gene deletion (*\*5*) on one haplotype and a hybrid tandem arrangement (*\*36+*10*)—shown via an insertion—on the second haplotype (**Figure 4B)**. In the equivalent short-read WGS data, it can be difficult to identify both the gene deletion and the hybrid tandem star allele in the same individual. Separate analysis of a complex *CYP2B6* star allele (*CYP2B6*29*) identified in previous short-read analysis (Twesigomwe et al. 2024) showed that it was called by hapdiff but not the alignment-based callers used in this study, demonstrating that some of these complex alleles may not be represented in our initial high-confidence SV set (**Figure S15**).

Finally, because a major goal of this project is to create a dataset that can be used for filtering and prioritizing disease-associated SVs, we tested whether the SVs identified in these first 100 samples could be used to accurately filter SVs in cases with known disease-associated SVs. We used Jasmine to identify unique SVs—those not found in the high-confidence set from the 100 samples presented here—in 16 positive control cases known to carry a pathogenic SV previously identified by whole-genome (8 cases) or targeted (8 cases) ONT sequencing (Wilderman et al. 2024; Miller et al. 2021) **(Table 1)**. Among the 8 cases that had undergone whole-genome LRS, filtering reduced the average number of high-confidence SVs by 93% (from 22,743 to 1,664), and in all 16 cases the pathogenic SV was retained after filtering. This demonstrates that the high-confidence SV calls generated here can be used to filter and prioritize disease-causing SVs in cases where there is a high suspicion of a Mendelian condition.

**Table 1.** Results from filtering known disease-associated SVs using SV calls from 1KGP. SV lengths may be rounded or approximate.

| Number | Gene | SV type (type or length) | Beginning SV Count | SVs remaining after filtering | Remaining SVs in genic regions | Filtering Result |
| --- | --- | --- | --- | --- | --- | --- |
| **Targeted long-read sequencing** |  |  |  |  |  |  |
| 1 | *ALMS1* | Insertion (*Alu*)<sup>a</sup> | N/A | N/A | N/A | Present |
| 2 | *HPS1* | Deletion (1.9 kbp)<sup>a</sup> | N/A | N/A | N/A | Present |
| 3 | *AGL* | Deletion (1.5 kbp)<sup>a</sup> | N/A | N/A | N/A | Present |
| 4 | *HYDIN* | Deletion (1.7 kbp)<sup>c</sup> | N/A | N/A | N/A | Present |
| 5 | *HYDIN* | Deletion (10 kbp)<sup>c</sup> | N/A | N/A | N/A | Present |
| 6 | *Multiple* | Duplication (2 Mbp)<sup>b</sup> | N/A | N/A | N/A | Present |
| 7 | *TP53* | Insertion (300 bp)<sup>c</sup> | N/A | N/A | N/A | Present |
| 8 | *HPRT1* | Inversion (17 Mbp)<sup>a</sup> | N/A | N/A | N/A | Present |
| **Whole-genome long-read sequencing** |  |  |  |  |  |  |
| 1 | *NSD1* | Insertion (110 bp)<sup>c</sup> | 18422 | 1584 | 685 | Present |
| 2 | *FGA* | Deletion (4.1 kbp)<sup>c</sup> | 22834 | 1999 | 269 | Present |
| 3 | *FANCD2* | Deletion (410 bp)<sup>c</sup> | 22933 | 1503 | 327 | Present |
| 4 | *ABCD1* | Insertion (2.8 kbp)<sup>c</sup> | 23216 | 1757 | 557 | Present |
| 5 | *DBT* | Deletion (3 kbp)<sup>c</sup> | 23800 | 3194 | 1361 | Present |
| 6 | *AGRN* | Deletion (2.3 kbp)<sup>c</sup> | 23346 | 419 | 171 | Present |
| 7 | *COL5A1* | Inversion (1.5 Mbp)<sup>c</sup> | 24361 | 2038 | 417 | Present |
| 8 | *AGL* | Insertion (*Alu*)<sup>c</sup> | 23033 | 820 | 259 | Present |

SVs reported in other studies include: <sup>a</sup> Miller *et al.* 2021, <sup>b</sup> Wilderman *et al.* 2024, <sup>c</sup> unpublished.

### Analysis of disease-associated repeat expansions

Tandem repeat expansions—such as short tandem repeats (STRs) and variable number tandem repeats (VNTRs)—at more than 60 loci have been implicated in human diseases such as the GGC expansion in the 5’UTR of *XYLT1* (MIM: 608124) associated with Baratella-Scott syndrome (MIM: 300881) or the intronic GGGGCC expansion in C9orf72 (MIM: 614260) associated with amyotrophic lateral sclerosis and/or frontotemporal dementia (MIM: 105550) (Depienne and Mandel 2021; Hannan 2018; Tanudisastro et al. 2024; Chaisson et al. 2023). Many pathogenic repeat expansions associated with Mendelian disease are difficult to size or fully sequence-resolve using short-read sequencing methods, meaning clinically relevant interruptions in the repeat may not be easily identified (Chaisson et al. 2023; Tanudisastro et al. 2024). Thus, there is interest in using LRS to evaluate repeat expansions genome-wide and at clinically relevant loci (Dolzhenko et al. 2023; Reis et al. 2023; Sulovari et al. 2019).

We used vamos to perform genome-wide haplotype-resolved analysis of 562,005 loci— including 66 loci associated with disease—that consisted of both simple and complex repeat units **(Figure 5A, Figure S16, File S2)**. Using these data, we identified candidate pathogenic-sized expansions in three genes: *RFC1*, *ATXN10*, and *FGF14*. Expansions in *RFC1*, which are associated with autosomal recessive cerebellar ataxia, neuropathy, vestibular areflexia syndrome (CANVAS, MIM #614575), were observed in five samples ranging from 359 to 712 repeat units in size **(Figure 5B, Figure S17)**. Pathogenic expansions in this gene are typically 400 repeat units or larger and are motif-dependent, with expansions of an AAGGG repeat being the most common pathogenic expansion (Cortese et al. 2019; Scriba et al. 2020; Beecroft et al. 2020). Our observation that some of these samples carried the AAGGG repeat unit while others carried a nonpathogenic repeat unit, such as AAAAG, was similar to recent work that identified expansions in *RFC1* of varying repeat motifs in 5 of 100 HPRC samples (Dolzhenko et al. 2023; Cortese et al. 1993) **(Figure 5C)**. That we observed an expansion in 5% of samples was not unexpected, as the carrier frequency of *RFC1* expansions has been reported to be 1–5% across at least two populations (Cortese et al. 2019; Fan et al. 2020; Akçimen et al. 2019).

**Figure 5.**
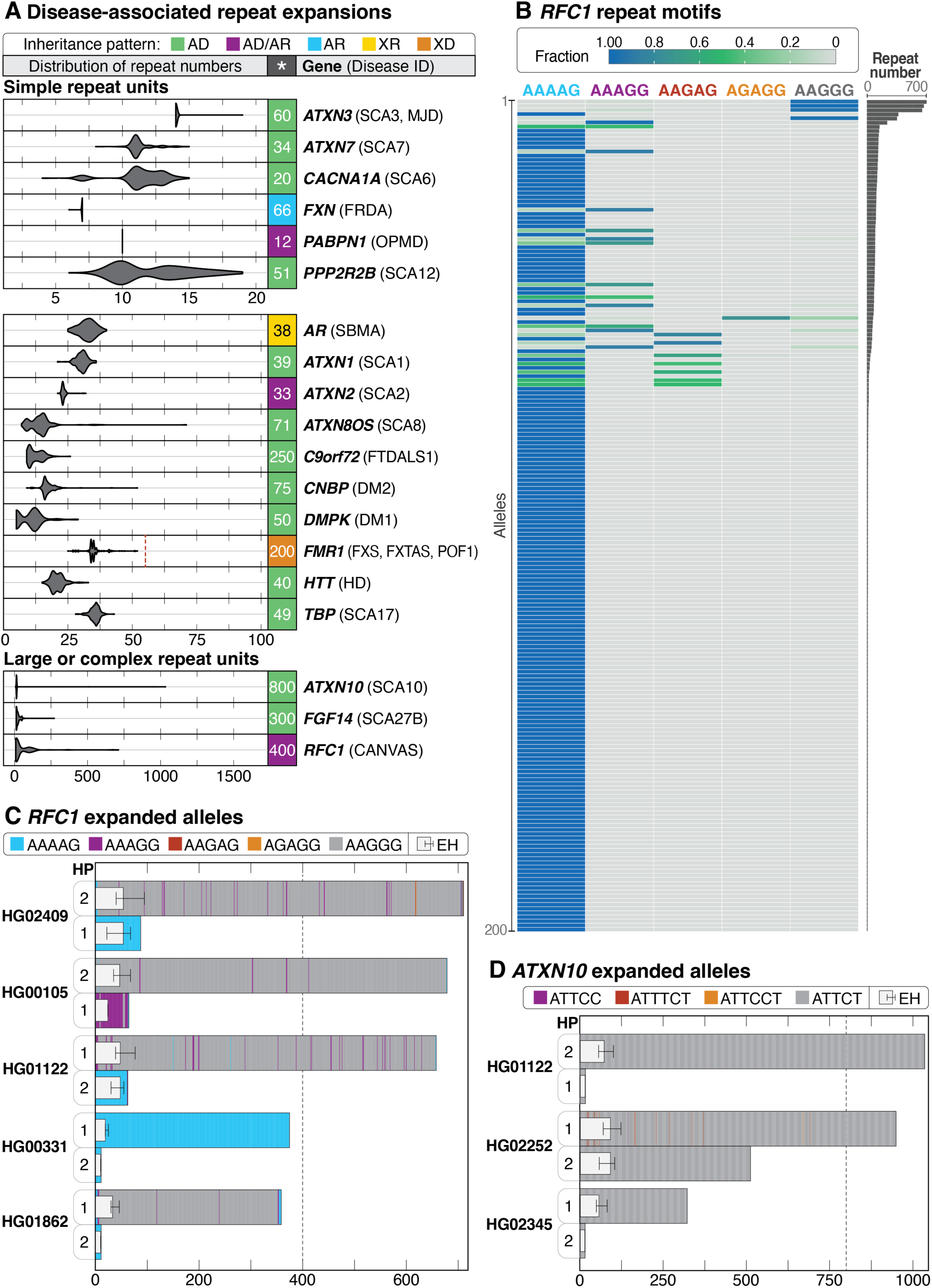
Evaluation of repeat expansions known to be associated with Mendelian conditions. **A:** Haplotype-resolved repeat expansions of selected repeat loci for simple and complex repeat units. Pathogenic repeat size is shown to the right of each plot (*), the associated condition is in parentheses, and the full name of each condition can be found in Table S10. The pathogenic repeat size for *FMR1* is listed as 200 repeats, but a dashed vertical line represents the 55-repeat threshold that puts 46,XX and 46,XY individuals at risk for fragile X-associated tremor/ataxia syndrome (FXTAS, MIM #300623) and 46,XX individuals at risk of fragile X-associated primary ovarian insufficiency (POF1/FXPOI, MIM #311360). (AD, autosomal dominant; AD/AR, autosomal dominant/recessive; AR, autosomal recessive; XR, X-linked recessive; XD, X-linked dominant.) **B:** Among 200 haplotypes (*y*-axis), an expansion in *RFC1* near or over 400 repeat units was seen in 5 haplotypes. The fraction of each motif within a single haplotype is shown. AAGGG is the most common pathogenic repeat expansion; additional pathogenic expansions include ACAGG (not shown), and a mixed AAAGG/AAGGG expansion.(Cortese et al. 1993) **C:** Haplotype (HP)-resolved detail of *RFC1* repeat expansions in five samples with an expansion of one allele. Haplotypes are assigned arbitrarily. Dotted line represents the position of full penetrance alleles typically seen at 400 repeat units. **D:** Three samples with expansions in *ATXN10* larger than 280 ATTCT repeats were observed, one of which carries one allele larger than 800 repeat units and one allele around 500 repeat units in size. The dotted line at 800 repeat units represents the position of the lower end of the full penetrance range. ExpansionHunter (EH) estimates are overlayed atop the bar plots in (C) and (D), placed on HP1 or HP2 based on their length.

Expansions were also observed in *ATXN10*, which is associated with autosomal dominant spinocerebellar ataxia type 10 (SCA10, MIM #603516), a slowly progressive ataxia with typical age of onset between 12 and 48 years and full-penetrance alleles varying from 800 to 4,500 ATTCT repeats (Matsuura and Ashizawa 1993; Alonso et al. 2006; Raskin et al. 2007). Two of the 100 samples were heterozygous for alleles larger than 800 motifs, one of which had a second allele with 511 repeat units **(Figure 5D, Figure S18)**. In addition, two other samples harbored expansions close to or larger than 280 repeat units, which has been reported as causative in one individual with ataxia (Matsuura et al. 2006). However, three of the four large alleles are purely ATTCT and there is evidence suggesting that interruptions of ATTCC are necessary for the allele to be pathogenic (Morato Torres et al. 2022).

Finally, we also identified a single sample with an expansion of 272 GAA units in *FGF14*, the locus associated with autosomal dominant spinocerebellar ataxia 27B (SCA27B, MIM: 620174) (Pellerin et al. 2023), within the range (250–400 repeat units) previously reported to be associated with reduced penetrance **(Figure S19)**.

To determine whether any of the 10 expansions (5 expanded *RFC1* alleles, 5 expanded *ATXN10* alleles) would be identified using short-read data, we ran ExpansionHunter on short-read data from all affected samples (Dolzhenko et al. 2023). In all cases, when an expanded allele was present, the corresponding ExpansionHunter estimate was larger than the normal allele but, in most cases, still significantly underestimated the size of the expansion **(Figure 5C**, **Figure 5D, Table S8)**. For example, in *ATXN10*, LRS identified a normal allele (15 repeat units) and an expansion in *ATXN10* of more than 1,000 repeat units in HG01122. The ExpansionHunter estimates for this position in this sample are 15 repeat units (range 15–15) and 73 units (range 56–101), thus the normal allele was correctly estimated but the expanded allele was markedly underestimated.

### Evaluation of genome-wide methylation patterns and identification of novel differentially methylated loci

A major advantage of LRS is the ability to simultaneously capture both DNA sequence and modification information (Logsdon et al. 2020). This allows for a more detailed evaluation of how changes in sequence, such as a repeat expansion or transposable element insertion, may alter the local epigenetic landscape and may identify variants missed by prior analysis or lead to prioritization of an unexpected locus. We evaluated methylation both genome-wide and at loci associated with imprinting disorders. Among 69 of the 70 46,XX samples sequenced, we found that 39% (27/69) had X-chromosome methylation patterns suggesting skewed X-inactivation (**Figure 6A**, **Table S9**).

**Figure 6.**
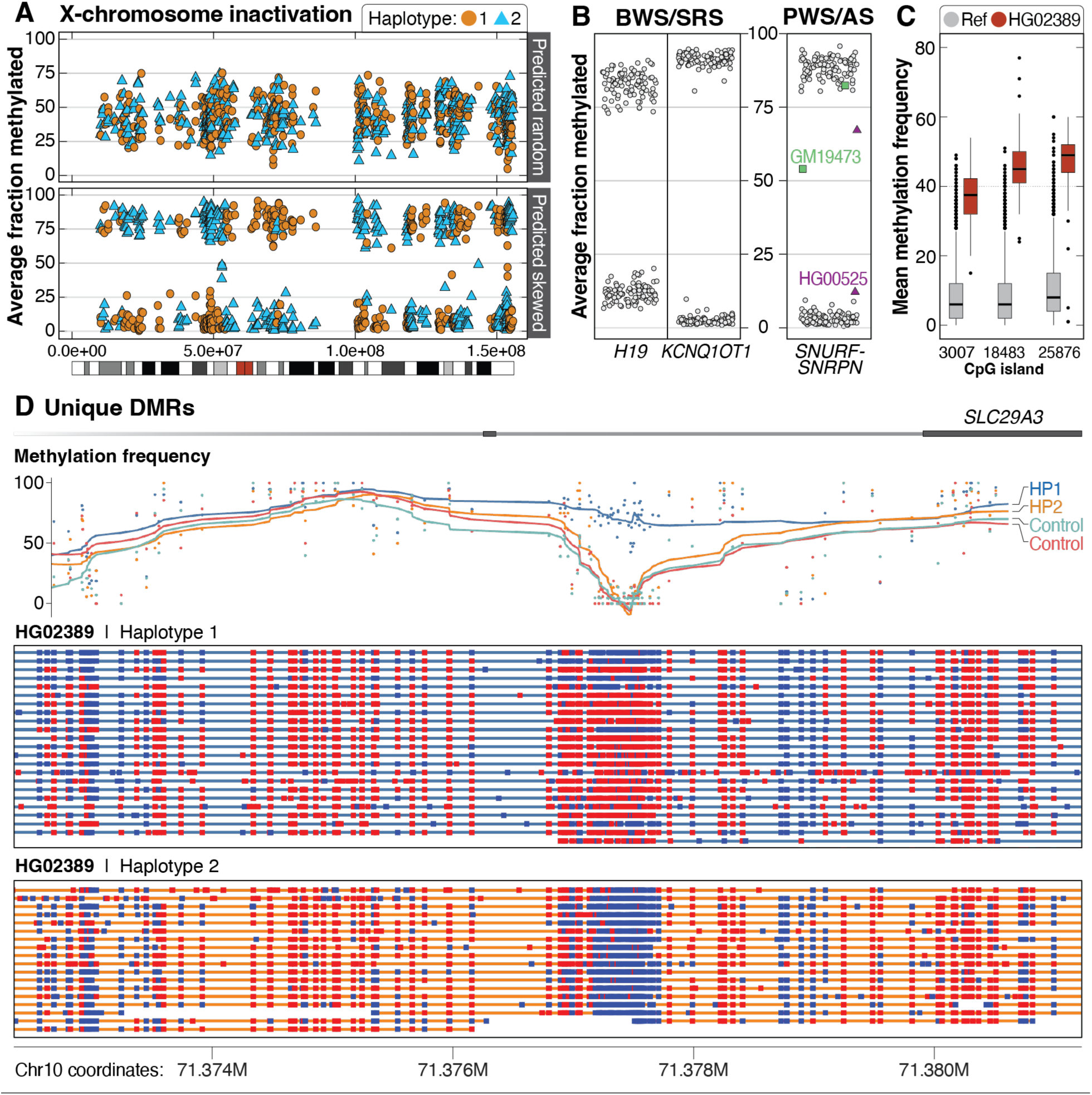
Patterns of methylation among the 1000 Genomes samples. **A.** Among 69 46,XX samples, 42 had mixed X-chromosome inactivation (top, example from HG01414), while 27 were skewed (bottom, example from HG01801). The color differences are related to breaks in phasing and do not suggest methylation is mixed along a single haplotype. **B.** Haplotype-resolved methylation fraction is shown for three imprinted loci associated with four imprinting disorders. Methylated (>75%) or unmethylated (<25%) fraction at IC1 in *H19* and IC2 in *KCNQ1OT1*, which are associated with BWS and SRS on 11p15.5. Haplotype-resolved methylation fraction is also shown for the CpG island within *SNURF-SNRPN* that is evaluated when testing for PWS or AS. Two samples have either gain (GM19473) or loss (HG00525) of methylation at this locus. **C.** Unique distinct methylation differences within defined CpG islands were identified in individual samples. An example from HG02389 shows three CpG sites with increased methylation (red boxes) compared to controls (gray). **D.** As an example, we identified one haplotype in HG02389 that has increased methylation at an internal CpG site (3007) within an intron of *SLC29A3*. Methylation frequency by haplotype is shown for HG02389 and one control (HG03022). Methylation status is shown for individual reads for each haplotype from HG02389 only (red indicates a methylated CpG; blue indicates an unmethylated CpG).

We then performed genome-wide principal component analysis of methylation to evaluate whether samples would correlate with ancestry or if patterns of X-inactivation would be apparent **(Figure S20)**. This analysis revealed one sample, GM18864, that clustered with 46,XY samples but was reported to be 46,XX. Because we validated each sample using SNVs from short-read sequencing, we wondered whether this sample had lost, or was losing, an X-chromosome and found that the average X chromosome depth of coverage was approximately 55% of the full-length autosomes in the LRS data, and approximately 75% in the short-read sequencing data, suggesting loss of an X chromosome in this sample (Pedersen et al. 2020).

Next, we evaluated methylation patterns at two disease-associated loci: 11p15.5, which is associated with both Beckwith-Wiedemann Syndrome (BWS, MIM #130650) and Silver-Russell syndrome (SRS, MIM #180860) (Saal et al. 1993; Shuman et al. 1993); and 15q11.2-q13, which is associated with both Prader-Willi syndrome (PWS, MIM #176270) and Angelman syndrome (AS, MIM #105830) (Dagli et al. 1993; Driscoll et al. 1993). Each of these regions contains CpG islands known to be differentially methylated depending on which parent the locus is inherited from. For the 11p15.5 region, we found that in all samples, one haplotype was completely methylated while the other was completely unmethylated at two imprinting centers known as IC1 and IC2 **(Figure 6B)**. Evaluation of haplotype-resolved methylation at the *SNURF-SNRPN* locus on 15q11.2 revealed two samples, GM19473 and HG00525, where one haplotype was 25%–75% methylated. Visual evaluation of these samples showed that one haplotype of GM19473 had increased methylation while one haplotype of HG00525 had reduced methylation, which was unexpected and further demonstrates that changes in methylation can occur throughout the genome in these cell lines, even at well-established differentially methylated loci **(Figure S21)**.

We used MeOW to analyze differences in methylation at CpG sites genome-wide and identified 134 CpGs with methylation differences across 37 samples, with a median of 2 DMRs per sample. Among the 134 sites, 125 were observed in only one sample, 8 were seen in 2 samples, and 1 CpG had a distinct methylation pattern in 3 samples **(Table S10)**. As an example, 3 DMRs were found in HG02389 **(Figure 6C)**, including a hypermethylated CpG in *SLC29A3* not present in controls **(Figure 6D)**. We observed both hypermethylation (86 CpGs) and hypomethylation (48 CpGs) among the 134 CpGs and identified four samples with more than 10 DMRs, with 3 of 4 having mixed patterns of methylation (GM19462, HG03548, and HG02817) and one (GM19473) with only increased methylation **(Figure S22)**. We investigated whether changes in gene expression were associated with these changes and found that among the 15 samples from the African superpopulation with a DMR, there was an enrichment of expression outliers near the DMR with increasingly stringent z-score thresholds, suggesting changes in gene expression corresponding to these methylation changes **(Figure S23)**.

## DISCUSSION

Current approaches to clinical genetic testing are incomplete as they are unable to capture the full spectrum of disease-causing variation (Wojcik et al. 2023). These limitations exist because: 1) new technologies, such as LRS, are not yet widely implemented in the clinical testing lab; 2) computational tools are not yet able to efficiently capitalize on the data provided by these new technologies and those that can have substantial computational requirements; and 3) databases for filtering and prioritizing variants identified using new technologies are not yet available. The 1KGP-ONT Consortium plans to sequence at least 800 1KGP samples to characterize variation in genomic regions that are difficult to evaluate using short-read technology. Sequencing ≥800 samples will capture a more complete catalog of variation, especially rare yet presumably benign variants across the 1KGP populations. The expanded collection will enable a more accurate estimate of allele frequency for these challenging variants and an expanded analysis of haplotypic and epigenetic variation.

Here, we describe the initial analysis of the first 100 samples sequenced to >30x depth of coverage and a read N50 >50 kbp, which was possible because of the use of HMW DNA isolated directly from cell culture **(Figure 1)**. This resulted in high sensitivity for SV detection using both assembly– and alignment-based approaches, which allowed us to identify an average of 24,543 SVs per sample, similar to prior analysis of other 1KGP samples by the HGSVC and HPRC (Ebert et al. 2021; Liao et al. 2023). Our efforts complement recent work that identified an average of 16,065 SVs in 888 1KGP samples sequenced to lower coverage (15x median depth of coverage) and median read length (6.2 kbp), while demonstrating the advantage of sequencing HMW DNA (Noyvert et al. 2023).

We performed one of the most comprehensive benchmarking analyses to date of SNVs, indels, and SVs using data from the ONT platform. We found that, consistent with prior studies, data generated on the ONT platform has a higher recall and precision than Illumina-based approaches for SNVs in well-characterized genomic regions and performs well for indels, specifically outside of homopolymers (Kolmogorov et al. 2023). Because all data from these first 100 samples were generated on the R9.4.1 pore, we anticipate that improvements in chemistry, such as the use of the R10.4.1 pore, will reduce context-specific errors (such as are seen in homopolymers) and result in improved concordance with truth sets for SNV and indel calls. Because of this expectation, we have transitioned ongoing sequencing to the R10.4.1 pore. SV benchmarking also revealed high F1 scores for three samples for which orthogonal calls were available, highlighting how the R9.4.1 pore is sufficient for this application.

SVs were called using four alignment-based and one assembly-based caller. After merging, a high-confidence SV call set comprising 124,927 SVs was generated that we show can be used for filtering and variant prioritization. Genome-wide evaluation of these high-confidence SVs revealed 378 that were within or encompassed an exon of a medically relevant gene. The low number of SVs intersecting medically relevant genes was reassuring, as we expect there to be selection against these events within coding regions of the genome. Nevertheless, we did identify one SV—an approximately 141-bp insertion in exon 15 of *RPGR*, a gene with an X-linked phenotype—in a 46,XY sample near two similar insertions that have been reported as VUSs in ClinVar. Because the 1KGP samples came from presumably healthy individuals, it could be that this event is associated with a later onset of an associated phenotype or that the insertion is benign. Identification of this insertion in a 1KGP sample is valuable as it may lead to functional studies that clarify the nature of the variant. Analogous to what has been reported for the relatively common occurrence of single nucleotide loss-of-function mutations in otherwise healthy individuals, the presence of an SV in a gene does not necessarily imply the variant is pathogenic (MacArthur et al. 2012). Indeed, early studies of human population samples using SNP microarrays identified extremely rare CNVs > 500 kbp in length among individuals without overt disease (Cooper et al. 2011).

We also ascertained whether LRS might provide better resolution of SVs in key pharmacogenes such as *CYP2D6*. While it was clear that LRS enabled unambiguous characterization of gene deletions (e.g. *CYP2D6*5*) and duplication events (even within the same individual) **(Figure 4B)**, some hybrid rearrangements (e.g. *CYP2B6*29*) remained difficult to fully resolve. Alleles such as this can be used to: 1) evaluate assembly, alignment, and SV calling algorithms; 2) evaluate how differences in read quality contribute to SV resolution; and 3) establish best practices for accurately interrogating SVs in complex pharmacogenes using LRS (Twesigomwe et al. 2023, 2024).

Genome-wide evaluation of select repeat expansions revealed expansions in complex alleles not previously reported and difficult to identify using short-read technology **(Figure 5)**. Repeat expansions in three genes (*RFC1*, *ATXN10*, and *FGF14*) were seen in 9 total samples and included repeat units that have been associated with diseases in the past. Because the individuals recruited to the 1KGP were presumably healthy, these individuals may be at risk of developing symptoms associated with these conditions later in life, or they may be carrying alleles that are nonpathogenic because of sequence interruptions that we did not detect. Alternatively, these expansions may simply be an artifact of the cell culture process and should be considered when these samples are used in other experiments.

Finally, we evaluated patterns of methylation genome-wide and at loci associated with disease. We observed large-scale changes, such as skewed X-inactivation, in over one-third of 46,XX samples and unique changes, such as novel differential methylation that correlates with changes in local gene expression. These changes provide a mechanism by which distinct signals from samples maintained in cell culture can be explained. Furthermore, this finding demonstrates the potential limitations of using immortalized cell lines to infer epigenetic signatures.

Sequencing of 1KGP samples is ongoing and we expect the analysis of a larger number of samples to further refine many of the findings in this study. A majority of the analysis presented here was performed using GRCh38 as a reference due to its widespread use in clinical and research laboratories; work is ongoing to evaluate the impact of the more complete CHM13 T2T genome on variant calling (Nurk et al. 2022). Overall, we anticipate that the dataset provided here will hasten the use of LRS to evaluate individuals with suspected Mendelian conditions for whom a precise molecular diagnosis remains elusive. This work not only provides valuable resources for candidate variant filtering and analysis but also emphasizes the critical need for ongoing investment in technology, software, and database development to fully realize the benefits of LRS. The more comprehensive analysis that can be performed using LRS— such as the identification and resolution of complex SVs, improved phasing, and incorporation of associated methylation information—will allow clinical and research teams to stop focusing on “what’s the next best test” when evaluating an individual with a suspected genetic condition and instead focus on interpreting those variants that were previously difficult to detect or that may involve a novel gene. Together, these efforts will lead to improved clinical outcomes, new gene-phenotype associations, the use of novel therapies, and an end to the diagnostic odyssey for many of the individuals and their families who are living with an unsolved or incompletely understood genetic condition.

## METHODS

### Cell culture

B-Lymphocyte LCLs from the 1000 Genomes Project were purchased from the NHGRI Sample Repository for Human Genetic Research at the Coriell Institute for Medical Research. Cells were shipped overnight at ambient temperature in T25 flasks filled with carbon dioxide-equilibrated transport media. Upon receipt in the lab (day 0), cells were transferred directly to an incubator at 37°C and kept overnight. On day 1, cells and media were transferred to a T75 flask and 20 mL fresh complete RPMI-1640 media (15% fetal bovine serum, 1% penicillin-streptomycin) was added. On day 2, 10 mL of cells were removed for cryopreservation and 10 mL of fresh media was replaced. On day 3, cells were harvested by centrifugation at 300 rcf and resuspended in PBS for counting. An aliquot of 2.5×10å7 cells were pelleted by centrifugation at 300 rcf and resuspended in 3 mL Puregene Cell Lysis Buffer.

### DNA isolation, library preparation, sequencing, and base calling

HMW DNA was isolated from LCLs using either the NEB Monarch HMW DNA Extraction from Cells kit according to manufacturer’s directions or the Puregene DNA Purification from Cultured Cells kit. Puregene extractions were performed using the manufacturer instructions for large volume extractions or a small volume extraction protocol with the following modifications. Small volume cells were resuspended in 1.0 mL of Purgene Cell Lysis Buffer and 6 µL of Qiagen RNase A Solution was added. Both extraction methods were incubated at 37 °C for 40 min. For the small volume method, 333 µL of Puregene Protein Precipitation Solution was added, and both methods were incubated on ice for 10 minutes following homogenization. After DNA precipitation, washes were performed using 666 µL of 70% EtOH. DNA was resuspended with Puregene DNA Hydration Solution or Tris-EDTA buffer and allowed to incubate for 48 hr at 4°C or 24 hr at room temperature before quantifying. Extracted DNA was quantified on a Qubit Fluorometer (Invitrogen) using the dsDNA High Sensitivity Assay. DNA quality for sequencing was assessed using a NanoDrop Spectrophotometer (ThermoFisher) and Agilent Femtopulse following quantification.

After isolation and QC of HMW DNA, libraries for sequencing were prepared using the ligation sequencing kit (SQK-LSK110, ONT), loaded onto a R9.4.1 flow cell, and run on a PromethION 24 sequencer for 24 hr before being washed and reloaded. On average, 3 libraries were loaded and 1.5 flow cells were used per sample. A unique library identifier was used for each wash and reload. Following sequencing, libraries were basecalled using Guppy version 6 (ONT) using the super accurate model with 5mCG modifications or with 5mCG and 5hmCG modifications. Run performance was evaluated using cramino (v0.14.1) (De Coster and Rademakers 2023) **(Table S2)**.

### Alignment and variant calling pipeline

After base calling, FASTQ files with methylation tags were created from unaligned bam files using samtools (v1.17) (Li et al. 2009) and aligned to GRCh38 (GCA_000001405.15_GRCh38_no_alt_analysis_set) with minimap2 (v2.24) (Li 2018). SNVs and indels were called with the Clair3 (v1.0.4) R9 model and bam files were haplotagged using Longphase (Lin et al. 2022; Zheng et al. 2022). Structural variants were called by Sniffles2 (v2.0.7) (Smolka et al. 2024) using default parameters, SVIM (v1.4.2) (Heller and Vingron 2019) using default parameters, and cuteSV (v2.0.3) (Jiang et al. 2020) using the following specific parameters: ––max_cluster_bias_INS 100 ––diff_ratio_merging_INS 0.3 ––max_cluster_bias_DEL 100 ––diff_ratio_merging_DEL 0.3 ––genotype.

### Napu pipeline

The Napu pipeline (Nanopore Analysis Pipeline) was run using default parameters (Kolmogorov et al. 2023). Input files were merged unaligned BAM files for each sample, the GRCh38 reference genome (GCA_000001405.15_GRCh38_no_alt_analysis_set_maskedGRC_exclusions) and corresponding VNTR annotations provided in the Napu GitHub repository.

### Sample validation

For each sample, Clair3 variant calls from the in-house minimap2 alignment pipeline were validated against 1KGP Illumina GATK VCF files (Byrska-Bishop et al. 2022). Specifically, SNVs on chr21 and within the GIAB HG002 high confidence regions were compared using hap.py (Krusche) with Illumina GATK as the truth set and ONT Clair3 calls as the query set. All samples were confirmed to have a SNV precision > 0.98, suggesting sample concordance (nonconcordant samples showed SNV precision <0.50).

### SNV and indel benchmarking

Original sequencing data for 5 benchmarking samples (HG002/NA24385, HG003/NA24149, HG004/NA24143, HG00733, and HG02723) was downloaded and converted from FAST5 to POD5 format using the POD5 Python Package (ONT), then base called with Dorado 0.5.0 (ONT) using the super accurate model with 5mCG modifications. Each sample was downsampled using samtools to match the approximate depth of coverage of the 100 samples presented here and processed with both the in-house alignment pipeline and the Napu pipeline.

SNV and indel calls made using DeepVariant for HG002, HG00733, and HG02723 from the HPRC as well as those for HG002, HG003/NA24149, and HG004/NA24143 from GIAB were obtained (Shafin et al. 2020; Liao et al. 2023). Short-read SNV and indel calls made by GIAB using GATK for HG00733, and HG02723 for chromosomes 1–22 were downloaded and concatenated using bcftools (Danecek et al. 2021; Wagner et al. 2022). All VCFs were preprocessed for ‘FILTER = PASS’ and limited to variants on chromosomes 1–22.

We used hap.py (Krusche) to compare SNV and indel calls from the Napu pipeline to calls from the HPRC and GIAB. The HG002 SNV and indel high-confidence benchmarking bed (HG002_GRCh38_1_22_v4.2.1_benchmark_noinconsistent.bed) was used to limit analysis to high-confidence regions. While sample-specific benchmarking beds are available from GIAB for HG003 and HG004, we chose to use the HG002 bed file for all samples as similar sample-specific filters do not exist for other samples, such as HG00733 or HG02723. In addition, a modified version of HG002_GRCh38_1_22_v4.2.1_benchmark_noinconsistent.bed which excluded homopolymers 4 bp or larger +/– 1 bp (based on GIAB_hg38_Stratifications_v3.3) was used (Dwarshuis et al. 2023).

### SNV and indel comparison between ONT and Illumina

We obtained Illumina GATK SNV/indel VCFs generated for these 100 samples from chromosomes 1–22 (Byrska-Bishop et al. 2022). Variants in this dataset were previously filtered for the following: FILTER=PASS, Genotype missingness < 5%, Pass HWE test (i.e., HWE p-value > 1e-10 in at least 1 of the 5 superpopulations), Mendelian error rate ≤ 5%, and Minor allele count (MAC) ≥ 2. PMDV VCFs for these 100 samples were generated using the Napu pipeline. Clair3 VCFs for the 100 samples were generated using our in-house alignment pipeline. Both PMDV and Clair3 VCFs were preprocessed for ‘FILTER = PASS’ and only variants on full chromosomes 1–22 using bcftools (Danecek et al. 2021). We then ran hap.py in a pairwise fashion (GATK vs. PMDV, GATK vs. Clair3) for each of the 100 samples using GATK as the truth set to calculate precision, recall, and F1 for each sample. For each VCF pair, 3 iterations were run: 1) no masking (all variants included); 2) using the GIAB high-confidence bed file to define confident regions; and 3) using the GIAB high-confidence bed file plus homopolymers 4 bp or greater +/– 1 bp removed.

### *De novo* genome assembly and assembly evaluation

Flye (v2.9.2) was run on all samples using the ‘--nano-hq’ option to obtain haploid assemblies (Kolmogorov et al. 2019). Assembled fasta files were aligned to the GRCh38 reference genome (GCA_000001405.15_GRCh38_no_alt_analysis_set_maskedGRC_exclusions) using minimap2 (v2.24) (Li 2018) with the following parameters: ‘-ax asm20 –B 2 –E 3,1 –O 6,100 –-cs –K 5G’. Starts and ends of aligned contigs were determined using bedtools (v2.3.0) (Quinlan and Hall 2010). The process was repeated for diploid assembly files ‘hapdup_dual_1.fasta’ and ‘hapdup_dual_2.fasta’ from the Napu (Shasta-Hapdup) pipeline (Kolmogorov et al. 2023).

To compute the fraction of the genome covered by assemblies and the contig NG50s, QUAST (v5.2.0) (Mikheenko et al. 2018) was run using the ‘--large’, ‘-x-for-Nx 75’ and ‘–– fragmented’ options on assembled fasta files from Flye and individually on diploid assembly files from the Napu (Shasta-Hapdup) pipeline. Mean QV scores used to evaluate the Flye and Napu (Shasta-Hapdup) assemblies were estimated using yak (r56) (https://github.com/lh3/yak). K-mer hash tables for each sample were generated from matching Illumina short-reads and the QV for the corresponding long-read assembly file was then computed.

For the Napu (Shasta-Hapdup) assemblies, yak (r56) was individually run on the two haplotype-resolved assembly files ‘hapdup_dual_1.fasta’ and ‘hapdup_dual_2.fasta’ and a mean QV for the two files was reported per sample. Matching Illumina datasets for the 100 samples and 2 benchmarking datasets (HG00733 and HG02723) were downloaded from https://www.internationalgenome.org/data-portal/data-collection/30x-grch38. Illumina data for the remaining 3 datasets used for benchmarking (HG002, HG003 and HG004) were downloaded from https://www.nist.gov/programs-projects/genome-bottle.

### Pangenome construction

Contigs of the 100 Shasta-Hapdup assemblies were partitioned by chromosome by mapping them against the GRCh38, CHM13 (v2.0) and HG002 (v1.0.1) human reference genomes using WFMASH (v0.12.6, commit 0b191bb) pangenome aligner (Marco-Sola et al. 2021). On each set of contigs, we used PGGB (v0.5.4, commit 0317e7f) to build chromosome-level unbiased pangenome variation graphs (Garrison et al. 2023). We used ODGI (v0.8.3, commit 861b1c0) to compute similarity matrixes from the pangenome graphs and used R (v4.2.2) to perform the principal component analysis (Guarracino et al. 2022; R Core Team 2021).

### Analysis of assembly contig breakpoints

We characterized the position of assembly breakpoints using precomputed segdup and RepeatMasker positions downloaded from UCSC (Bailey et al. 2002; Kent et al. 2002). The position of assembly breaks were categorized as “Satellite” (only satellite repeats), “SegDup+Satellite” (segdups and satellite repeats), “SegDup” (only segdups) or “Neither” (outside segdups and satellite repeat regions).

### Gene assembly stats

A list of medically relevant genes was downloaded from OMIM (https://www.omim.org/). Gaps in either the Flye or Napu (Shasta-Hapdup) assemblies were defined using the GenomicRanges R package (Lawrence et al. 2013). Gaps were defined as regions in the genome where no contig was mapped. After filtering for regions representing OMIM genes, mapping_fraction for each OMIM gene was calculated as follows:

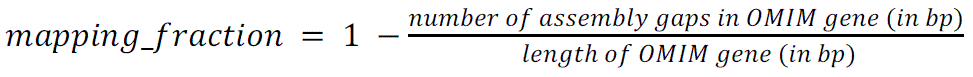

Data from the Shasta-Hapdup and Flye assemblies were filtered for OMIM genes with mapping_fraction less than 1 or for genes where more than one contig spanned the gene indicating an incomplete or broken assembly. If at least one of two Shasta-Hapdup assemblies had a mapping_fraction less than 1 or multiple contigs spanning a gene it was counted as an incomplete assembly for that gene.

### Calculation of median contig sizes by superpopulation

Contig sizes for each sample were calculated by taking the difference between the start and end position in the BED file. For each sample, the median contig size for contigs larger than 1 Mb was calculated separately for the Shasta-Hapdup and Flye assemblies.

### SV analysis, merging, and benchmarking

All SV VCFs were preprocessed to standardize SV type annotations and to include SVs that passed filtering (FILTER = PASS), were ≥ 50 bp in length (if SV length was not reported for the SV type, the variant was kept), and were located on full-length chromosomes (chromosome 1-22, X, Y, and M) using bcftools commands specific to each caller. After preprocessing, the number of SVs (INS, DEL, INV, DUP, and BND) were counted per sample and individual VCFs for each call set (minimap2 pipeline: Sniffles2, cuteSV, and SVIM; Napu pipeline: Sniffles2 and hapdiff) were concatenated and parsed by SV type. After concatenation, the length of all INS or DEL events for all 100 samples were calculated. We then calculated novel SVs per sample (for each caller) by adding samples in reverse alphanumeric order by reverse alphabetical ancestry and counting the number of new SVs included, with samples of AFR ancestry added last. For each iteration, SVs within a call set were merged with Jasmine (--allow_intrasample –-output_genotypes –-ignore_strand –-dup_to_ins –-centroid_merging) and then the resultant output VCF was parsed by SV type.

To analyze overlapping SVs, calls from all 5 callers were merged using Jasmine. The Jasmine output for each sample was parsed using the “SUPP_VEC’’ field indicating which of the 5 callers supported each output variant. The minimum support flag was included to retain only SVs that were called by at least a minimum number of the 5 callers. A confident call set was defined by variants called (per sample) by hapdiff and at least 2 unique alignment-based callers (i.e., Sniffles2 and CuteSV from our in-house pipeline, or Sniffles2 from the Napu pipeline and SVIM from the in-house pipeline), but not if the call was supported only by Sniffles2 calls from both the Napu and in-house pipeline.

To perform SV benchmarking, downsampled ONT data from HG002/GM24385, HG00733, and HG02723 were processed using the Napu pipeline and SV calls from Sniffles2 and hapdiff were preprocessed as above. We obtained Sniffles2 calls from the HPRC and preprocessed them as above. Each ONT sample (for both Sniffles2 and hapdiff) was benchmarked to the HPRC (truth) calls using Truvari (v4.1.0) (English et al. 2022) with the following options: –-pick multi –-chunksize 2000 –r 2000 –-dup-to-ins –-pctseq 0. A GRCh19 to GRCh38 liftover of the GIAB HG002 SV Tier1 benchmarking bed was used to define regions for inclusion.

We benchmarked HG002 SV calls against the draft GIAB T2TQ100 HG002 GRCh38 SV benchmark (https://ftp-trace.ncbi.nlm.nih.gov/ReferenceSamples/giab/data/AshkenazimTrio/analysis/NIST_HG002_DraftBenchmark_defrabbV0.015-20240215/). The draft benchmark was generated using v0.015 of DeFrABB (https://github.com/usnistgov/giab-defrabb). Briefly, DeFrABB uses the variant calls and diploid assembled regions identified by dipcall (Li et al. 2018) from assembly–assembly alignments. V1.01 of the HG002 Q100 assembly (https://github.com/marbl/hg002) and a modified version of GRCh38 (https://ftp-trace.ncbi.nlm.nih.gov/ReferenceSamples/giab/release/references/GRCh38/GRCh38_GIABv3_no_alt_analysis_set_maskedGRC_decoys_MAP2K3_KMT2C_KCNJ18.fasta.gz) were used as inputs. We started with regions with a 1:1 alignment between each assembled haplotype and the reference (except the X and Y chromosomes) and then excluded gaps in the assembly and their flanking sequences as well as any large repeats (satellites, tandem repeats >10 kb, and segdups) that have a break in the assembly to reference alignment on either haplotype. The SV call sets were compared to the draft GIAB SV benchmark set using truvari refine (v4.2.2.dev0+1cd03b2) (English et al. 2022). First, truvari bench was run using the options (–– sizemin 50 –-pick ac), then truvari refine was run using the truvari bench results as input with options (--recount –-use-region-coords –-use-original-vcfs –-align mafft –-use-original –-reference ref/GCA_000001405.15_GRCh38_no_alt_analysis_set.fasta.gz –-regions candidate.refine.bed), where candidate.refine.bed is a bed file with target regions for refinement generated by truvari bench.

### Filtering and prioritization of SVs

Sniffles2 SV calls made using our in-house pipeline from cases sequenced on the ONT platform that were known to have a disease-causing SV were preprocessed as above and merged using Jasmine (parameters above) along with Sniffles2 SV calls from the Napu pipeline from the 100 samples. The Jasmine output was parsed for SVs present only in the case sample and annotated with the following: 1) does the variant intersect a protein-coding gene as defined by GENCODE release 45 (Frankish et al. 2021); 2) is the gene associated with a phenotype in OMIM; 3) does the SV intersect an exon of a canonical protein-coding exon defined by GENCODE release 45; and 4) does the SV intersect a complex genomic feature such as a centromere, segdup, or gap in GRCh38.

### eQTL analysis

Following the SV-eQTL method presented by Kirsche et al. (2023), we performed SV-eQTL calling within the 65 samples that have both long-read ONT DNA sequencing data and short-read RNA sequencing data from MAGE **(Figure S11)**. Briefly, this approach fits a linear model between the SV genotypes (0=homozygous ref, 1=heterozygous alt, 2=homozygous alt) with the normalized MAGE expression measurements and sex information using the OLS module in python. To calculate gene-level eQTL p-values, we applied Bonferroni correction to the minimal eQTL p-value for each gene, adjusting by the number of eQTLs associated with that gene. Then, we further adjusted these gene-level *p*-values for multiple testing using the Benjamini-Hochberg method with an FDR rate below 20%. The higher FDR rate was needed due to the limited number of samples, but we noted that SV-eQTL pairs common to this analysis and the Kirsche *et al*. analysis typically had more significant *p*-values reported in Kirsche *et al*. (2023).

### Identification of SVs in medically relevant genes

The 5 SV callers (in-house pipeline: Sniffles2, cuteSV, and SVIM; Napu pipeline: Sniffles2 and hapdiff) for each of the 100 samples were merged using Jasmine and parsed for confident calls for each sample. The confident calls for each sample were then merged using Jasmine. The output from the intersample Jasmine merge was intersected with a custom-built bed file of genomic coordinates of medically relevant exons. An ideogram of the genomic locations of each of the “custom confident” SVs that intersects a medically relevant exon was plotted in R using KaryoploteR (Gel and Serra 2017). All genomic boundaries were defined by GENCODE release 45 using the following definitions: 1) protein-coding genic regions are genes with gene_type and protein_coding, 2) UTRs are defined as Ensembl_canonical and in protein_coding genes, 3) intronic regions are defined as protein_coding genic regions outside of Ensembl_canonical protein_coding exons, and 4) medically relevant exons are defined as Ensembl_canonical exons in OMIM genes.

### Repeat masking of the first 100 assemblies

Each assembled haplotype from the Napu pipeline (Shasta-Hapdup) was masked using RepeatMasker (http://www.repeatmasker.org/) with the rmblast search engine (‘-e rmblast’) and the built-in human transposable element database (‘-species human’). HG002 and HG005 RepeatMasker output was downloaded from the HPRC. CHM13 T2T (hs1) RepeatMasker output was downloaded from UCSC (Nurk et al. 2022). The transposable element fraction data of the Shasta-Hapdup assemblies, HG002, and HG005 were extracted from the standard ‘.tbl’ output of Repeat Masker. The CHM13 T2T genome transposable element fraction data was adapted from Hoyt *et al*. (Hoyt et al. 2022). The number of L1HS in each genome was counted by extracting lines matching ‘L1HS’ from the RepeatMasker standard ‘.out’ output. The number of full-length L1HS elements was counted by extracting the lines from the RepeatMasker output file by searching for ‘L1HS’ and filtering for the distance between the starting and ending coordinate of the L1HS is ≥ 6,000 bp.

### Tandem Repeat Genotyping

Repeats were genotyped using vamos v1.2.6 (Ren et al. 2023) and a list of genome-wide simple repeat loci was made by intersecting the original motif set from vamos (https://zenodo.org/records/8357361) with the GIAB Tier 1 regions and the UCSC simple repeat regions (Benson 1999; Zook et al. 2020). This was combined with the position and motifs for 66 disease-associated loci derived from the STRchive (https://github.com/hdashnow/STRchive). This resulted in 562,005 total loci. Nonreference alleles for *ATXN10* not included in the STRchive were obtained from (Morato Torres et al. 2022). A bed file with the coordinates and metadata for each locus is provided **(Table S9)**. The haplotype-resolved hapdiff assemblies from the NAPU pipeline were used to genotype the repeat and evaluate sequence context. Since haplotypes are assigned arbitrarily, all “mat” assemblies were assigned haplotype 1, and all “pat” assemblies were assigned haplotype 2. Because the haplotype-resolved assemblies can fill in missing sequences, only the haplotype 1 value for X chromosome loci was considered for 46,XY individuals. The output from vamos is a per-sample per-haplotype VCF of the genotype of each allele. The VCFs were combined with the vamos combine_vcf.py script (https://github.com/ChaissonLab/vamos/blob/master/snakefile/pyscript/combine_vcf.py). Each haplotype is a separate sample in the combined VCF in **File S2**.

ExpansionHunter v5.0.0 was run to genotype STRs on all samples using the ExpansionHunter genome-wide STR catalog v1.0.0 (https://github.com/Illumina/RepeatCatalogs/tree/master) containing 174,293 STR loci using the streaming analysis mode (`-m streaming`) as recommended for large variant catalogs (Dolzhenko et al. 2019). TRTools v5.0.2 mergeSTR method was then used to generate a multi-sample VCF file (Mousavi et al. 2021).

### Methylation analysis

Haplotype-resolved, whole-genome methylation pileup files were generated using modkit v0.1.11 (ONT) using the flags –-cpg –-ref –-ignore h –-combine-strands –-partition-tag HP. Strands were combined for one read count value using the ‘--combine_strands’ option for each CpG corresponding to CpGs in the GRCh38 reference (GCA_000001405.15_GRCh38_no_alt_analysis_set_maskedGRC_exclusions_v2.fasta). Pileup files were generated from the PMDV haplotagged bam file from the Napu pipeline. Haplotype-resolved pileup files were subset for CpGs within defined X chromosome CpG islands using bedtools v2.30.0 (Quinlan and Hall 2010). For X chromosome analysis, the average fraction of methylated reads was calculated for each CpG island on the X chromosome. Visualizing patterns at this level, we selected 20 samples that exhibited a skewed distribution and filtered CpG islands found in the majority of samples (≥16) where the average fraction methylated +/– the standard deviation did not overlap across haplotypes. This resulted in 397 informative CpG islands, which were plotted for each sample. Samples were filtered to include only CpG islands within that list where the mean coverage was ≥ 5 reads. The X-chromosome inactivation pattern was then predicted by taking the median difference between the fraction methylated between haplotypes at each informative CpG island.

We subsetted CpG islands at the *H19* (chr11:1997582-2003510) and *KCNQ1OT1* (chr11:2698718-2701029) loci and *SNURF*-*SNRPN* (chr15:24954857-24956829) (Akbari et al. 2022). The average fraction of reads methylated was calculated per sample and per haplotype.

Unique DMRs were identified with the Methylation Operation Wizard (MeOW) using a leave-one-out analysis (Zalusky and Miller 2024). Whole-genome methylation frequencies were quantified for CpG dinucleotide positions within previously defined CpG islands. To simplify analysis, 24 datasets that included 5hmCG methylation in addition to 5mCG methylation were excluded. The probability of methylation across all reads for each position was averaged and rescaled to a [0,1] interval. The probability of differential methylation was determined using the bootstrapped beta regression test option in MeOW and filtered using both the benjamin-hochberg corrected *p*-values and a Cohen’s *d* cutoff of 1.5. Comparisons were restricted to CpG islands with 50 or more CpG positions containing non-N bases in all 76 sample datasets, comprising 20,836 regions. Methylation for the *SLC29A3* DMR was visualized with modbamtools v0.4.8 (Razaghi et al. 2022).

Principal component analysis of methylation data was performed by taking the average fraction methylated of each CpG island calculated from the per-CpG dataset. PCA was performed comparing 75 samples using prcomp from the stats package in R 4.3.1.

To evaluate associated changes in gene expression, DMRs were subset to those occurring in African Functional Genomics Resource (AFGR) samples that had RNA-sequencing data available for analysis (n=15 individuals, n=66 DMRs) (DeGorter et al. 2023). The coordinates of each DMR were expanded by 10 kbp and intersected with protein-coding genes based on Gencode v35 gene models using plyranges left_overlap_join (Frankish et al. 2021). Expression *z*-scores were pulled for the 85 genes occurring near DMRs based on that criteria. To test enrichment, a varying expression absolute *z*-score threshold (0.1, 0.5, 1, 1.5, …, 4) was set to determine expression outliers, then the log odds ratio estimates and standard errors (SE) were calculated from a logistic regression across all sample-gene pairs, (model: Expression_Outlier ∼ Sample_has_DMR + ε). Log odds ratios were plotted with 95% CI, estimate +/– 1.96*SE.

## DATA ACCESS

Data for all samples sequenced as part of the 1000 Genomes Project ONT Sequencing Consortium are publicly available at https://s3.amazonaws.com/1000g-ont/index.html. Data from the 100 samples reported here, as well as summary analysis data, are available at https://s3.amazonaws.com/1000g-ont/index.html?prefix=FIRST_100_FREEZE/. Data and code related to pangenome analyses are available at https://github.com/AndreaGuarracino/1000G-ONT-F100-PGGB.

## ACKNOWLEDGMENTS

SBG is supported by NIH grant 5T32HG000035-29; WDC is a recipient of a postdoctoral fellowship from FWO [12ASR24N]; EG and AG are supported by NIH grants R01HG013017 and U01DA057530 and NSF grant 2118744; SG is supported by NIH grant 5R50CA243890; TDJ is supported by NIH grant T32HG000044; MK is supported by Intramural NIH funding; SBM, TDJ, and ER is supported by NIH Grant U01HG011762; MCS is supported by NIH grants U24HG010263, R03CA272952, and U01CA253481 and the Lustgarten Foundation grant 90101412; FJS is supported by NIH grants 1U01HG011758-01, 1UG3NS132105-01, and U01AG058589; AAS is supported by an NSF postdoctoral research fellowship in biology [NSF 22-623]; RNM and LY are supported by NIH grants 5R35GM142733-03 and 5R21AI174130-02; EEE is supported by NIH grant HG010169 and EEE is an investigator of the Howard Hughes Medical Institute; DEM is supported by the NIH Director’s Early Independence Award DP5OD033357. The GREGoR Consortium is funded by the National Human Genome Research Institute of the National Institutes of Health, through the following grants: U01HG011758, U01HG011755, U01HG011745, U01HG011762, U01HG011744, and U24HG011746. The content is solely the responsibility of the authors and does not necessarily represent the official views of the National Institutes of Health. No human subjects, live vertebrates, or higher invertebrate research was undertaken as part of this manuscript. Certain commercial equipment, instruments, or materials are identified to adequately specify experimental conditions or reported results. Such identification does not imply recommendation or endorsement by the National Institute of Standards and Technology, nor does it imply that the equipment, instruments, or materials identified are necessarily the best available for the purpose.

## SUPPLEMENTAL MATERIAL

### Supplemental Figures

- **Figure S1.** Precision, recall, and F1 scores for SNV and indel calls for ONT and Illumina small variant calls using PacBio assemblies as the truth set
- **Figure S2.** Precision, recall, and F1 scores for SNV and indel calls for ONT small variant calls from both the Napu (PMDV) and alignment (Clair3) pipelines using Illumina data as the truth set.
- **Figure S3.** Percent of genome assembled by Flye and Shasta-Hapdup
- **Figure S4.** IGV screenshots of chromosome 7 assemblies
- **Figure S5.** Pangenome principal component analysis
- **Figure S6.** Evaluation of L1HS elements in the first 100 assemblies.
- **Figure S7.** Evaluation of per-sample insertions and deletions by type, chromosome, and SV caller
- **Figure S8.** Distribution of SV events genome-wide, all populations
- **Figure S9.** Concordance of SVs by caller and evaluation of joint SV calls
- **Figure S10.** Example of false-positive SVs called only by hapdiff
- **Figure S11.** Evaluating the functional impact of structural variants
- **Figure S12.** Evaluation of SVs in exons of medically relevant genes
- **Figure S13.** IGV view of a previously described deletion that includes *HBM*, *HBA2*, *HBA1*, *HBQ1*
- **Figure S14.** Evaluation of X-linked SVs in 46,XY individuals
- **Figure S15.** Phased IGV screenshot depicting long-read alignments involving two pharmacogene hybrid star alleles
- **Figure S16.** Repeat expansion plots for 66 disease-associated loci.
- **Figure S17.** IGV screenshots of repeat expansions in RFC1 observed in 5 samples.
- **Figure S18.** IGV screenshots of *ATXN10* repeat expansions.
- **Figure S19.** IGV screenshots of repeat expansion within *FGF14*
- **Figure S20.** Principal component analysis using methylation
- **Figure S21.** Evaluating methylation differences at *SNURF-SNRPN*
- **Figure S22.** Output from MeOW showing DMRs within 3 samples that have 10 or more DMRs
- **Figure S23.** Unique DMRs are associated with nearby changes in gene expression

### Supplemental Tables

- **Table S1.** Demographics
- **Table S2.** Sequencing statistics (Cramino)
- **Table S3.** Downsampled benchmarking sample statistics
- **Table S4.** Chromosome-wise contig break percentages
- **Table S5.** Masked TEs
- **Table S6.** SV Benchmarking
- **Table S7.** SVs in OMIM exons
- **Table S8.** STRchive disease metadata
- **Table S9.** X chromosome inactivation status
- **Table S10.** MeOW results d1.5_p01 with AFGR expression data

### Supplemental Files

- **File S1**. OMIM gene assembly statistics from Flye and Shasta-Hapdup across 100 samples, including proximity to a segmental duplication.
- **File S2**. Combined VCF of vamos output for each sample.

## REFERENCES

1. 1000 Genomes Project Consortium, Auton A, Brooks LD, Durbin RM, Garrison EP, Kang HM, Korbel JO, Marchini JL, McCarthy S, McVean GA, et al. 2015. A global reference for human genetic variation. Nature 526: 68–74.

2. Akbari V, Garant J-M, O’Neill K, Pandoh P, Moore R, Marra MA, Hirst M, Jones SJM. 2022. Genome-wide detection of imprinted differentially methylated regions using nanopore sequencing. Elife 11: e77898.

3. Akçimen F, Ross JP, Bourassa CV, Liao C, Rochefort D, Gama MTD, Dicaire M-J, Barsottini OG, Brais B, Pedroso JL, et al. 2019. Investigation of the RFC1 Repeat Expansion in a Canadian and a Brazilian Ataxia Cohort: Identification of Novel Conformations. Front Genet 10: 1219.

4. AlAbdi L, Shamseldin HE, Khouj E, Helaby R, Aljamal B, Alqahtani M, Almulhim A, Hamid H, Hashem MO, Abdulwahab F, et al. 2023. Beyond the exome: utility of long-read whole genome sequencing in exome-negative autosomal recessive diseases. Genome Medicine 15: 114.

5. Alonso I, Jardim LB, Artigalas O, Saraiva-Pereira ML, Matsuura T, Ashizawa T, Sequeiros J, Silveira I. 2006. Reduced penetrance of intermediate size alleles in spinocerebellar ataxia type 10. Neurology 66: 1602–1604.

6. Audano PA, Sulovari A, Graves-Lindsay TA, Cantsilieris S, Sorensen M, Welch AE, Dougherty ML, Nelson BJ, Shah A, Dutcher SK, et al. 2019. Characterizing the Major Structural Variant Alleles of the Human Genome. Cell 176: 663–675.e19.

7. Bailey JA, Gu Z, Clark RA, Reinert K, Samonte RV, Schwartz S, Adams MD, Myers EW, Li PW, Eichler EE. 2002. Recent segmental duplications in the human genome. Science 297: 1003–1007.

8. Beecroft SJ, Cortese A, Sullivan R, Yau WY, Dyer Z, Wu TY, Mulroy E, Pelosi L, Rodrigues M, Taylor R, et al. 2020. A Māori specific RFC1 pathogenic repeat configuration in CANVAS, likely due to a founder allele. Brain 143: 2673–2680.

9. Benson G. 1999. Tandem repeats finder: a program to analyze DNA sequences. Nucleic Acids Res 27: 573–580.

10. Byrska-Bishop M, Evani US, Zhao X, Basile AO, Abel HJ, Regier AA, Corvelo A, Clarke WE, Musunuri R, Nagulapalli K, et al. 2022. High-coverage whole-genome sequencing of the expanded 1000 Genomes Project cohort including 602 trios. Cell 185: 3426–3440.e19.

11. Cameron DL, Di Stefano L, Papenfuss AT. 2019. Comprehensive evaluation and characterisation of short read general-purpose structural variant calling software. Nat Commun 10: 3240.

12. Chaisson MJP, Sanders AD, Zhao X, Malhotra A, Porubsky D, Rausch T, Gardner EJ, Rodriguez OL, Guo L, Collins RL, et al. 2019. Multi-platform discovery of haplotype-resolved structural variation in human genomes. Nat Commun 10: 1784.

13. Chaisson MJP, Sulovari A, Valdmanis PN, Miller DE, Eichler EE. 2023. Advances in the discovery and analyses of human tandem repeats. Emerging Topics in Life Sciences ETLS20230074.

14. Chen R, Wang X, Dai Z, Wang Z, Wu W, Hu Z, Zhang X, Liu Z, Zhang H, Cheng Q. 2021. TNFSF13 Is a Novel Onco-Inflammatory Marker and Correlates With Immune Infiltration in Gliomas. Front Immunol 12: 713757.

15. Cohen ASA, Farrow EG, Abdelmoity AT, Alaimo JT, Amudhavalli SM, Anderson JT, Bansal L, Bartik L, Baybayan P, Belden B, et al. 2022. Genomic answers for children: Dynamic analyses of >1000 pediatric rare disease genomes. Genet Med 24: 1336–1348.

16. Cooper GM, Coe BP, Girirajan S, Rosenfeld JA, Vu TH, Baker C, Williams C, Stalker H, Hamid R, Hannig V, et al. 2011. A copy number variation morbidity map of developmental delay. Nat Genet 43: 838–846.

17. Cortese A, Reilly MM, Houlden H. 1993. RFC1 CANVAS / Spectrum Disorder. In GeneReviews® (eds. M.P. Adam, J. Feldman, G.M. Mirzaa, R.A. Pagon, S.E. Wallace, L.J. Bean, K.W. Gripp, and A. Amemiya), University of Washington, Seattle, Seattle (WA) http://www.ncbi.nlm.nih.gov/books/NBK564656/ (Accessed February 5, 2024).

18. Cortese A, Simone R, Sullivan R, Vandrovcova J, Tariq H, Yau WY, Humphrey J, Jaunmuktane Z, Sivakumar P, Polke J, et al. 2019. Biallelic expansion of an intronic repeat in RFC1 is a common cause of late-onset ataxia. Nat Genet 51: 649–658.

19. Dagli AI, Mathews J, Williams CA. 1993. Angelman Syndrome. In GeneReviews® (eds. M.P. Adam, G.M. Mirzaa, R.A. Pagon, S.E. Wallace, L.J. Bean, K.W. Gripp, and A. Amemiya), University of Washington, Seattle, Seattle (WA) http://www.ncbi.nlm.nih.gov/books/NBK1144/ (Accessed October 10, 2023).

20. Damaraju N, Miller AL, Miller DE. 2024. Long-Read DNA and RNA Sequencing to Streamline Clinical Genetic Testing and Reduce Barriers to Comprehensive Genetic Testing. The Journal of Applied Laboratory Medicine 9: 138–150.

21. Danecek P, Bonfield JK, Liddle J, Marshall J, Ohan V, Pollard MO, Whitwham A, Keane T, McCarthy SA, Davies RM, et al. 2021. Twelve years of SAMtools and BCFtools. Gigascience 10: giab008.

22. Danforth DN. 2016. Genomic Changes in Normal Breast Tissue in Women at Normal Risk or at High Risk for Breast Cancer. Breast Cancer (Auckl) 10: 109–146.

23. De Coster W, Rademakers R. 2023. NanoPack2: population-scale evaluation of long-read sequencing data. Bioinformatics 39: btad311.

24. DeGorter MK, Goddard PC, Karakoc E, Kundu S, Yan SM, Nachun D, Abell N, Aguirre M, Carstensen T, Chen Z, et al. 2023. Transcriptomics and chromatin accessibility in multiple African population samples. 2023.11.04.564839. https://www.biorxiv.org/content/10.1101/2023.11.04.564839v1 (Accessed February 26, 2024).

25. Depienne C, Mandel J-L. 2021. 30 years of repeat expansion disorders: What have we learned and what are the remaining challenges? The American Journal of Human Genetics 108: 764–785.

26. Dolzhenko E, Deshpande V, Schlesinger F, Krusche P, Petrovski R, Chen S, Emig-Agius D, Gross A, Narzisi G, Bowman B, et al. 2019. ExpansionHunter: a sequence-graph-based tool to analyze variation in short tandem repeat regions. Bioinformatics 35: 4754–4756.

27. Dolzhenko E, English A, Dashnow H, Brandine GDS, Mokveld T, Rowell WJ, Karniski C, Kronenberg Z, Danzi MC, Cheung WA, et al. 2023. Resolving the unsolved: Comprehensive assessment of tandem repeats at scale. 2023.05.12.540470. https://www.biorxiv.org/content/10.1101/2023.05.12.540470v1 (Accessed May 15, 2023).

28. Driscoll DJ, Miller JL, Cassidy SB. 1993. Prader-Willi Syndrome. In GeneReviews® (eds. M.P. Adam, J. Feldman, G.M. Mirzaa, R.A. Pagon, S.E. Wallace, L.J. Bean, K.W. Gripp, and A. Amemiya), University of Washington, Seattle, Seattle (WA) http://www.ncbi.nlm.nih.gov/books/NBK1330/ (Accessed February 7, 2024).

29. Dwarshuis N, Kalra D, McDaniel J, Sanio P, Jerez PA, Jadhav B, Huang W (Eddy), Mondal R, Busby B, Olson ND, et al. 2023. The GIAB genomic stratifications resource for human reference genomes. 2023.10.27.563846. https://www.biorxiv.org/content/10.1101/2023.10.27.563846v1 (Accessed February 22, 2024).

30. Ebert P, Audano PA, Zhu Q, Rodriguez-Martin B, Porubsky D, Bonder MJ, Sulovari A, Ebler J, Zhou W, Serra Mari R, et al. 2021. Haplotype-resolved diverse human genomes and integrated analysis of structural variation. Science 372: eabf7117.

31. Eichler EE. 2019. Genetic Variation, Comparative Genomics, and the Diagnosis of Disease. N Engl J Med 381: 64–74.

32. English AC, Menon VK, Gibbs RA, Metcalf GA, Sedlazeck FJ. 2022. Truvari: refined structural variant comparison preserves allelic diversity. Genome Biology 23: 271.

33. Fahim AT, Daiger SP, Weleber RG. 1993. Nonsyndromic Retinitis Pigmentosa Overview. In GeneReviews® (eds. M.P. Adam, J. Feldman, G.M. Mirzaa, R.A. Pagon, S.E. Wallace, L.J. Bean, K.W. Gripp, and A. Amemiya), University of Washington, Seattle, Seattle (WA) http://www.ncbi.nlm.nih.gov/books/NBK1417/ (Accessed February 27, 2024).

34. Fan Y, Zhang S, Yang J, Mao C-Y, Yang Z-H, Hu Z-W, Wang Y-L, Liu Y-T, Liu H, Yuan Y-P, et al. 2020. No biallelic intronic AAGGG repeat expansion in RFC1 was found in patients with late-onset ataxia and MSA. Parkinsonism Relat Disord 73: 1–2.

35. Farashi S, Harteveld CL. 2018. Molecular basis of α-thalassemia. *Blood Cells*, Molecules, and Diseases 70: 43–53.

36. Firth HV, Richards SM, Bevan AP, Clayton S, Corpas M, Rajan D, Vooren SV, Moreau Y, Pettett RM, Carter NP. 2009. DECIPHER: Database of Chromosomal Imbalance and Phenotype in Humans Using Ensembl Resources. The American Journal of Human Genetics 84: 524–533.

37. Frankish A, Diekhans M, Jungreis I, Lagarde J, Loveland JE, Mudge JM, Sisu C, Wright JC, Armstrong J, Barnes I, et al. 2021. GENCODE 2021. Nucleic Acids Research 49: D916– D923.

38. Garrison E, Guarracino A, Heumos S, Villani F, Bao Z, Tattini L, Hagmann J, Vorbrugg S, Marco-Sola S, Kubica C, et al. 2023. Building pangenome graphs. 2023.04.05.535718. https://www.biorxiv.org/content/10.1101/2023.04.05.535718v1 (Accessed March 3, 2024).

39. Gel B, Serra E. 2017. karyoploteR: an R/Bioconductor package to plot customizable genomes displaying arbitrary data. Bioinformatics 33: 3088–3090.

40. Guarracino A, Heumos S, Nahnsen S, Prins P, Garrison E. 2022. ODGI: understanding pangenome graphs. Bioinformatics 38: 3319–3326.

41. Hannan AJ. 2018. Tandem repeats mediating genetic plasticity in health and disease. Nat Rev Genet 19: 286–298.

42. Harvey WT, Ebert P, Ebler J, Audano PA, Munson KM, Hoekzema K, Porubsky D, Beck CR, Marschall T, Garimella K, et al. 2023. Whole-genome long-read sequencing downsampling and its effect on variant-calling precision and recall. Genome Res 33: 2029–2040.

43. Heller D, Vingron M. 2019. SVIM: structural variant identification using mapped long reads. Bioinformatics 35: 2907–2915.

44. Hiatt SM, Lawlor JMJ, Handley LH, Ramaker RC, Rogers BB, Partridge EC, Boston LB, Williams M, Plott CB, Jenkins J, et al. 2021. Long-read genome sequencing for the molecular diagnosis of neurodevelopmental disorders. HGG Adv 2: 100023.

45. Hijikata A, Suyama M, Kikugawa S, Matoba R, Naruto T, Enomoto Y, Kurosawa K, Harada N, Yanagi K, Kaname T, et al. 2024. Exome-wide benchmark of difficult-to-sequence regions using short-read next-generation DNA sequencing. Nucleic Acids Research 52: 114–124.

46. Hoyt SJ, Storer JM, Hartley GA, Grady PGS, Gershman A, de Lima LG, Limouse C, Halabian R, Wojenski L, Rodriguez M, et al. 2022. From telomere to telomere: The transcriptional and epigenetic state of human repeat elements. Science 376: eabk3112.

47. Huisman TH, Wrightstone RN, Wilson JB, Schroeder WA, Kendall AG. 1972. Hemoglobin Kenya, the product of fusion of amd polypeptide chains. Arch Biochem Biophys 153: 850–853.

48. International HapMap Consortium. 2005. A haplotype map of the human genome. Nature 437: 1299–1320.

49. Jiang T, Liu Y, Jiang Y, Li J, Gao Y, Cui Z, Liu Y, Liu B, Wang Y. 2020. Long-read-based human genomic structural variation detection with cuteSV. Genome Biology 21: 189.

50. Kent WJ, Sugnet CW, Furey TS, Roskin KM, Pringle TH, Zahler AM, Haussler and D. 2002. The Human Genome Browser at UCSC. Genome Res 12: 996–1006.

51. Kirsche M, Prabhu G, Sherman R, Ni B, Battle A, Aganezov S, Schatz MC. 2023. Jasmine and Iris: population-scale structural variant comparison and analysis. Nat Methods 20: 408– 417.

52. Koenig Z, Yohannes MT, Nkambule LL, Goodrich JK, Kim HA, Zhao X, Wilson MW, Tiao G, Hao SP, Sahakian N, et al. 2023. A harmonized public resource of deeply sequenced diverse human genomes. 2023.01.23.525248. https://www.biorxiv.org/content/10.1101/2023.01.23.525248v3 (Accessed September 4, 2023).

53. Kolmogorov M, Billingsley KJ, Mastoras M, Meredith M, Monlong J, Lorig-Roach R, Asri M, Alvarez Jerez P, Malik L, Dewan R, et al. 2023. Scalable Nanopore sequencing of human genomes provides a comprehensive view of haplotype-resolved variation and methylation. Nat Methods 20: 1483–1492.

54. Kolmogorov M, Yuan J, Lin Y, Pevzner PA. 2019. Assembly of long, error-prone reads using repeat graphs. Nat Biotechnol 37: 540–546.

55. Krusche P. Haplotype comparison tools / hap.py. http://github.com/illumina/hap.py.

56. Lawrence M, Huber W, Pagès H, Aboyoun P, Carlson M, Gentleman R, Morgan MT, Carey VJ. 2013. Software for Computing and Annotating Genomic Ranges. PLOS Computational Biology 9: e1003118.

57. Lee S, Wheeler MM, Patterson K, McGee S, Dalton R, Woodahl EL, Gaedigk A, Thummel KE, Nickerson DA. 2019. Stargazer: a software tool for calling star alleles from next-generation sequencing data using CYP2D6 as a model. Genet Med 21: 361–372.

58. Li H. 2018. Minimap2: pairwise alignment for nucleotide sequences. Bioinformatics 34: 3094– 3100.

59. Li H, Bloom JM, Farjoun Y, Fleharty M, Gauthier L, Neale B, MacArthur D. 2018. A synthetic-diploid benchmark for accurate variant-calling evaluation. Nat Methods 15: 595–597.

60. Li H, Handsaker B, Wysoker A, Fennell T, Ruan J, Homer N, Marth G, Abecasis G, Durbin R, 1000 Genome Project Data Processing Subgroup. 2009. The Sequence Alignment/Map format and SAMtools. Bioinformatics 25: 2078–2079.

61. Li W, Lin L, Malhotra R, Yang L, Acharya R, Poss M. 2019. A computational framework to assess genome-wide distribution of polymorphic human endogenous retrovirus-K In human populations. PLoS Comput Biol 15: e1006564.

62. Liao W-W, Asri M, Ebler J, Doerr D, Haukness M, Hickey G, Lu S, Lucas JK, Monlong J, Abel HJ, et al. 2023. A draft human pangenome reference. Nature 617: 312–324.

63. Lin J-H, Chen L-C, Yu S-C, Huang Y-T. 2022. LongPhase: an ultra-fast chromosome-scale phasing algorithm for small and large variants. Bioinformatics 38: 1816–1822.

64. Logsdon GA, Vollger MR, Eichler EE. 2020. Long-read human genome sequencing and its applications. Nat Rev Genet 21: 597–614.

65. MacArthur DG, Balasubramanian S, Frankish A, Huang N, Morris J, Walter K, Jostins L, Habegger L, Pickrell JK, Montgomery SB, et al. 2012. A Systematic Survey of Loss-of-Function Variants in Human Protein-Coding Genes. Science 335: 823–828.

66. Marco-Sola S, Moure JC, Moreto M, Espinosa A. 2021. Fast gap-affine pairwise alignment using the wavefront algorithm. Bioinformatics 37: 456–463.

67. Matsuura T, Ashizawa T. 1993. Spinocerebellar Ataxia Type 10. In GeneReviews® (eds. M.P. Adam, J. Feldman, G.M. Mirzaa, R.A. Pagon, S.E. Wallace, L.J. Bean, K.W. Gripp, and A. Amemiya), University of Washington, Seattle, Seattle (WA) http://www.ncbi.nlm.nih.gov/books/NBK1175/ (Accessed February 5, 2024).

68. Matsuura T, Fang P, Pearson CE, Jayakar P, Ashizawa T, Roa BB, Nelson DL. 2006. Interruptions in the Expanded ATTCT Repeat of Spinocerebellar Ataxia Type 10: Repeat Purity as a Disease Modifier? Am J Hum Genet 78: 125–129.

69. Mikheenko A, Prjibelski A, Saveliev V, Antipov D, Gurevich A. 2018. Versatile genome assembly evaluation with QUAST-LG. Bioinformatics 34: i142–i150.

70. Miller DE, Sulovari A, Wang T, Loucks H, Hoekzema K, Munson KM, Lewis AP, Fuerte EPA, Paschal CR, Walsh T, et al. 2021. Targeted long-read sequencing identifies missing disease-causing variation. The American Journal of Human Genetics 108: 1436–1449.

71. Morato Torres CA, Zafar F, Tsai Y-C, Vazquez JP, Gallagher MD, McLaughlin I, Hong K, Lai J, Lee J, Chirino-Perez A, et al. 2022. ATTCT and ATTCC repeat expansions in the ATXN10 gene affect disease penetrance of spinocerebellar ataxia type 10. Human Genetics and Genomics Advances 3: 100137.

72. Morris CA. 1993. Williams Syndrome. In GeneReviews® (eds. M.P. Adam, J. Feldman, G.M. Mirzaa, R.A. Pagon, S.E. Wallace, L.J. Bean, K.W. Gripp, and A. Amemiya), University of Washington, Seattle, Seattle (WA) http://www.ncbi.nlm.nih.gov/books/NBK1249/ (Accessed February 24, 2024).

73. Mousavi N, Margoliash J, Pusarla N, Saini S, Yanicky R, Gymrek M. 2021. TRTools: a toolkit for genome-wide analysis of tandem repeats. Bioinformatics 37: 731–733.

74. Noyvert B, Erzurumluoglu AM, Drichel D, Omland S, Andlauer TFM, Mueller S, Sennels L, Becker C, Kantorovich A, Bartholdy BA, et al. 2023. Imputation of structural variants using a multi-ancestry long-read sequencing panel enables identification of disease associations. 2023.12.20.23300308. https://www.medrxiv.org/content/10.1101/2023.12.20.23300308v1 (Accessed December 23, 2023).

75. Nurk S, Koren S, Rhie A, Rautiainen M, Bzikadze AV, Mikheenko A, Vollger MR, Altemose N, Uralsky L, Gershman A, et al. 2022. The complete sequence of a human genome. Science 376: 44–53.

76. Ortiz-Aljaro P, Montes-Cano MA, García-Lozano J-R, Aquino V, Carmona R, Perez-Florido J, García-Hernández FJ, Dopazo J, González-Escribano MF. 2022. Protein and functional isoform levels and genetic variants of the BAFF and APRIL pathway components in systemic lupus erythematosus. Sci Rep 12: 11219.

77. Pedersen BS, Bhetariya PJ, Brown J, Kravitz SN, Marth G, Jensen RL, Bronner MP, Underhill HR, Quinlan AR. 2020. Somalier: rapid relatedness estimation for cancer and germline studies using efficient genome sketches. Genome Medicine 12: 62.

78. Pellerin D, Danzi MC, Wilke C, Renaud M, Fazal S, Dicaire M-J, Scriba CK, Ashton C, Yanick C, Beijer D, et al. 2023. Deep Intronic FGF14 GAA Repeat Expansion in Late-Onset Cerebellar Ataxia. N Engl J Med 388: 128–141.

79. Quinlan AR, Hall IM. 2010. BEDTools: a flexible suite of utilities for comparing genomic features. Bioinformatics 26: 841–842.

80. R Core Team. 2021. R: A language and environment for statistical computing. R Foundation for Statistical Computing. https://www.R-project.org/.

81. Raskin S, Ashizawa T, Teive HAG, Arruda WO, Fang P, Gao R, White MC, Werneck LC, Roa B. 2007. Reduced penetrance in a Brazilian family with spinocerebellar ataxia type 10. Arch Neurol 64: 591–594.

82. Razaghi R, Hook PW, Ou S, Schatz MC, Hansen KD, Jain M, Timp W. 2022. Modbamtools: Analysis of single-molecule epigenetic data for long-range profiling, heterogeneity, and clustering. 2022.07.07.499188. https://www.biorxiv.org/content/10.1101/2022.07.07.499188v1 (Accessed February 18, 2024).

83. Reis ALM, Rapadas M, Hammond JM, Gamaarachchi H, Stevanovski I, Ayuputeri Kumaheri M, Chintalaphani SR, Dissanayake DSB, Siggs OM, Hewitt AW, et al. 2023. The landscape of genomic structural variation in Indigenous Australians. Nature 624: 1–9.

84. Ren J, Gu B, Chaisson MJP. 2023. vamos: variable-number tandem repeats annotation using efficient motif sets. Genome Biology 24: 175.

85. Saal HM, Harbison MD, Netchine I. 1993. Silver-Russell Syndrome. In GeneReviews® (eds. M.P. Adam, J. Feldman, G.M. Mirzaa, R.A. Pagon, S.E. Wallace, L.J. Bean, K.W. Gripp, and A. Amemiya), University of Washington, Seattle, Seattle (WA) http://www.ncbi.nlm.nih.gov/books/NBK1324/ (Accessed February 7, 2024).

86. Scriba CK, Beecroft SJ, Clayton JS, Cortese A, Sullivan R, Yau WY, Dominik N, Rodrigues M, Walker E, Dyer Z, et al. 2020. A novel RFC1 repeat motif (ACAGG) in two Asia-Pacific CANVAS families. Brain 143: 2904–2910.

87. Shafin K, Pesout T, Chang P-C, Nattestad M, Kolesnikov A, Goel S, Baid G, Kolmogorov M, Eizenga JM, Miga KH, et al. 2021. Haplotype-aware variant calling with PEPPER-Margin-DeepVariant enables high accuracy in nanopore long-reads. Nat Methods 18: 1322–1332.

88. Shafin K, Pesout T, Lorig-Roach R, Haukness M, Olsen HE, Bosworth C, Armstrong J, Tigyi K, Maurer N, Koren S, et al. 2020. Nanopore sequencing and the Shasta toolkit enable efficient de novo assembly of eleven human genomes. Nat Biotechnol 38: 1044–1053.

89. Shuman C, Kalish JM, Weksberg R. 1993. Beckwith-Wiedemann Syndrome. In GeneReviews® (eds. M.P. Adam, J. Feldman, G.M. Mirzaa, R.A. Pagon, S.E. Wallace, L.J. Bean, K.W. Gripp, and A. Amemiya), University of Washington, Seattle, Seattle (WA) http://www.ncbi.nlm.nih.gov/books/NBK1394/ (Accessed February 7, 2024).

90. Smolka M, Paulin LF, Grochowski CM, Horner DW, Mahmoud M, Behera S, Kalef-Ezra E, Gandhi M, Hong K, Pehlivan D, et al. 2024. Detection of mosaic and population-level structural variants with Sniffles2. Nat Biotechnol 1–10.

91. Subramanian RP, Wildschutte JH, Russo C, Coffin JM. 2011. Identification, characterization, and comparative genomic distribution of the HERV-K (HML-2) group of human endogenous retroviruses. Retrovirology 8: 90.

92. Sulovari A, Li R, Audano PA, Porubsky D, Vollger MR, Logsdon GA, Human Genome Structural Variation Consortium, Warren WC, Pollen AA, Chaisson MJP, et al. 2019. Human-specific tandem repeat expansion and differential gene expression during primate evolution. Proc Natl Acad Sci U S A 116: 23243–23253.

93. Tanudisastro HA, Deveson IW, Dashnow H, MacArthur DG. 2024. Sequencing and characterizing short tandem repeats in the human genome. Nat Rev Genet 1–16.

94. Taylor DJ, Chhetri SB, Tassia MG, Biddanda A, Battle A, McCoy RC. 2023. Sources of gene expression variation in a globally diverse human cohort. 2023.11.04.565639. https://www.biorxiv.org/content/10.1101/2023.11.04.565639v4 (Accessed February 22, 2024).

95. Twesigomwe D, Drögemöller BI, Wright GEB, Adebamowo C, Agongo G, Boua PR, Matshaba M, Paximadis M, Ramsay M, Simo G, et al. 2024. Characterization of CYP2B6 and CYP2A6 Pharmacogenetic Variation in Sub-Saharan African Populations. Clin Pharmacol Ther 115: 576–594.

96. Twesigomwe D, Drögemöller BI, Wright GEB, Adebamowo C, Agongo G, Boua PR, Matshaba M, Paximadis M, Ramsay M, Simo G, et al. 2023. Characterization of CYP2D6 Pharmacogenetic Variation in Sub-Saharan African Populations. Clinical Pharmacology & Therapeutics 113: 643–659.

97. Wagner J, Olson ND, Harris L, Khan Z, Farek J, Mahmoud M, Stankovic A, Kovacevic V, Yoo B, Miller N, et al. 2022. Benchmarking challenging small variants with linked and long reads. Cell Genomics 2: 100128.

98. Wang T, Antonacci-Fulton L, Howe K, Lawson HA, Lucas JK, Phillippy AM, Popejoy AB, Asri M, Carson C, Chaisson MJP, et al. 2022. The Human Pangenome Project: a global resource to map genomic diversity. Nature 604: 437–446.

99. Wang Y, Zhao Y, Bollas A, Wang Y, Au KF. 2021. Nanopore sequencing technology, bioinformatics and applications. Nat Biotechnol 39: 1348–1365.

100. Wilderman A, D’haene E, Baetens M, Yankee TN, Winchester EW, Glidden N, Roets E, Van Dorpe J, Janssens S, Miller DE, et al. 2024. A distant global control region is essential for normal expression of anterior HOXA genes during mouse and human craniofacial development. Nat Commun 15: 1–23.

101. Wojcik MH, Reuter CM, Marwaha S, Mahmoud M, Duyzend MH, Barseghyan H, Yuan B, Boone PM, Groopman EE, Délot EC, et al. 2023. Beyond the exome: What’s next in diagnostic testing for Mendelian conditions. The American Journal of Human Genetics 110: 1229– 1248.

102. Yang L, Metzger GA, Padilla Del Valle R, Delgadillo Rubalcaba D, McLaughlin RN. 2024. Evolutionary insights from profiling LINE-1 activity at allelic resolution in a single human genome. EMBO J 43: 112–131.

103. Zalusky MP, Miller DE. 2024. Methylation Operation Wizard (MeOW): Identification of differentially methylated regions in long-read sequencing data. http://arxiv.org/abs/2402.17182 (Accessed February 27, 2024).

104. Zanger UM, Schwab M. 2013. Cytochrome P450 enzymes in drug metabolism: regulation of gene expression, enzyme activities, and impact of genetic variation. Pharmacol Ther 138: 103–141.

105. Zhao X, Collins RL, Lee W-P, Weber AM, Jun Y, Zhu Q, Weisburd B, Huang Y, Audano PA, Wang H, et al. 2021. Expectations and blind spots for structural variation detection from long-read assemblies and short-read genome sequencing technologies. Am J Hum Genet 108: 919–928.

106. Zheng Z, Li S, Su J, Leung AW-S, Lam T-W, Luo R. 2022. Symphonizing pileup and full-alignment for deep learning-based long-read variant calling. Nat Comput Sci 2: 797–803.

107. Zook JM, Hansen NF, Olson ND, Chapman L, Mullikin JC, Xiao C, Sherry S, Koren S, Phillippy AM, Boutros PC, et al. 2020. A robust benchmark for detection of germline large deletions and insertions. Nat Biotechnol 38: 1347–1355.

