## Supplemental Figures for "Nanopore sequencing of 1000 Genomes Project samples to build a comprehensive catalog of human genetic variation"

**Figure S1. Precision, recall, and F1 scores for SNV and indel calls for ONT and Illumina small variant calls using PacBio assemblies as the truth set.**

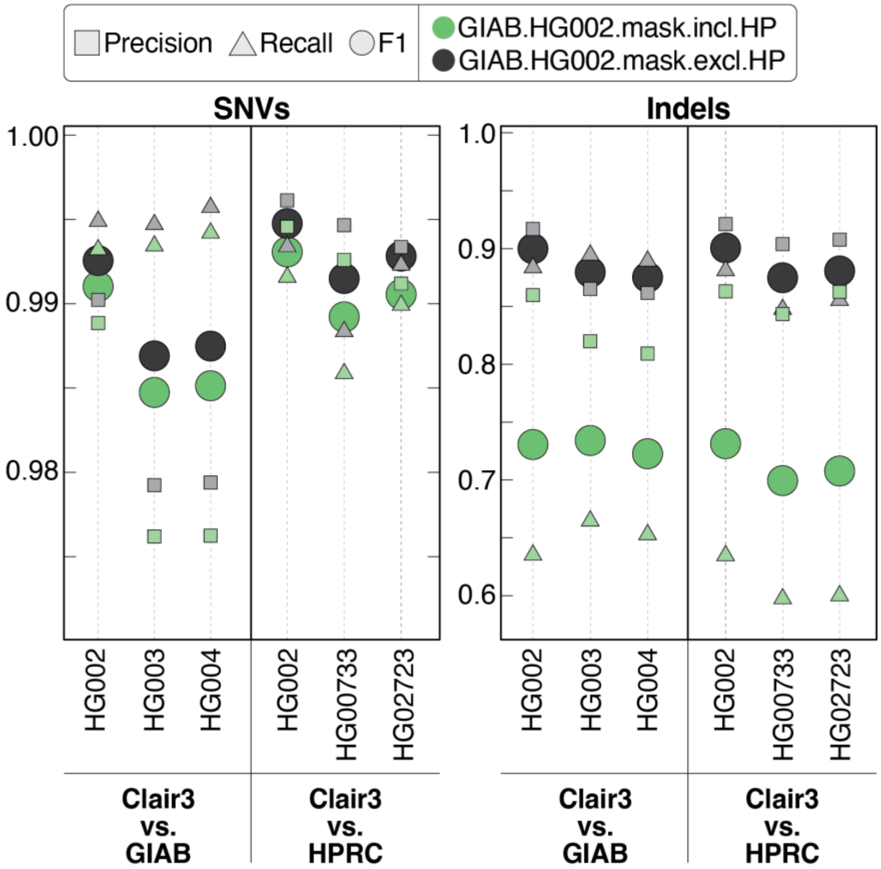

**Figure S2. Precision, recall, and F1 scores for SNV and indel calls for ONT small variant calls from both the Napu (PMDV) and alignment (Clair3) pipelines using Illumina data as the truth set.**
Variants called using PMDV are shown without masking (A), results that include masking can be found in the main text. Variants called by Clair3 are shown genome-wide (B), in GIAB high-confidence regions only (C), and in GIAB high-confidence regions excluding homopolymers 4 nucleotides or greater +/- one nucleotide on either side (D).

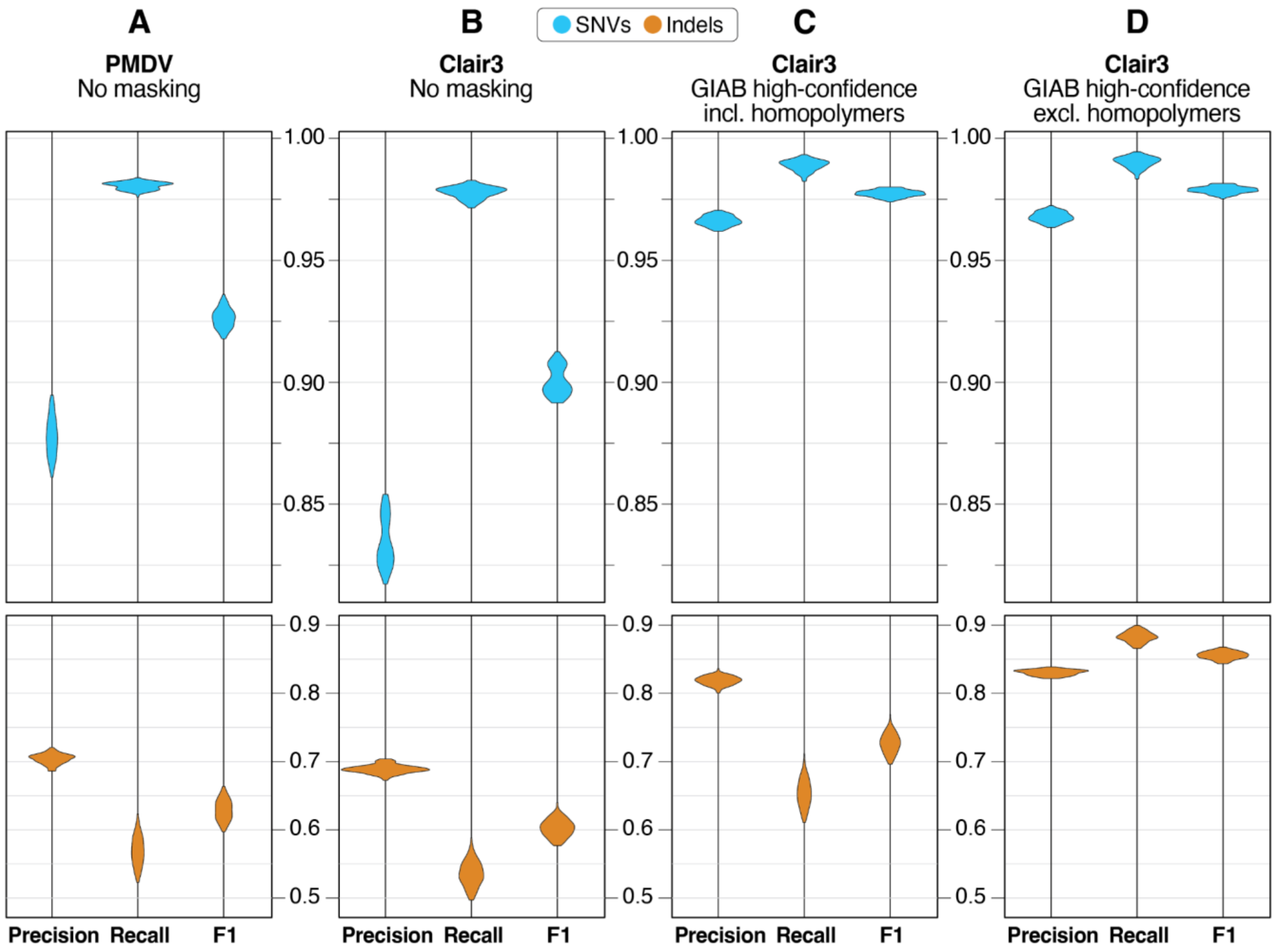

**Figure S3. Percent of genome assembled by Flye and Shasta-Hapdup.**
Violin plot showing the percentage of genome assembled for each sample by Flye and Shasta-Hapdup. The *x*-axis represents the assembler and the *y*-axis represents the percentage of GRCh38 reference genome covered by contigs for each assembly. The median values for Flye and Shasta-Hapdup assemblies for all 100 samples are 93.6% and 93.5%, respectively.

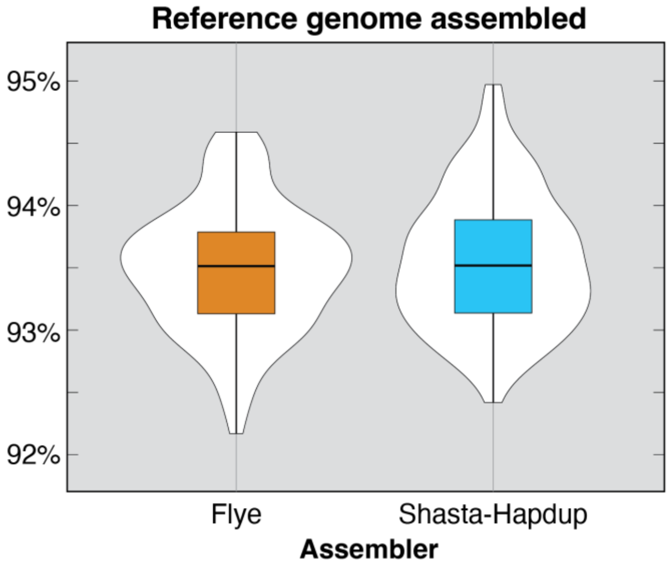

**Figure S4. IGV screenshots of chromosome 7 assemblies** (*see next page*).
**A**. Example of a location where the presence of a segmental duplication is associated with breaks in assemblies in both Flye and Shasta-Hapdup for HG02373. **B**. Example of a region with a discontinuity in the Flye assembly contigs in GM19035. This break occurs in a region outside of a segmental duplication or a satellite repeat. The alignment and coverage tracks at the top suggest no aberrations or increased coverage that could indicate increased likelihood of an assembly break. IGV view is approximately 2,900 bp. **C**. Example of a position where Shasta-Hapdup assembled contigs break in both haplotype-resolved assemblies in HG01801. This break occurs in a region outside of a segmental duplication or satellite repeat. The alignment and coverage tracks at the top suggest no aberrations or increased coverage that could indicate increased likelihood of assembly breaks. IGV view is approximately 23 kbp.

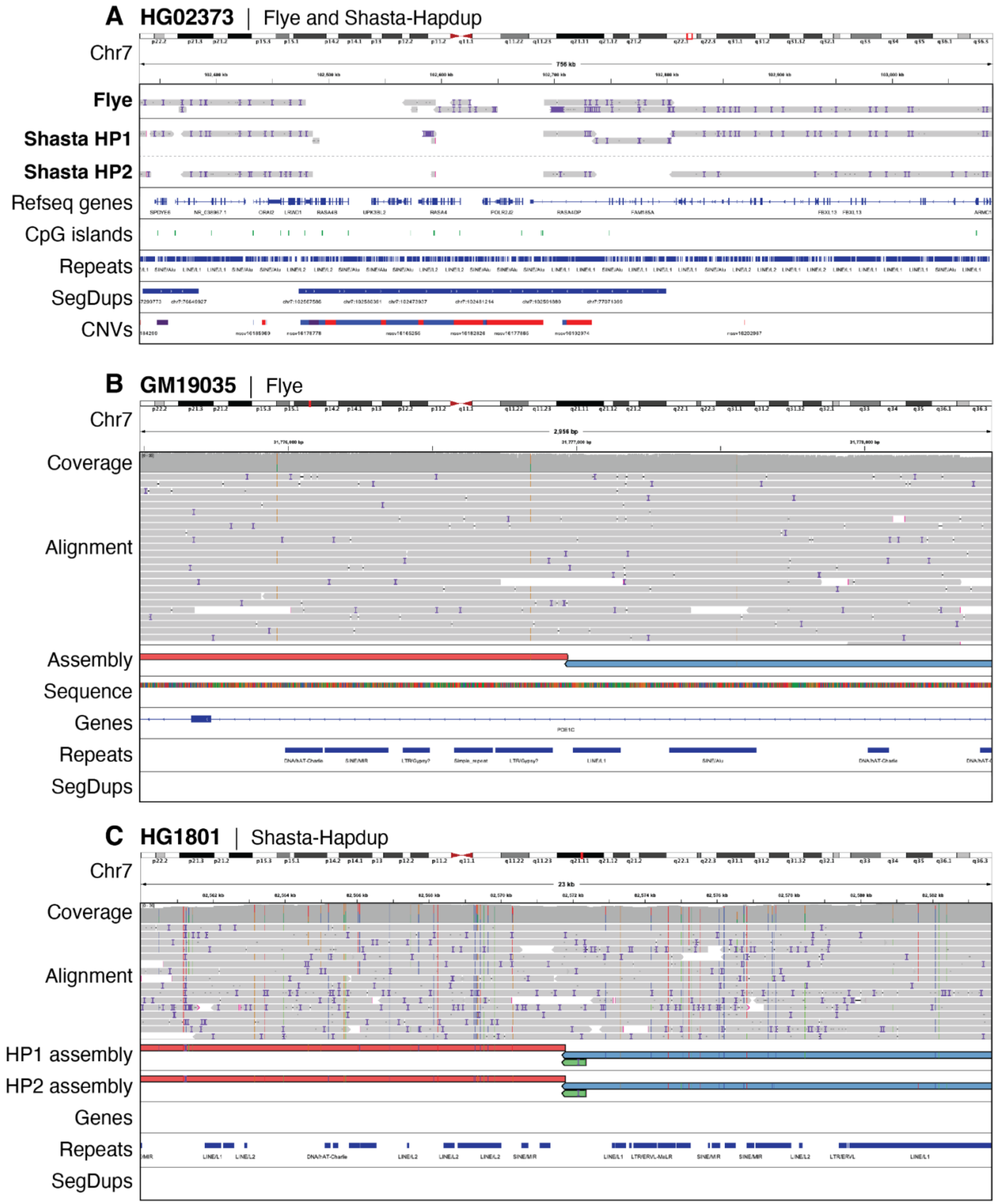
A HG02373 | Flye and Shasta-Hapdup
Chr7
Flye
Shasta HP1
Shasta HP2
Refseq genes
CpG islands
Repeats
SegDups
CNVs
B GM19035 | Flye
Chr7
Coverage
Alignment
Assembly
Sequence
Genes
Repeats
SegDups
C HG1801 | Shasta-Hapdup
Chr7
Coverage
Alignment
HP1 assembly
HP2 assembly
Genes
Repeats
SegDups

**Figure S5. Pangenome principal component analysis** (*see next page*).

**A.** Graph-based principal component analysis of chromosome 20 assemblies – including 100 Shasta-Hapdup samples, 44 HPRC samples, and the GRCh38, CHM13 (v2.0), and HG002 (v1.0.1) human reference genomes – revealed clear separation along the first principal component (PC1). The two clusters correspond to the HPRCy1 assemblies (right) and 100 Shasta-Hapdup assemblies (left). Because our assemblies cluster close to GRCh38, whose centromeric sequences were masked, we interpreted PC1 as representing centromeric diversity of the assemblies. Experience with ONT based assemblies suggests that the ONT-only assemblies are currently less able to assemble highly repetitive sequences compared to those which include HiFi data. **B.** Graph-based principal component analysis of euchromatic, non-centromeric fraction of the chromosome 20 assemblies—including 100 Shasta-Hapdup samples, 44 HPRC samples, and the GRCh38, CHM13 (v2.0), and HG002 (v1.0.1) human reference genomes—revealed no distinct separation between the Shasta-Hapdup and HPRC samples. The 4 reference genomes cluster because their entire sequence is considered, including the heterochromatic regions. Data points were visually distinguished by color and shape according to their group. In general, our assemblies are slightly more spread in both PC1 and PC2, which possibly relates to higher base-level error rates in the ONT-only assemblies.

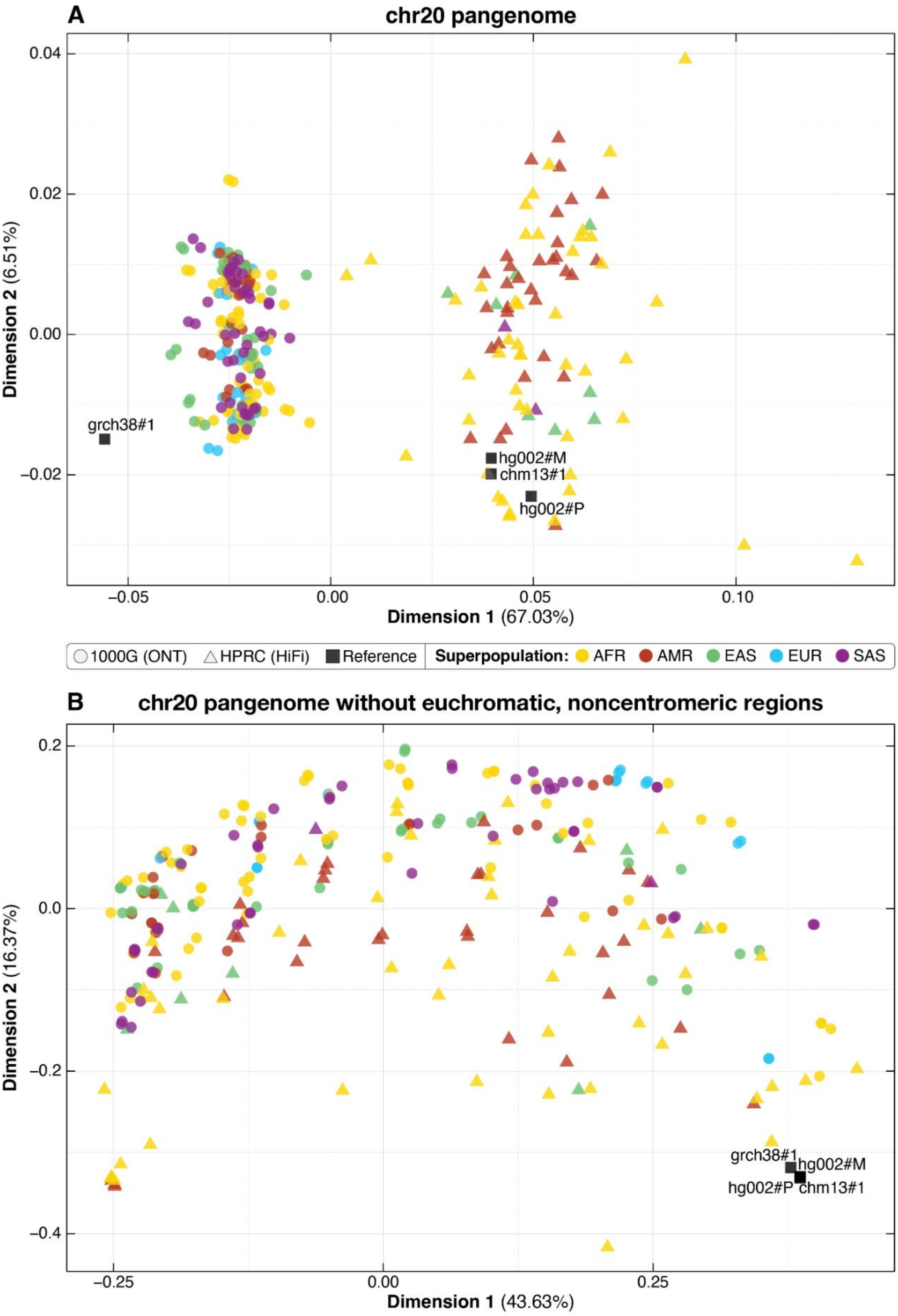
A
chr20 pangenome
0.04
0.02
0.00
−0.02
Dimension 2 (6.51%)
grch38#1
hg002#M
chm13#1
hg002#P
−0.05
0.00
0.05
0.10
Dimension 1 (67.03%)
1000G (ONT)
HPRC (HiFi)
Reference
Superpopulation: AFR AMR EAS EUR SAS
B
chr20 pangenome without euchromatic, noncentromeric regions
0.2
0.0
−0.2
−0.4
Dimension 2 (16.37%)
grch38#1
hg002#M
hg002#P
chm13#1
−0.25
0.00
0.25
Dimension 1 (43.63%)

**Figure S6. Evaluation of L1HS elements in the first 100 assemblies.**
Comparison of the number of L1HS in the major populations in the 100 samples (standard boxplots) against the number identified in phased haplotypes from HG002 and HG005 from GIAB and the haploid CHM13 T2T assembly.

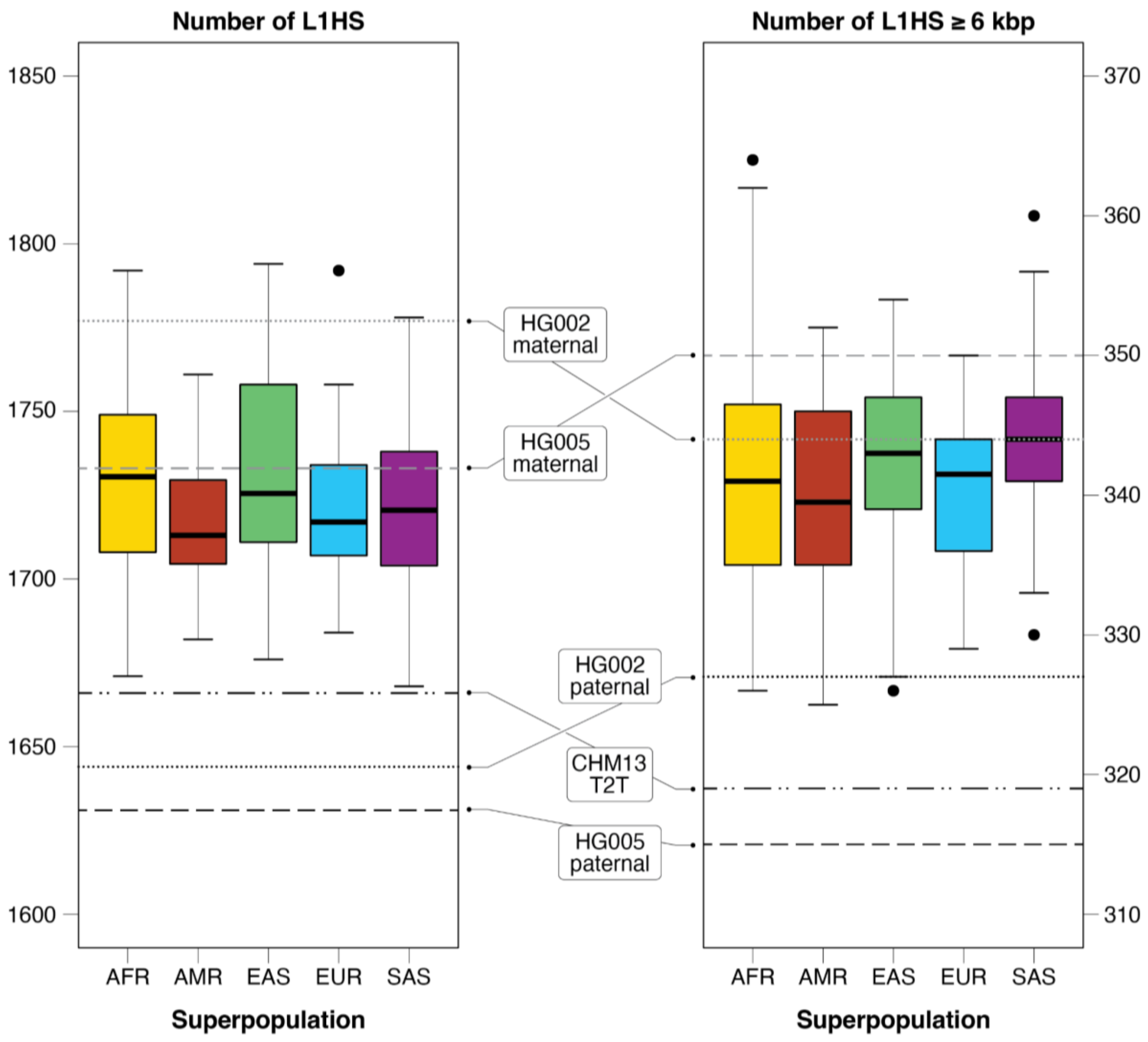

**Figure S7. Evaluation of per-sample insertions and deletions by type, chromosome, and SV caller.** The number of insertion and deletion SVs was evaluated by chromosome for each of the five SV callers across all 100 samples.

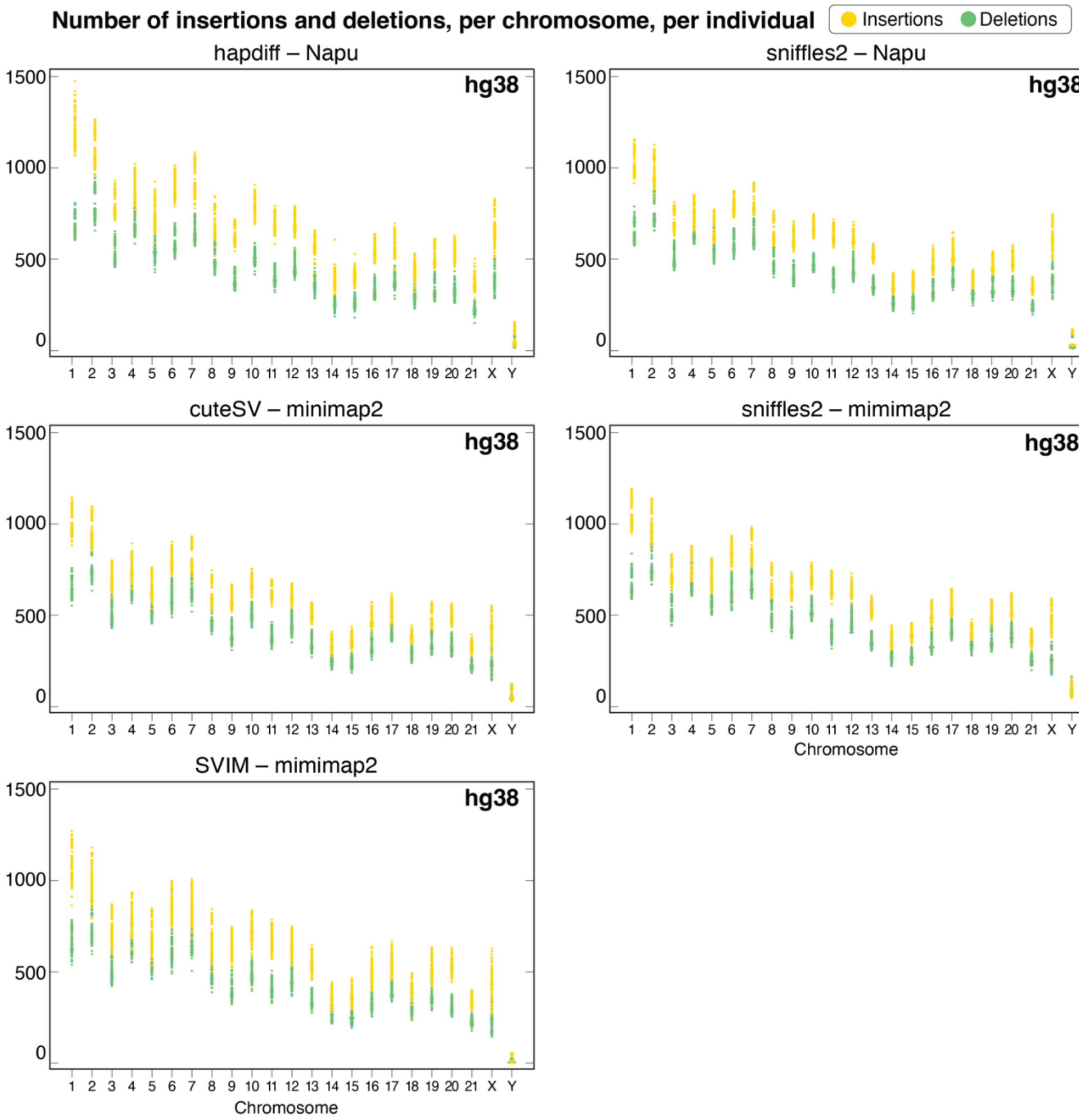

**Figure S8. Distribution of SV events genome-wide, all populations.**

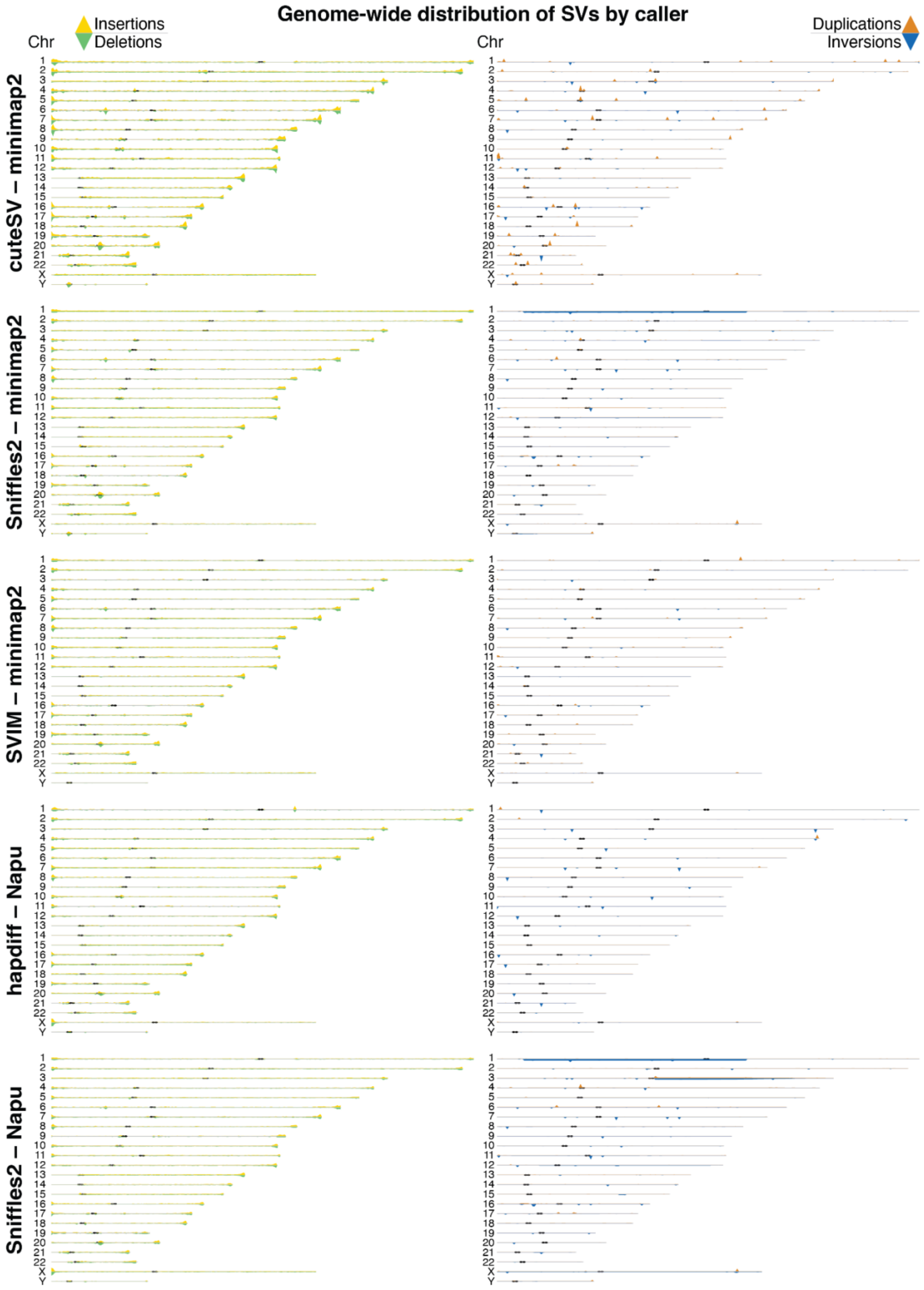

**Figure S9. Concordance of SVs by caller and evaluation of joint SV calls.**
**A,C**. Concordance of SV calls for all callers (A) and for only hapdiff (C) for insertion and deletion events. **B**. Number of SVs per individual called by at least 1, 2, 3 ,4 and 5 (all) callers. Based on Jasmine merging of all 5 SV callers per sample (cuteSV–minimap2, Sniffles2–minimap2, SVIM–minimap2, hapdiff–Napu, Sniffles2–minimap2). If an SV was called by at least one caller (in other words, called at all by any caller), it has a minimum support of 1. If an SV was called in an individual in at least 2 callers, it has a minimum support of 2, etc. 5 means that all 5 callers agree on the SV. We see the same expected distribution of AFR samples having more SVs in each support threshold.

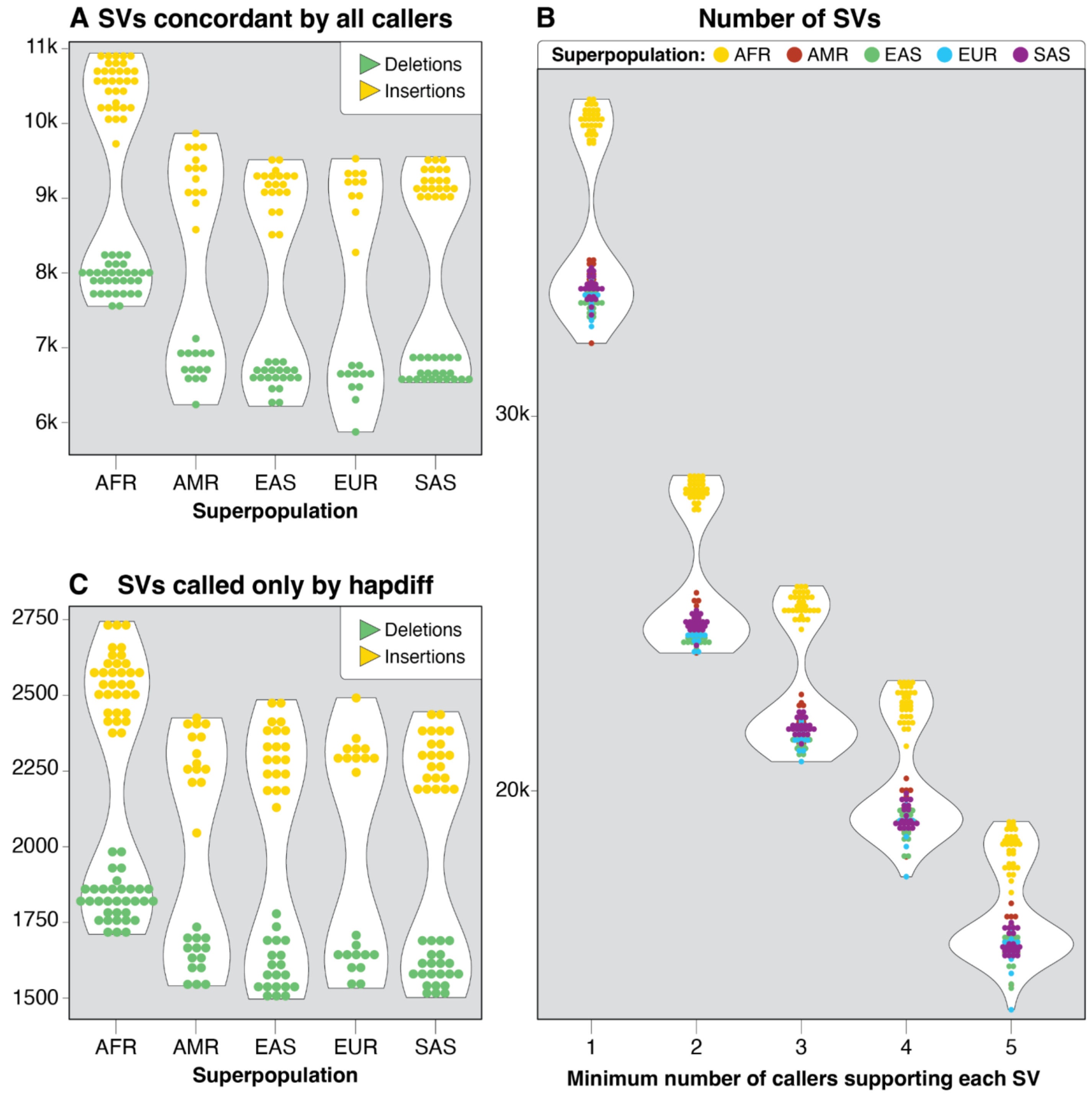

**Figure S10. Example of false positive SVs called only by hapdiff.**
We evaluated SVs called only by hapdiff to determine whether they represented false positive calls. **A**. IGV screenshot shows a single deletion in an assembly represented as two deletions in the alignment. **B**. IGV screenshot shows a call within a centromeric region found in only one assembly contig.

**A**

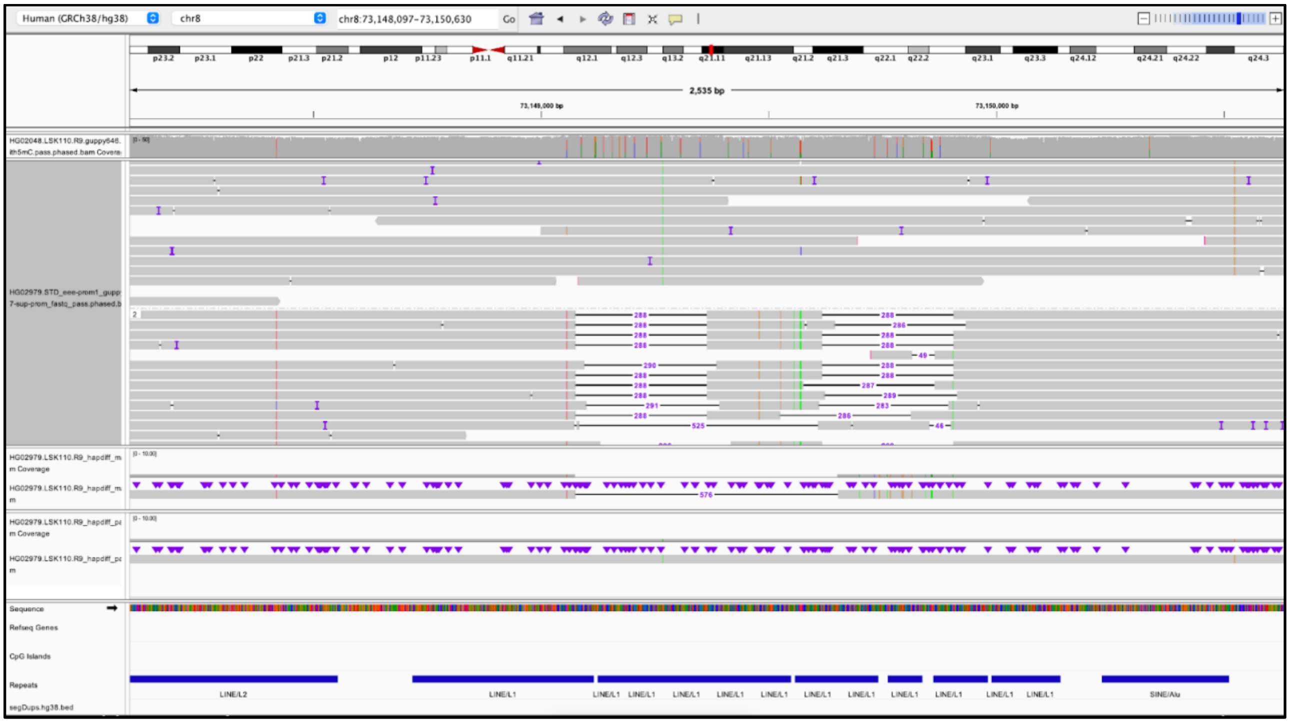

**B**

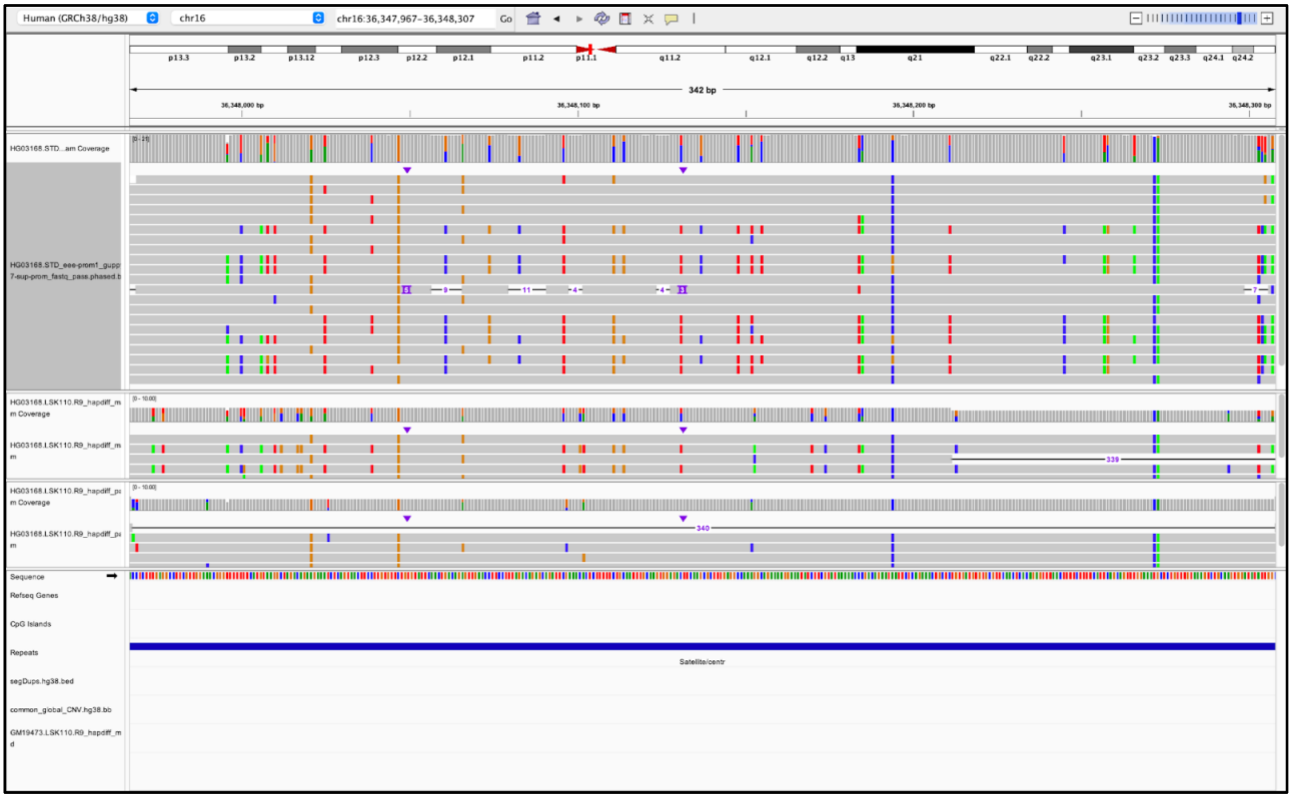

**Figure S11. Evaluating the functional impact of structural variants.**
**A.** Variant counts in the 65 samples common to both the 1000G-ONT and MAGE datasets (1 indicates heterozygous variants and 2 indicates homozygous variants). **B.** Relationship between the best-fit slope (β) derived from OLS regression and gene-level p-values. “All” refers to the use of identified SVs in selected samples for the eQTL analysis. “Novel” indicates the eQTL analysis conducted with SVs that were not reported in the previous long-read SV dataset of 31 samples by Kirsche *et al*. **C.** Integrative Genomics Viewer (IGV) screenshot of a 1235 bp deletion upstream of the 5’ TSS of the TNFSF13 gene. **D.** Distribution of genotypes and gene expression values in the 65 samples for the TNFSF13-associated deletion, along with the Benjamini-Hochberg corrected p-value. We note the p-value reported by Kirsche *et al.* is 0.045, largely due to the larger number of samples considered after short read genotyping in GTEx. **E.** IGV screenshot of a 96 bp deletion downstream of the IFFO2 gene. **F.** Distribution of genotypes and gene expression values in the 65 samples for the IFFO2-associated deletion, along with the Benjamini-Hochberg corrected p-value.

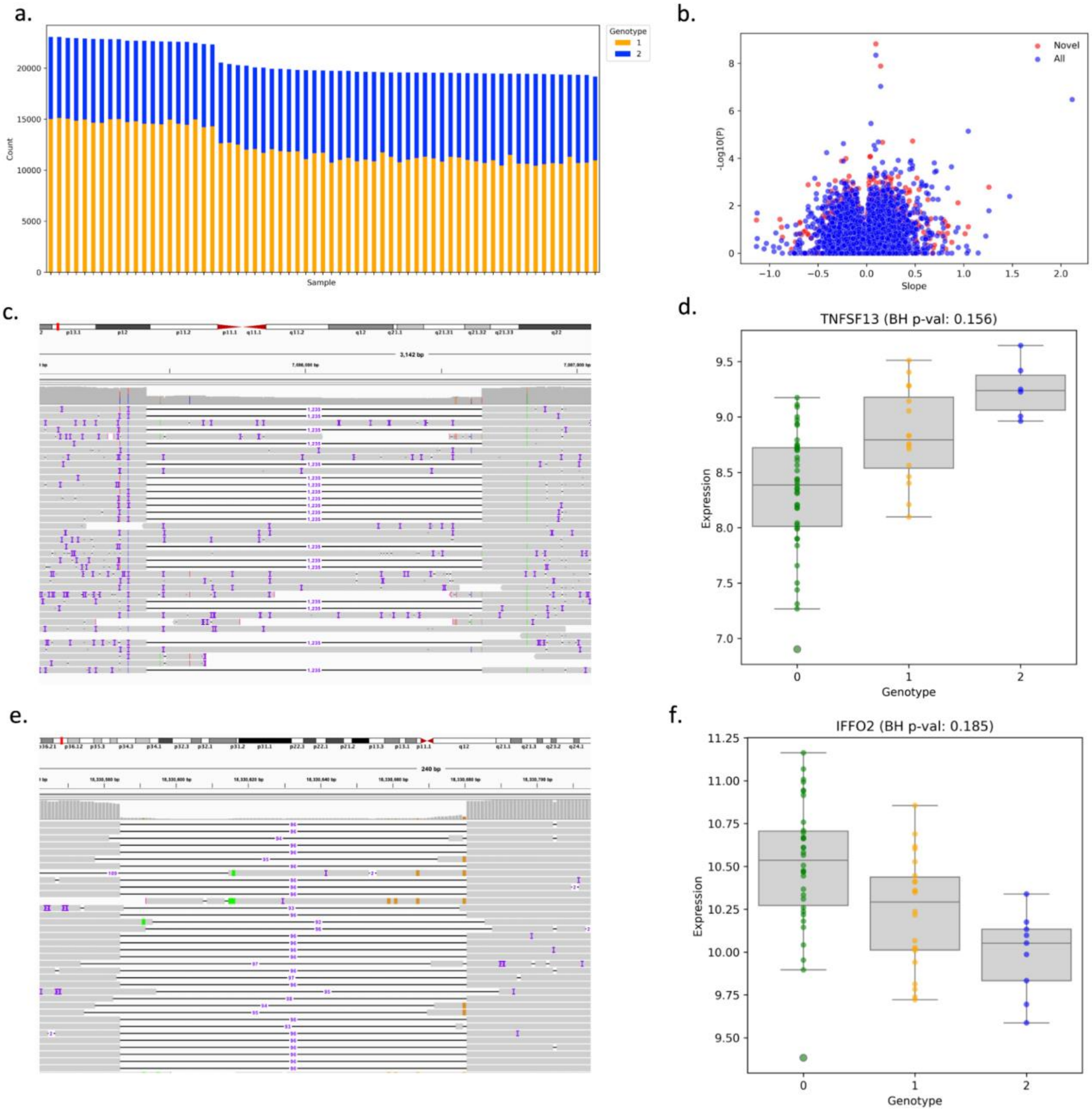

**Figure S12. Evaluation of SVs in exons of medically relevant genes.**
**A.** Number of samples harboring each INS/DEL intersecting an OMIM exon. **B.** Ideogram of where each SV that intersects with an OMIM exon is located in the GRCh38 genome

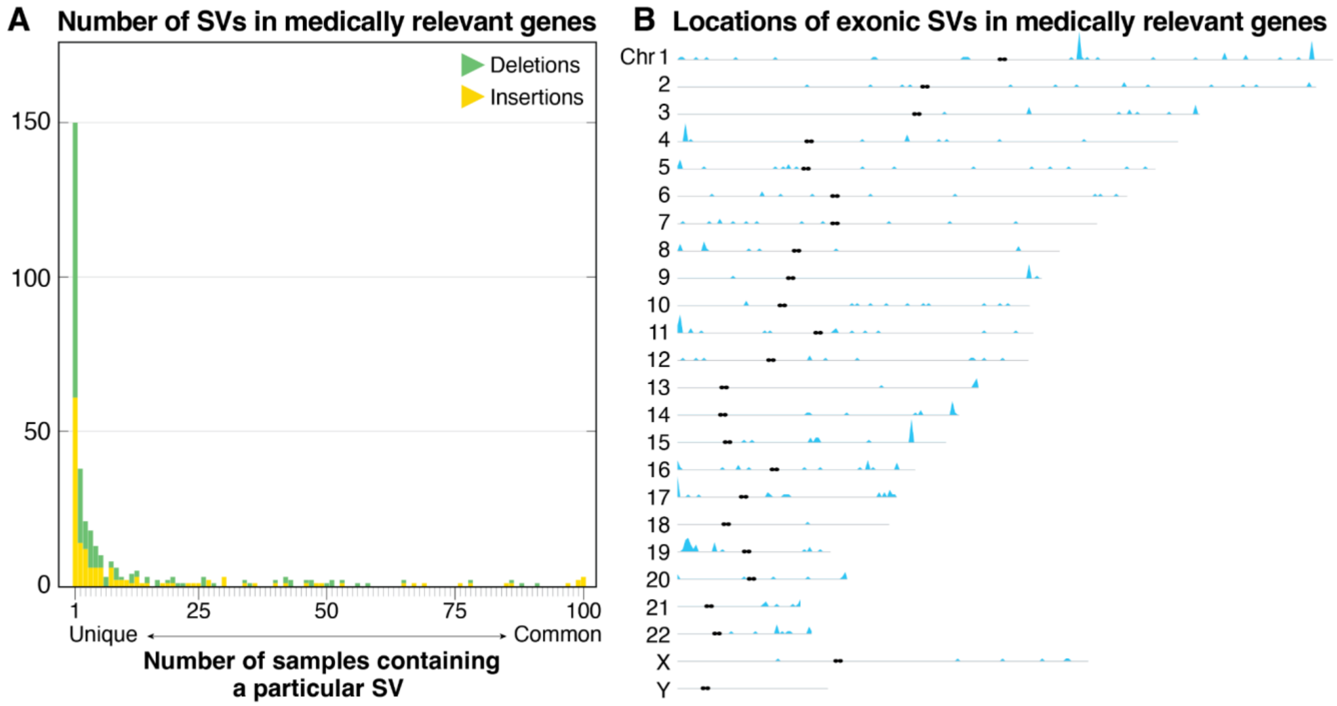

**Figure S13. IGV view of a previously described deletion that includes *HBM*, *HBA2*, *HBA1*, *HBQ1.***
A 19,304-bp deletion (NC_000016.10:g.165401_184701del) that includes *HBA1* and *HBA2* in HG01812 (Dia Chinese/EAS) and HG00728 (Southern Han Chinese/EAS). This deletion is commonly referred to as the Southeast Asian deletion (--SEA) and is associated with alpha-thalassemia (MIM: 604131).

**HG01812**

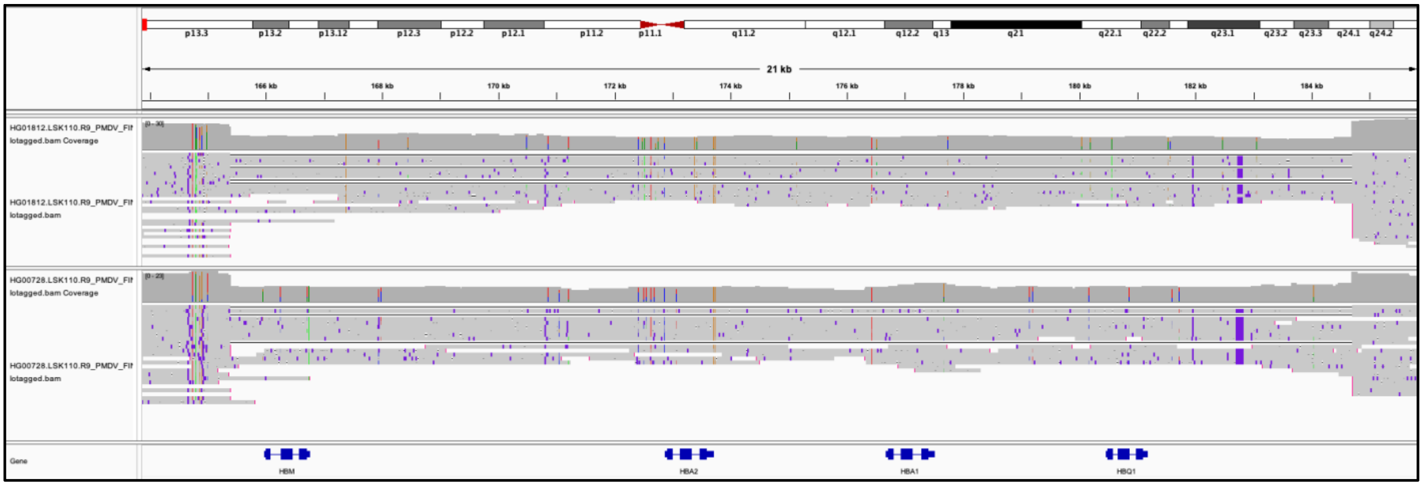

**Figure S14. Evaluation of X-linked SVs in 46,XY individuals.**
**A.** Screenshot of alignments and assemblies for an approximately 141-bp insertion in *RPGR* in GM18865. The top two tracks depict Coverage and Alignments. Track 3 and 4 show assembly-based calls derived from Shasta-Hapdup assemblies with 146-bp insertion on one haplotype and 141-bp insertion on the other. The bottom track depicts Flye assembly with a 141-bp insertion. **B.** Screenshot of short-read data from GM18865 and LRS data showing that the 141-bp insertion is not apparent in the short-read data.

**A**

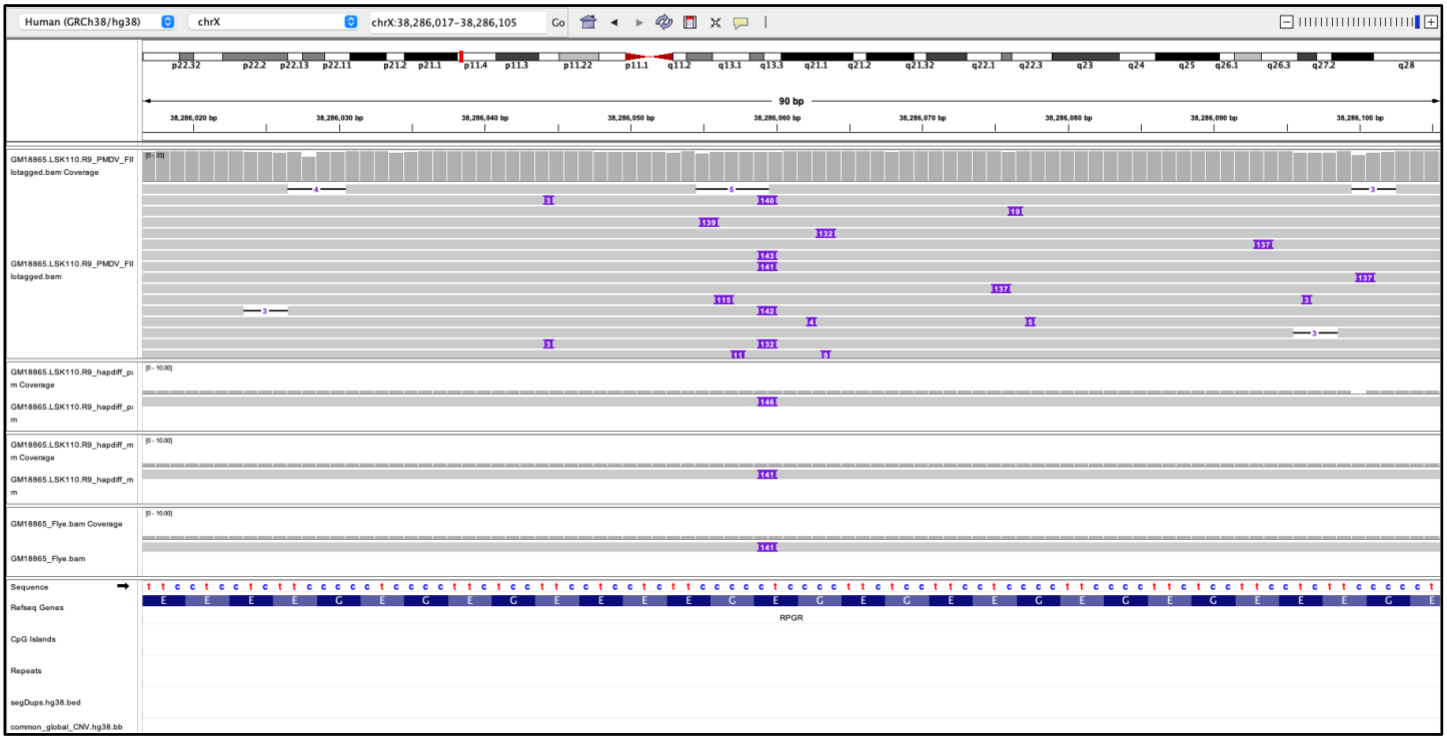

**B**

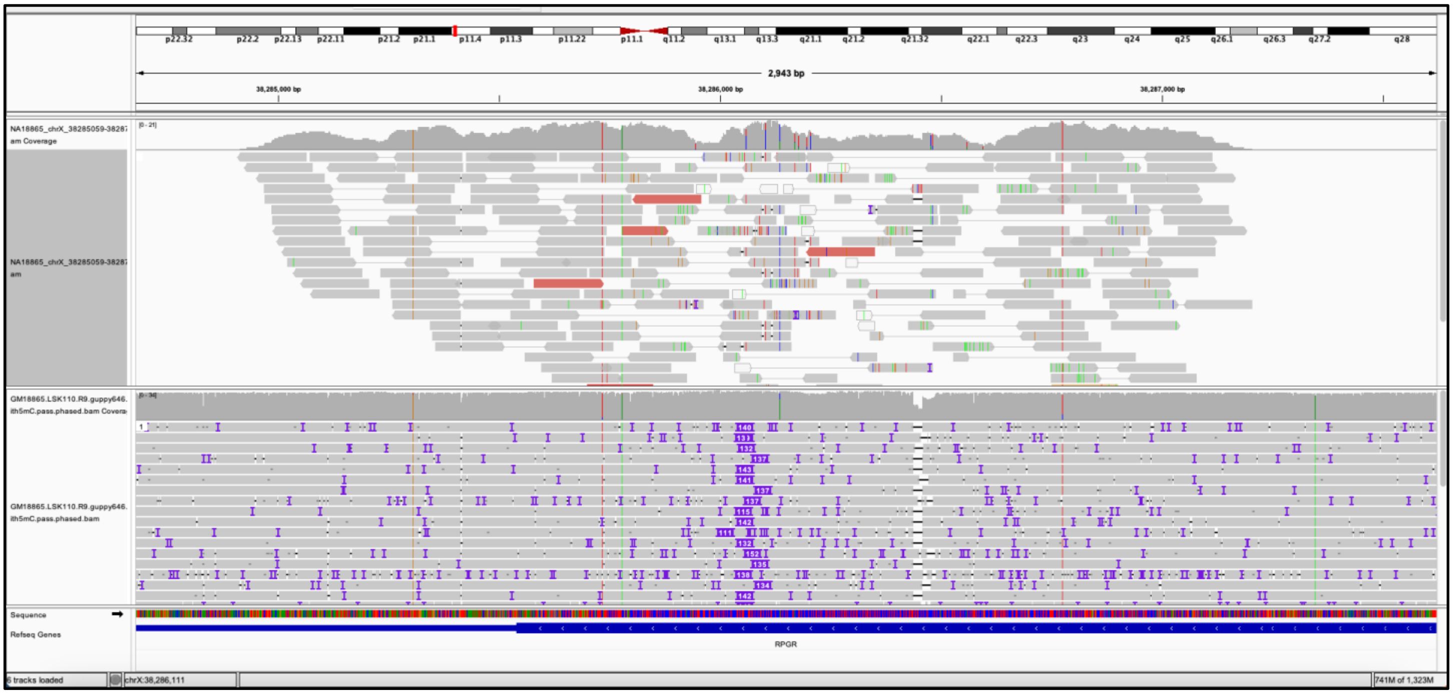

**Figure S15. Phased IGV screenshot depicting long-read alignments involving two pharmacogene hybrid star alleles.**
Long-read alignment across CYP2B6 and CYP2B7 for GM18871 from an individual from the Yoruba in Ibadan (Nigeria) who has the CYP2B6*18/*29 diplotype. Short-read alignments are included for comparison. CYP2B6*29 is a hybrid deletion allele with a CYP2B7-derived portion (across exons 1–4) and a CYP2B6-derived portion (covering exons 5-9). Read alignment results in reads aligned to either the gene (CYP2B6) or the pseudogene (CYP2B7), resulting in portions with numerous spurious SNVs.

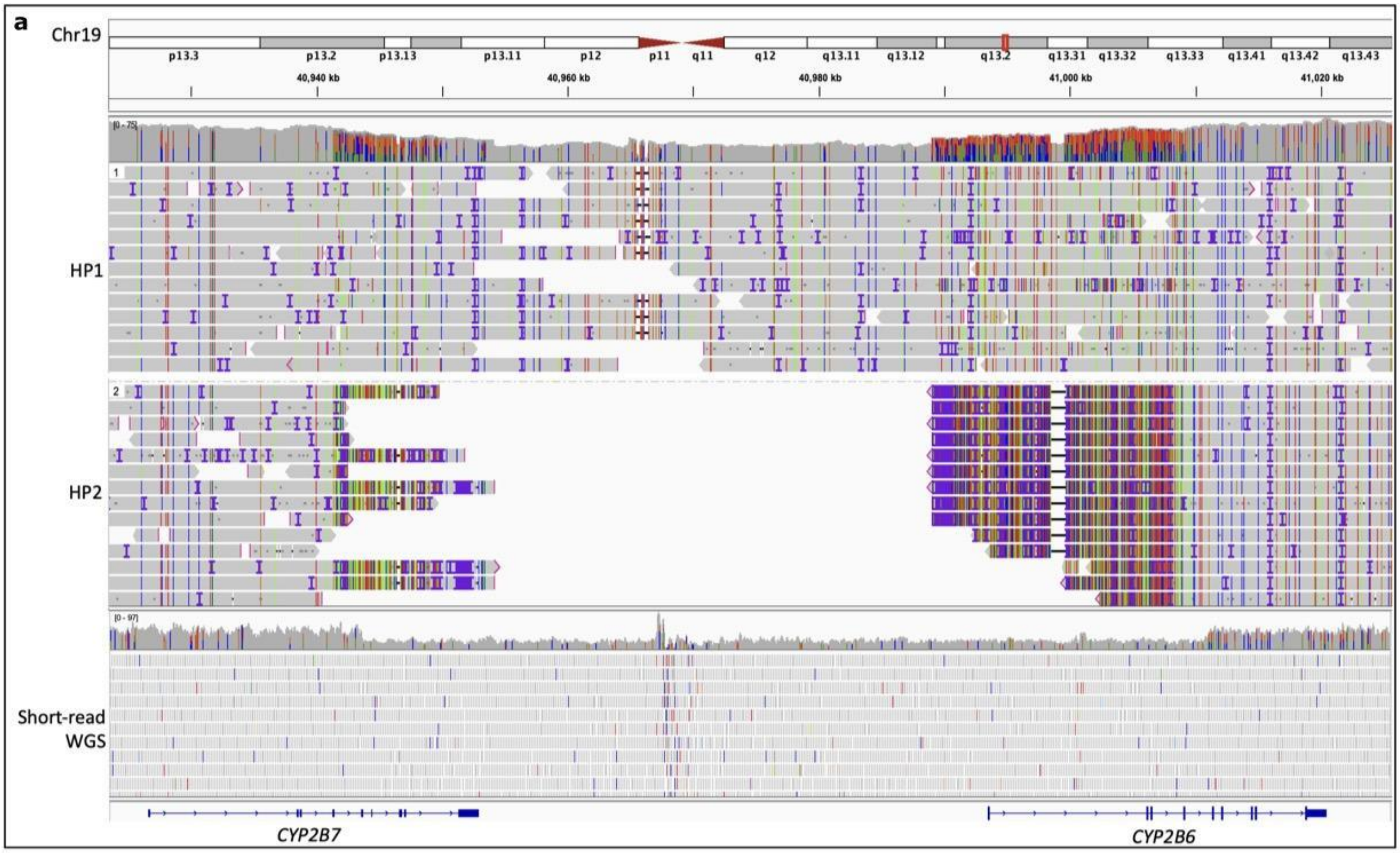

**Figure S16. Repeat expansion plots for 66 disease-associated loci.**
Violin plots for 66 disease-associated repeat expansion loci listed at STRchive: https://github.com/hdashnow/STRchive. Motif counts were genotyped with vamos and plotted with R.

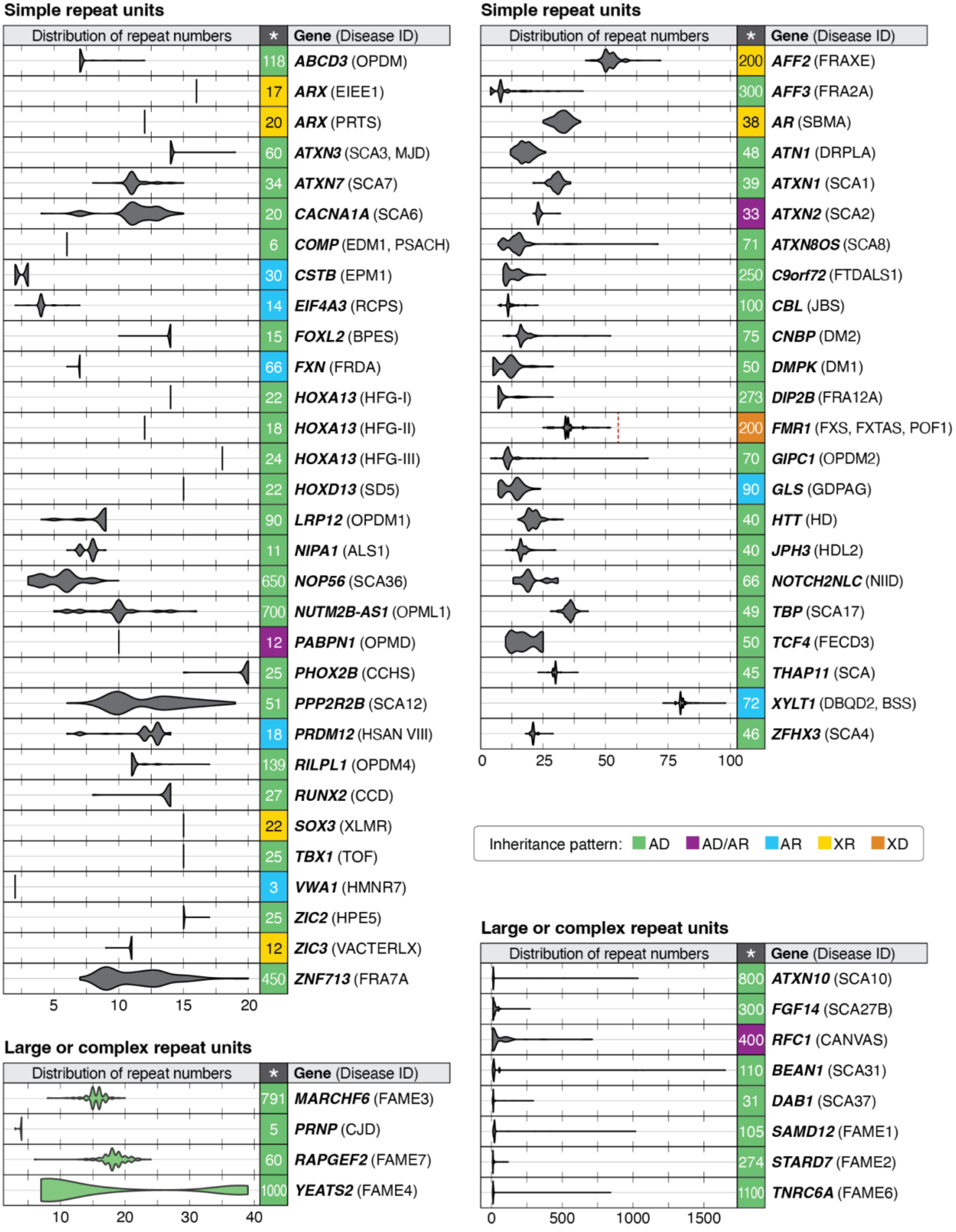

**Figure S17. IGV screenshots of repeat expansions in *RFC1* observed in 5 samples.**
IGV screenshots of reads for the five samples with the largest alleles at the CANVAS-associated locus at *RFC1*. The PMDV haplotagged alignment and the hapdiff assemblies from the NAPU pipeline for each sample are shown. The hapdiff assembly files from the NAPU pipeline were used for genotyping with vamos.

### HG00105

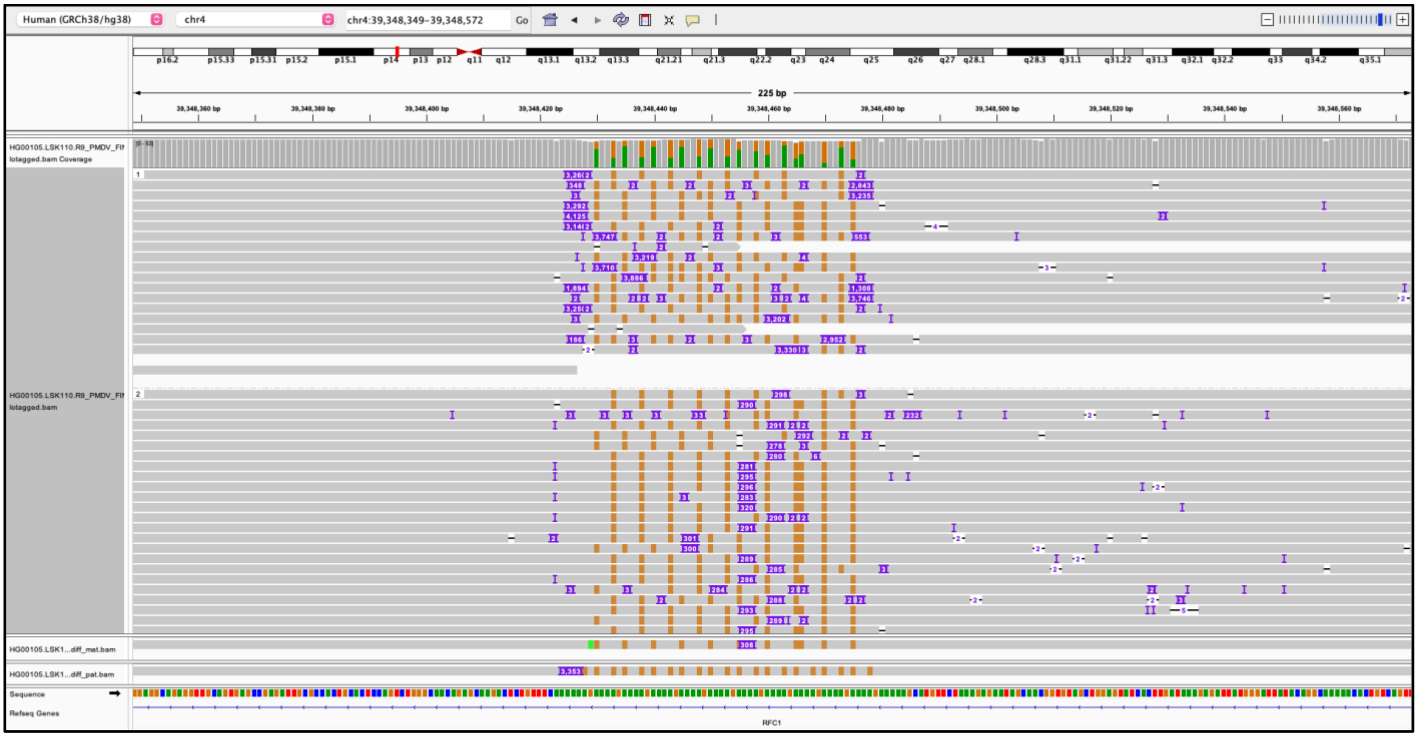

### HG00209

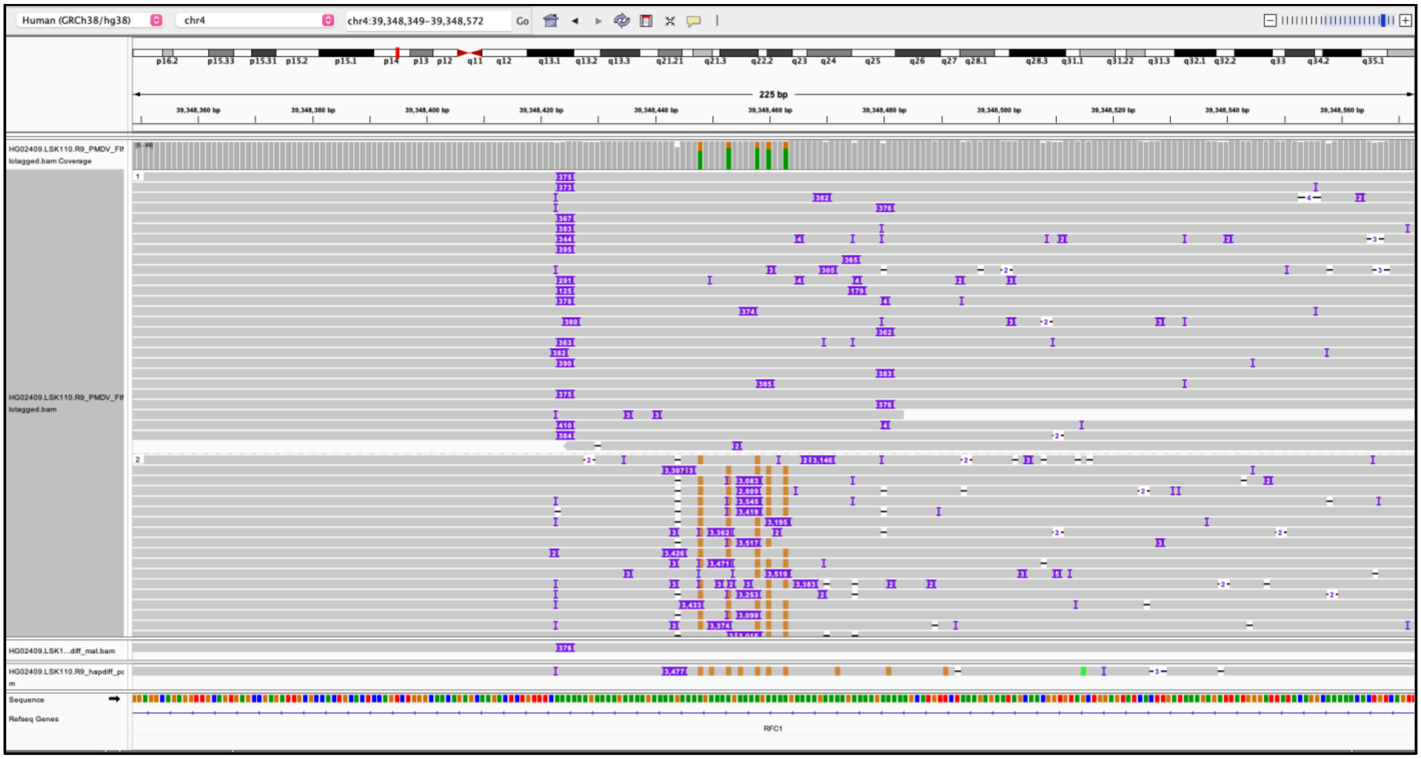

## HG01122

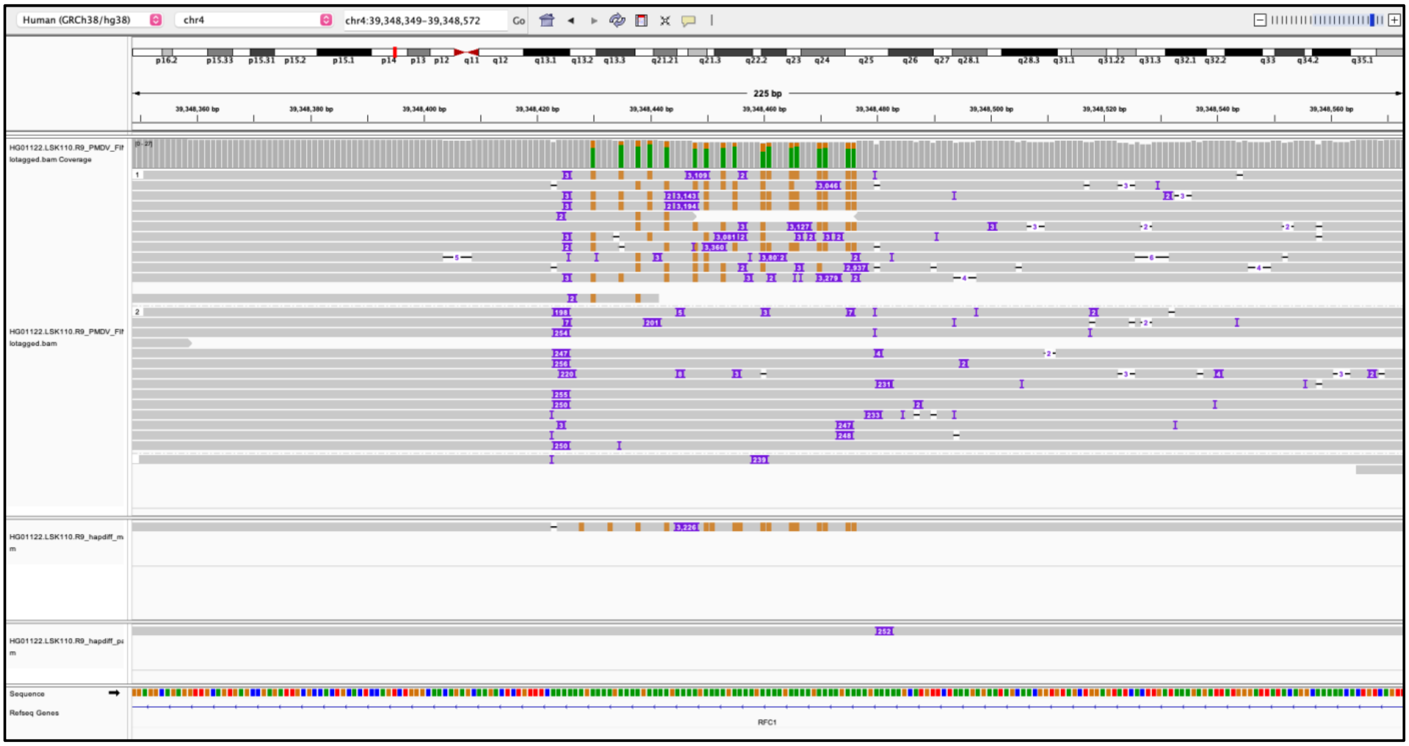

## HG00331

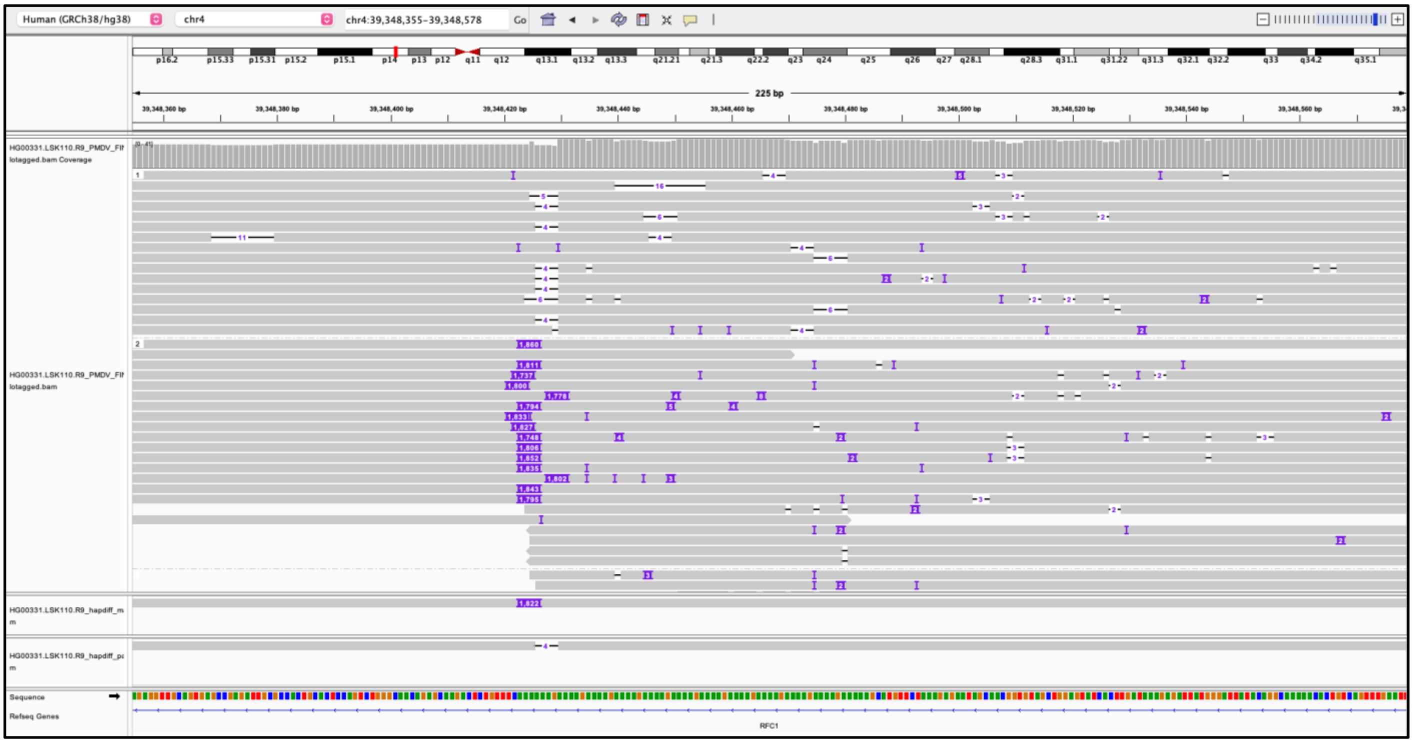

## HG01862

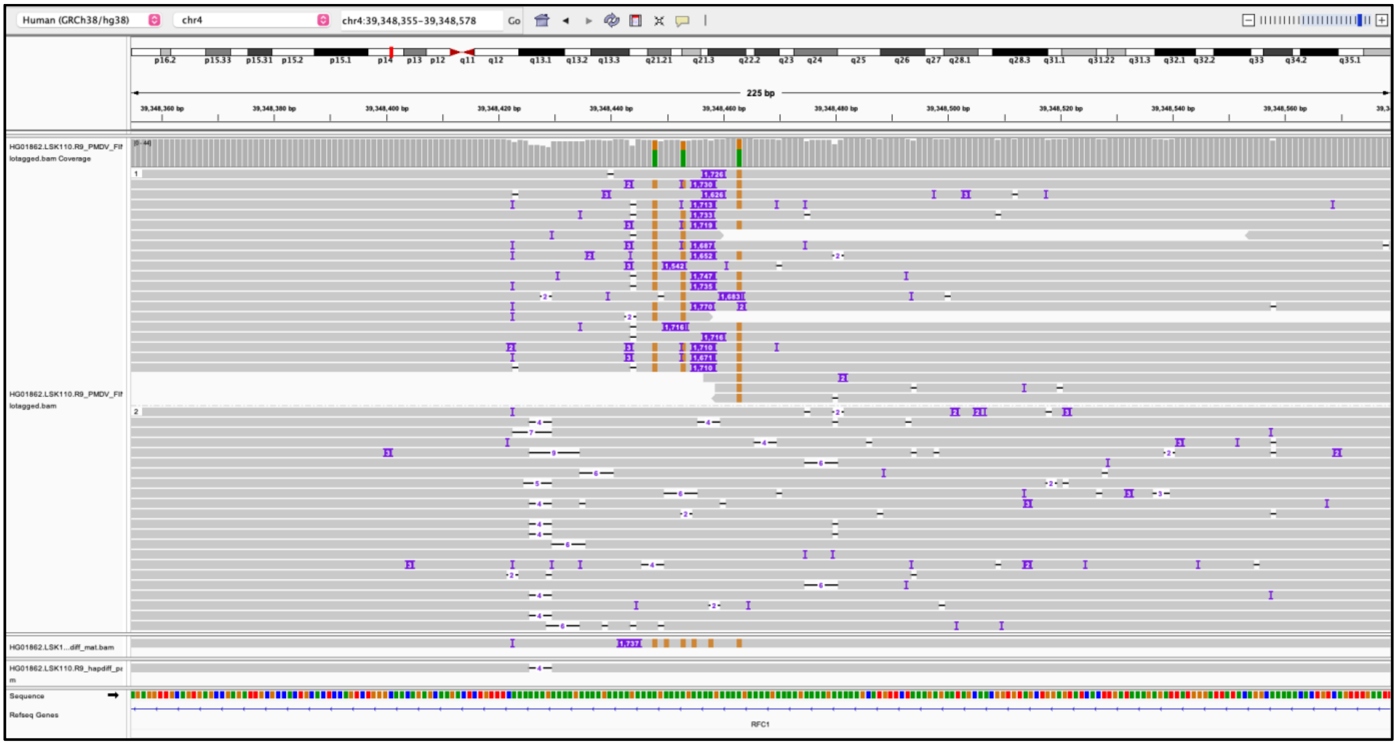

**Figure S18. IGV screenshots of *ATXN10* repeat expansions.**
IGV screenshots for the three samples at the SCA31-associated locus at *ATXN10* with over 250 motifs. The PMDV haplotagged alignment and the hapdiff assemblies from the NAPU pipeline for each sample are shown. The hapdiff assembly files were used for genotyping with vamos.

## HG01122

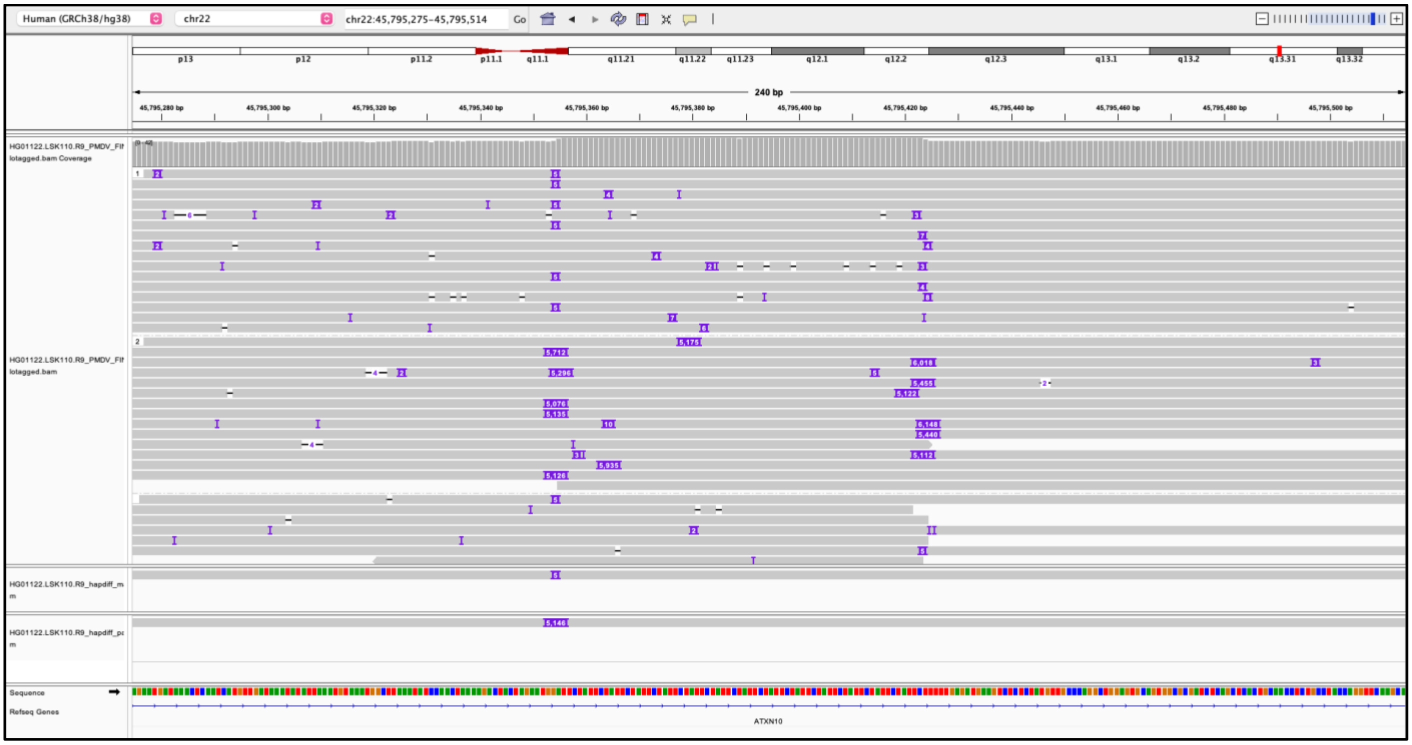

## HG02252

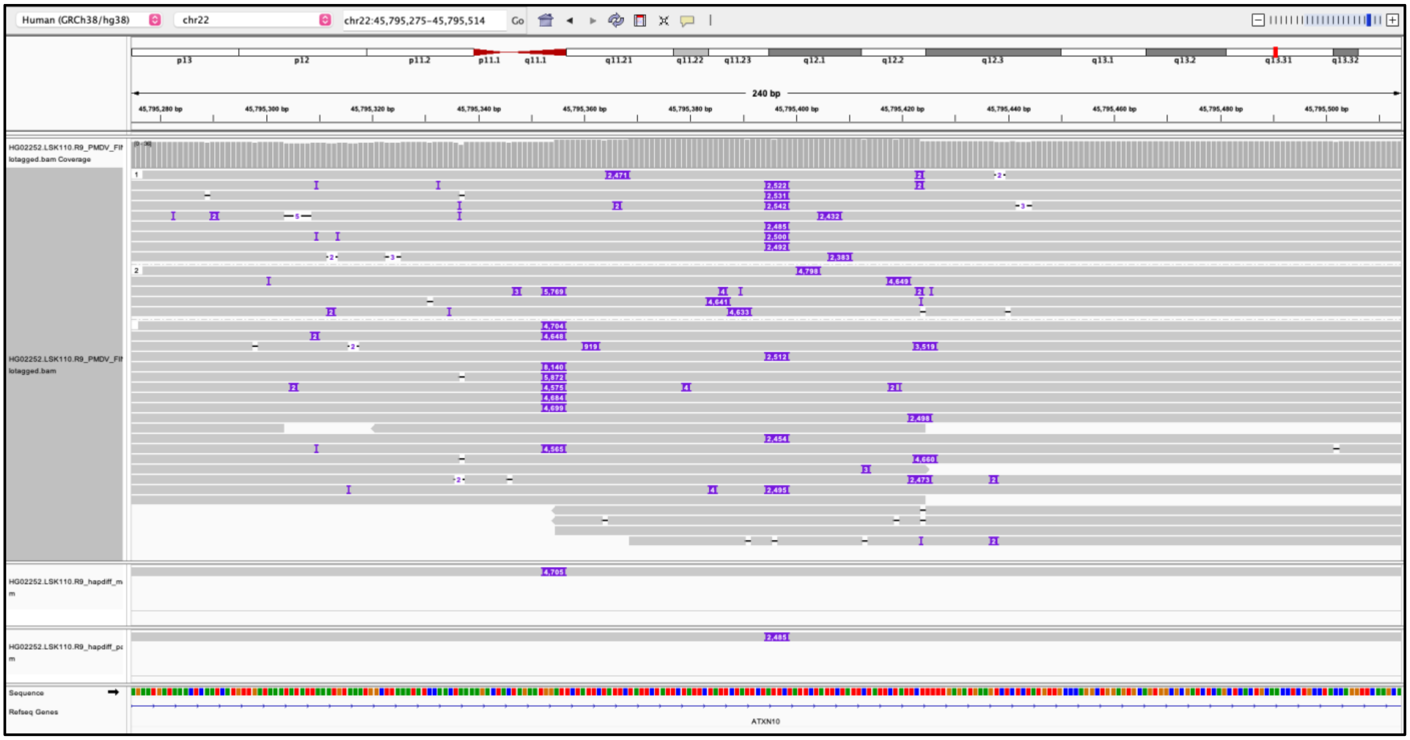

## HG02345

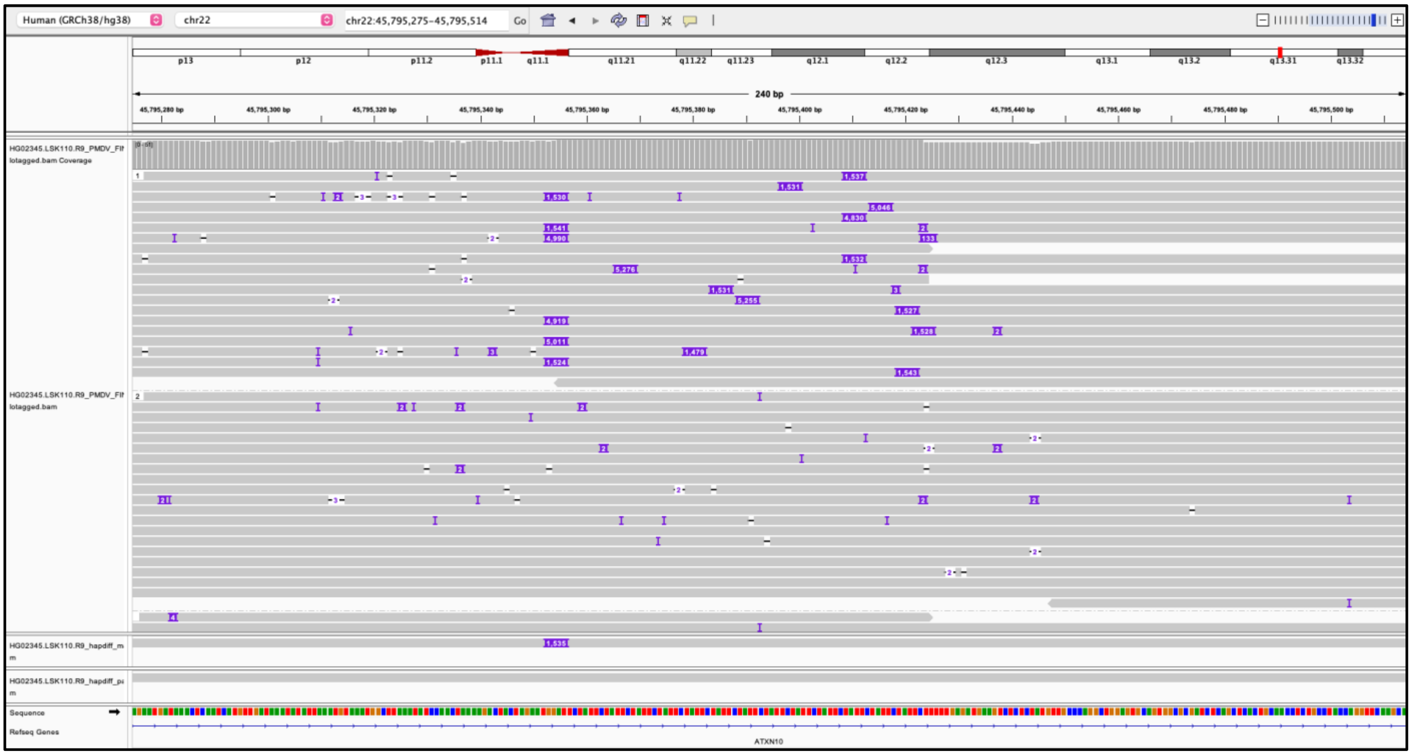
Human (GRCh38/hg38)
chr22
chr22:45,795,275-45,795,514
Go
240 bp
HG02345.LSK110.R9_PMDV_FIt lotagged.bam Coverage
HG02345.LSK110.R9_PMDV_FIt lotagged.bam
Sequence
Refseq Genes
ATXN10

**Figure S19. IGV screenshots of repeat expansion within *FGF14.***
IGV screenshot for the sample at the SCA27B-associated locus at *FGF14* with over 250 motifs. The PMDV haplotagged alignment and the hapdiff assemblies from the NAPU pipeline for each sample are shown. The hapdiff assembly files were used for genotyping with vamos.

**HG01501**

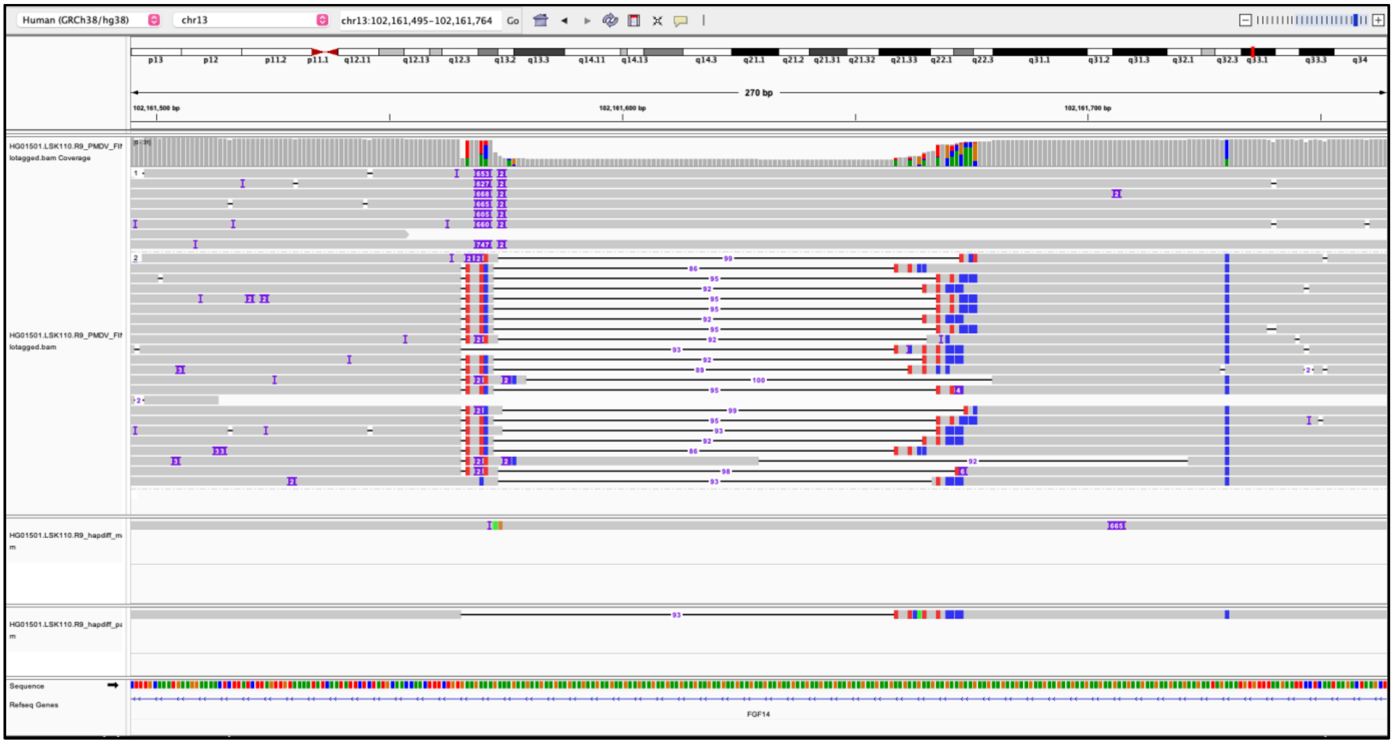

**Figure S20. Principal component analysis using methylation.**
The average methylation frequency for 27,050 CpG islands for 76 samples was used for performing PCA. One 46,XX sample (GM18864) clusters with the 46,XY samples.

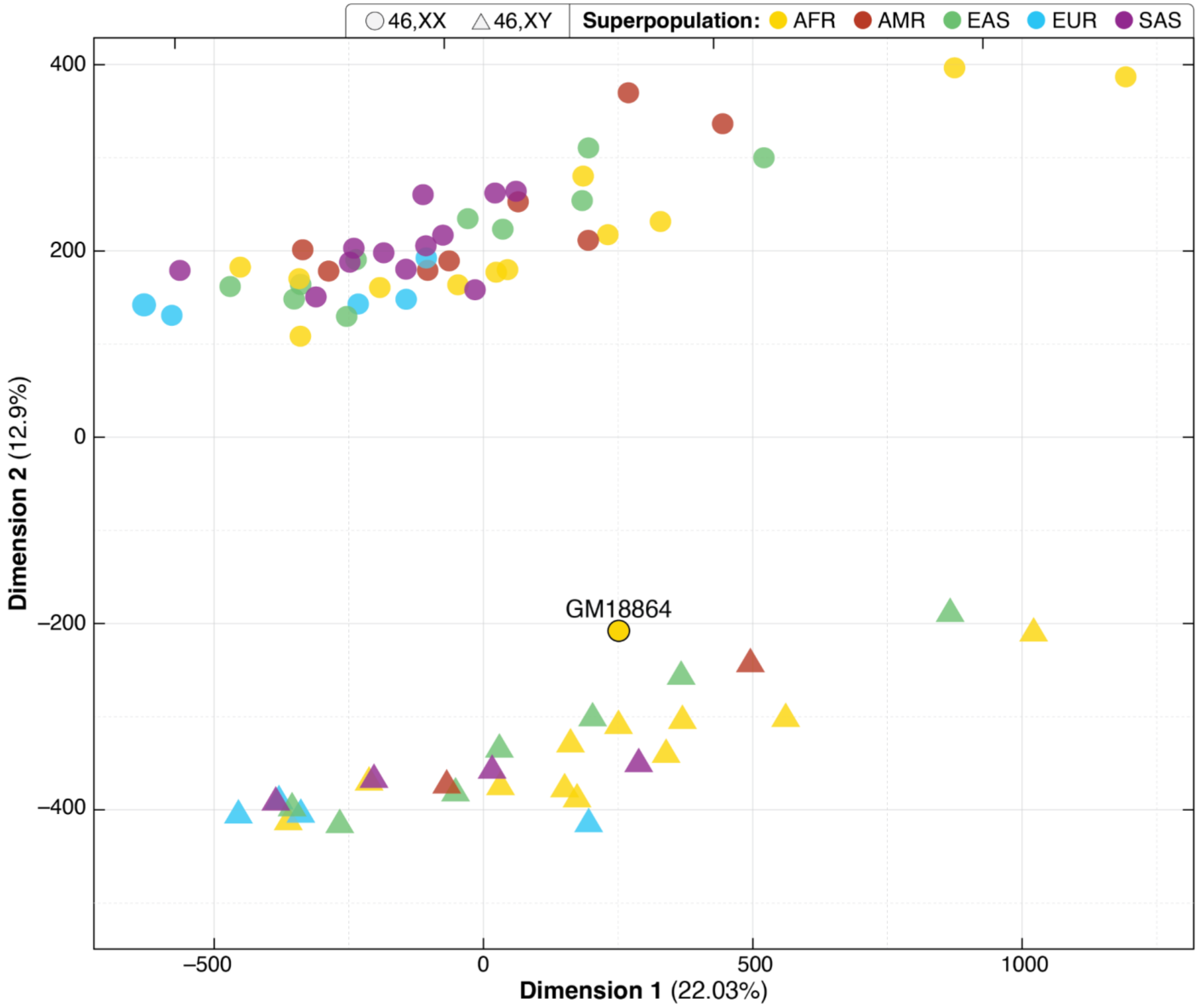

**Figure S21. Evaluating methylation differences at *SNURF-SNRPN*.**
**A**. Gain of methylation was observed in GM19473 (bottom), where both haplotypes are mostly methylated (red). HG03069 (top) is used as a control. **B**. Loss of methylation is seen in HP1 of HG00525 (top). HG04216 (bottom) is used as a control.

A

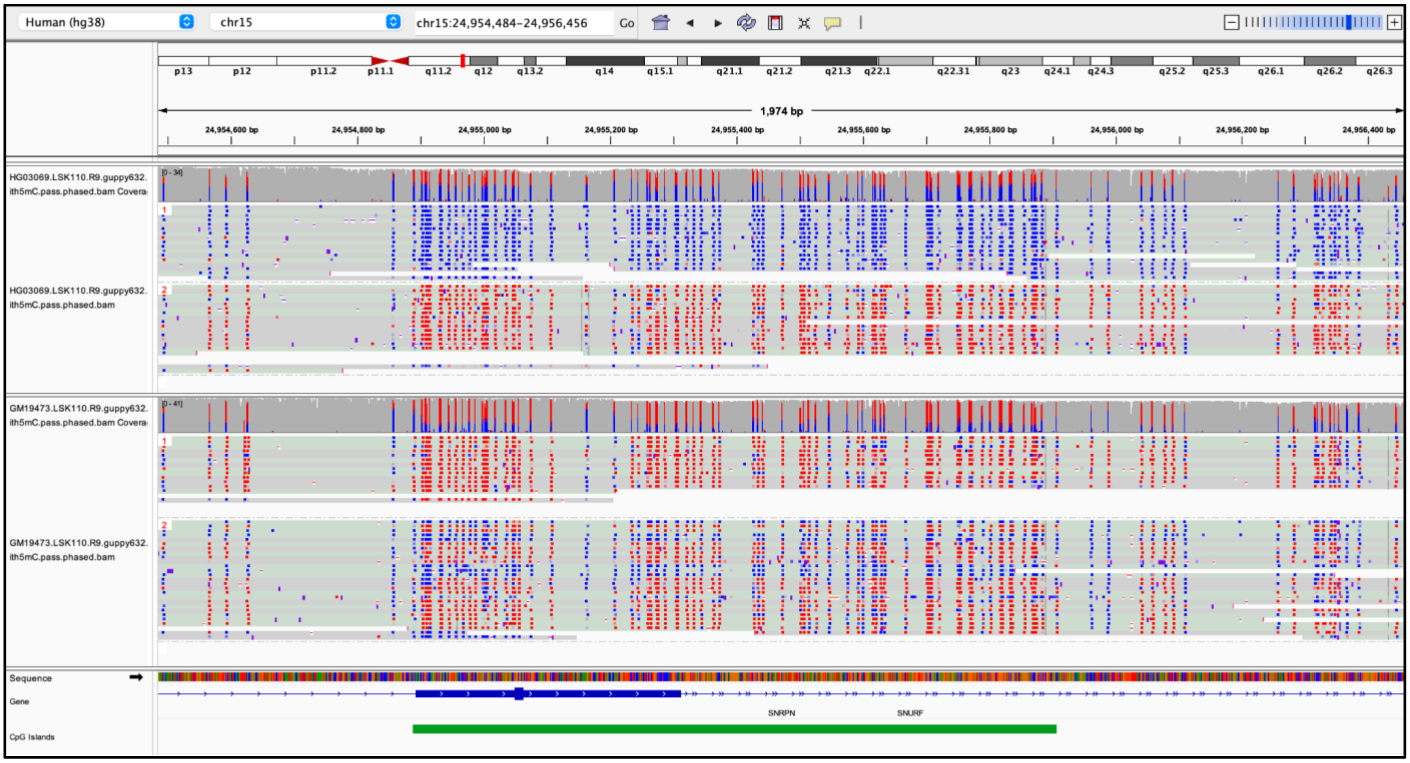

B

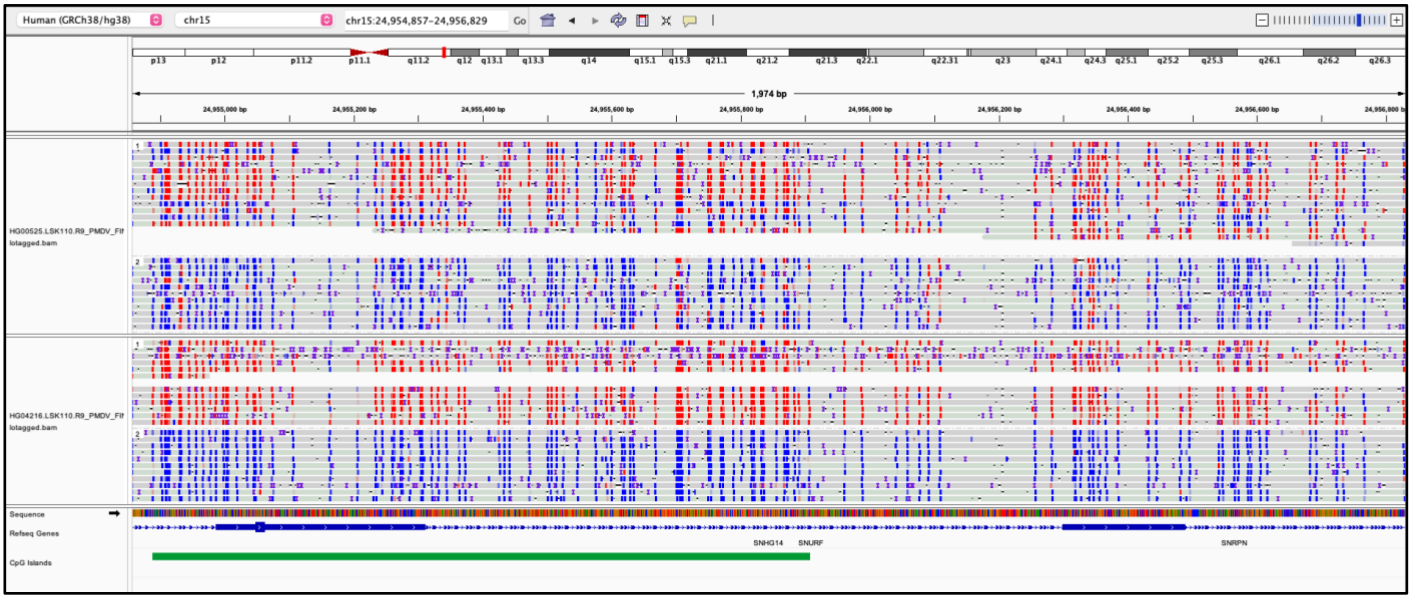

**Figure S22. Output from MeOW showing DMRs within 3 samples that have 10 or more DMRs.**

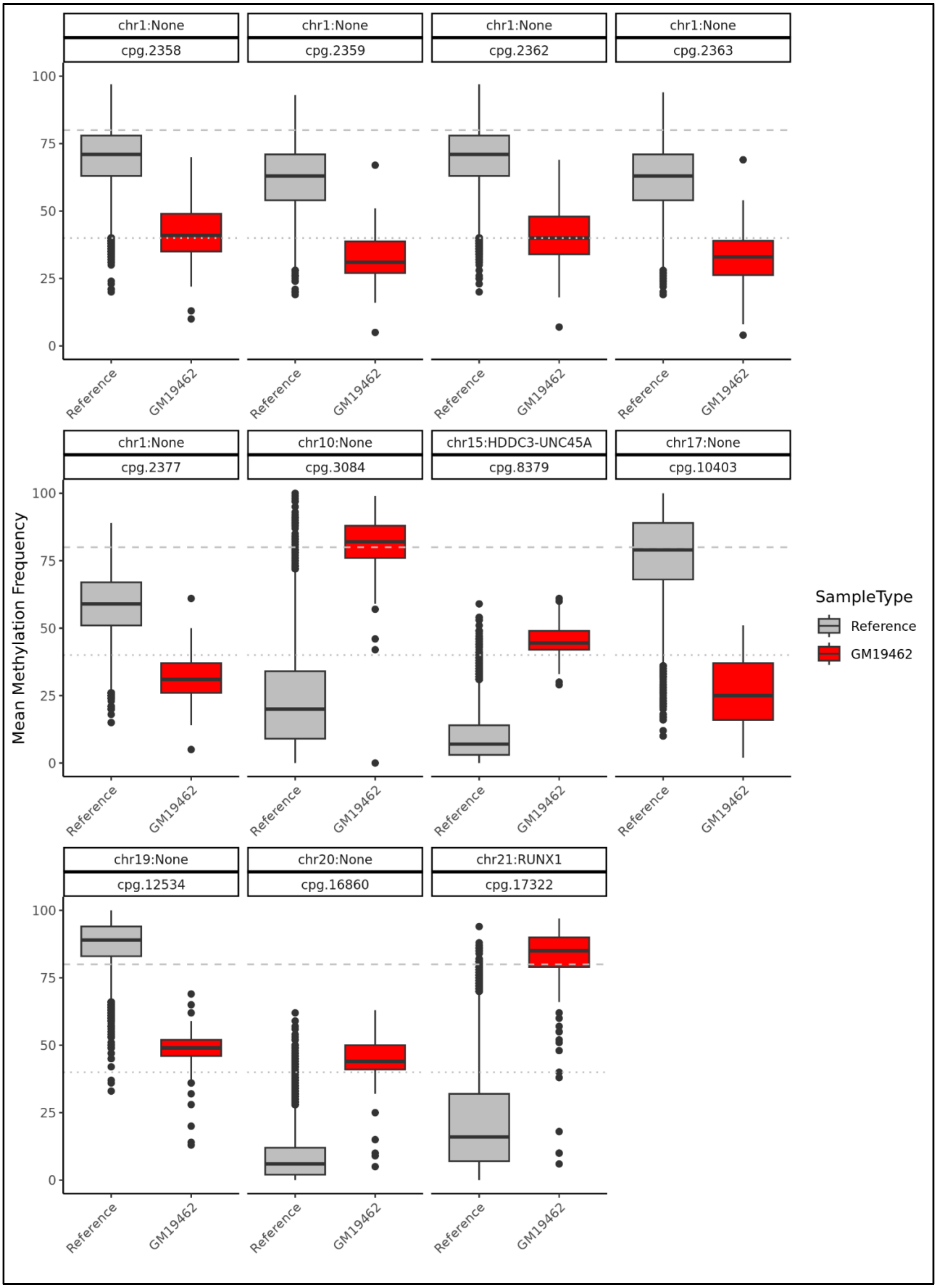

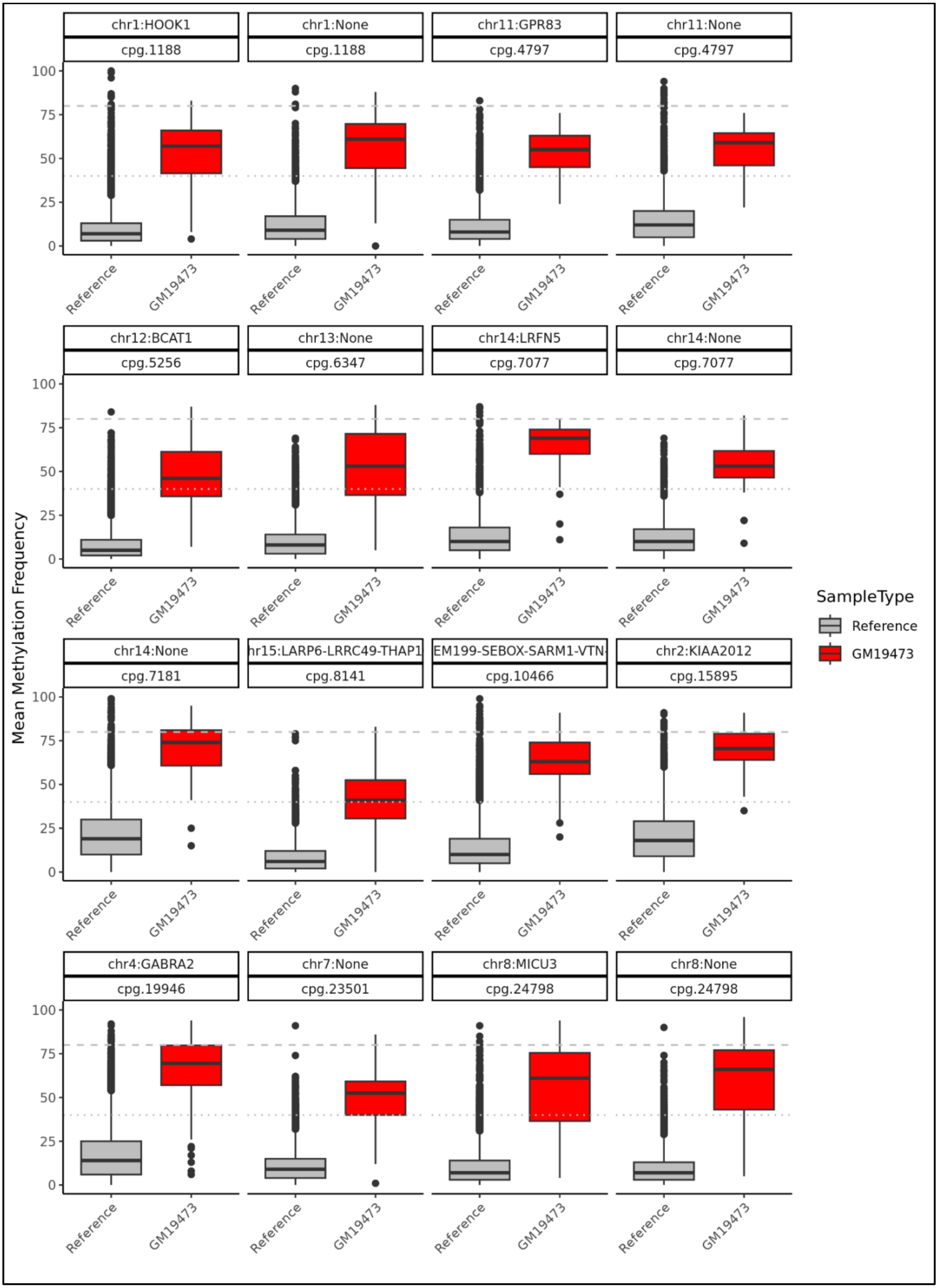
chr1:HOOK1
cpg.1188
chr1:None
cpg.1188
chr11:GPR83
cpg.4797
chr11:None
cpg.4797
chr12:BCAT1
cpg.5256
chr13:None
cpg.6347
chr14:LRFN5
cpg.7077
chr14:None
cpg.7077
chr14:None
cpg.7181
hr15:LARP6-LRRC49-THAP1
cpg.8141
EM199-SEBOX-SARM1-VTN-
cpg.10466
chr2:KIAA2012
cpg.15895
chr4:GABRA2
cpg.19946
chr7:None
cpg.23501
chr8:MICU3
cpg.24798
chr8:None
cpg.24798
Mean Methylation Frequency
0
25
50
75
100
Reference
GM19473
SampleType
Reference
GM19473

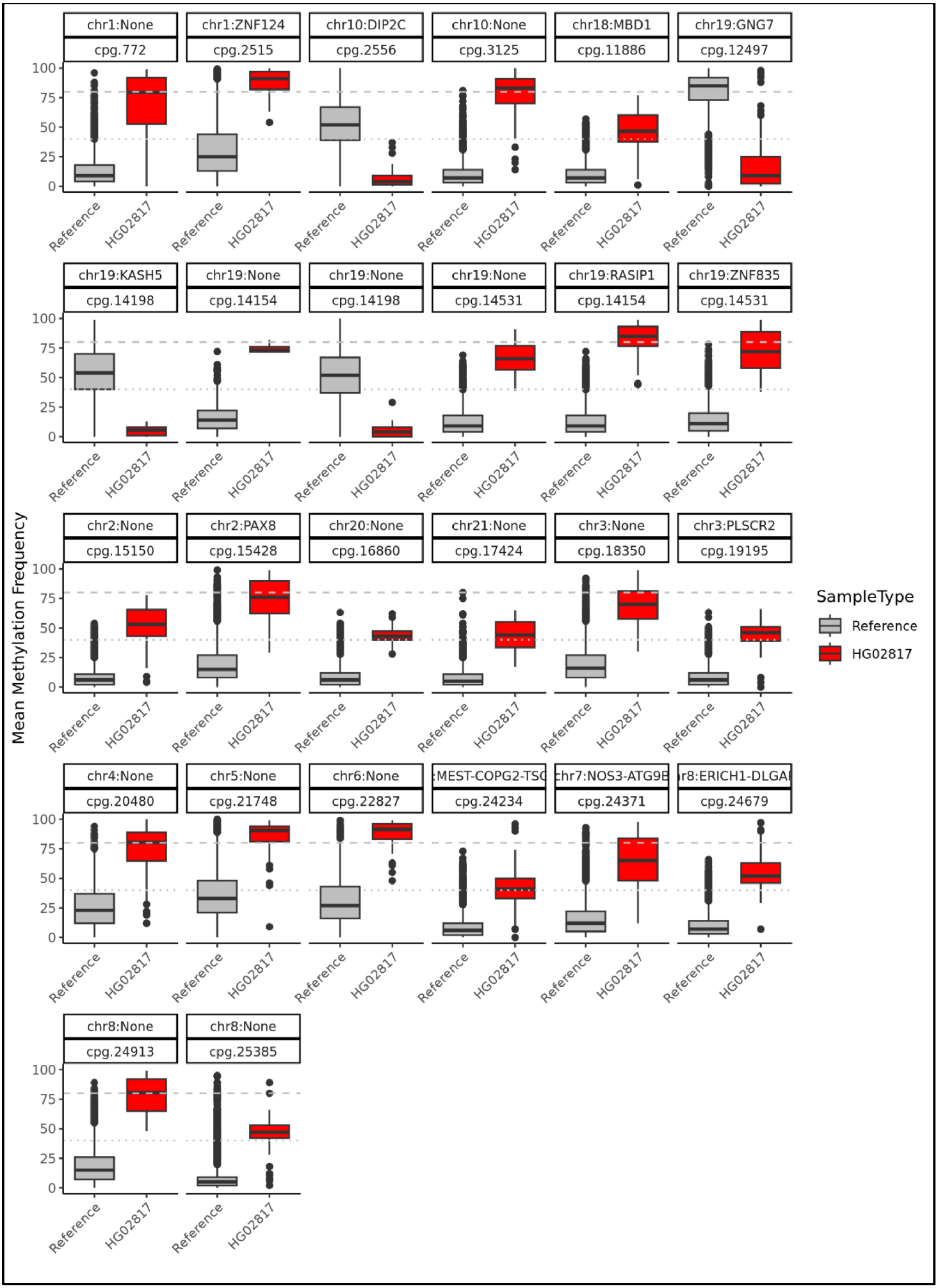
chr1:None cpg.772
chr1:ZNF124 cpg.2515
chr10:DIP2C cpg.2556
chr10:None cpg.3125
chr18:MBD1 cpg.11886
chr19:GNG7 cpg.12497
chr19:KASH5 cpg.14198
chr19:None cpg.14154
chr19:None cpg.14198
chr19:None cpg.14531
chr19:RASIP1 cpg.14154
chr19:ZNF835 cpg.14531
chr2:None cpg.15150
chr2:PAX8 cpg.15428
chr20:None cpg.16860
chr21:None cpg.17424
chr3:None cpg.18350
chr3:PLSCR2 cpg.19195
chr4:None cpg.20480
chr5:None cpg.21748
chr6:None cpg.22827
:MEST-COPG2-TSC cpg.24234
chr7:NOS3-ATG9B cpg.24371
r8:ERICH1-DLGAI cpg.24679
chr8:None cpg.24913
chr8:None cpg.25385
Mean Methylation Frequency
100
75
50
25
0
Reference
HG02817
SampleType
Reference
HG02817

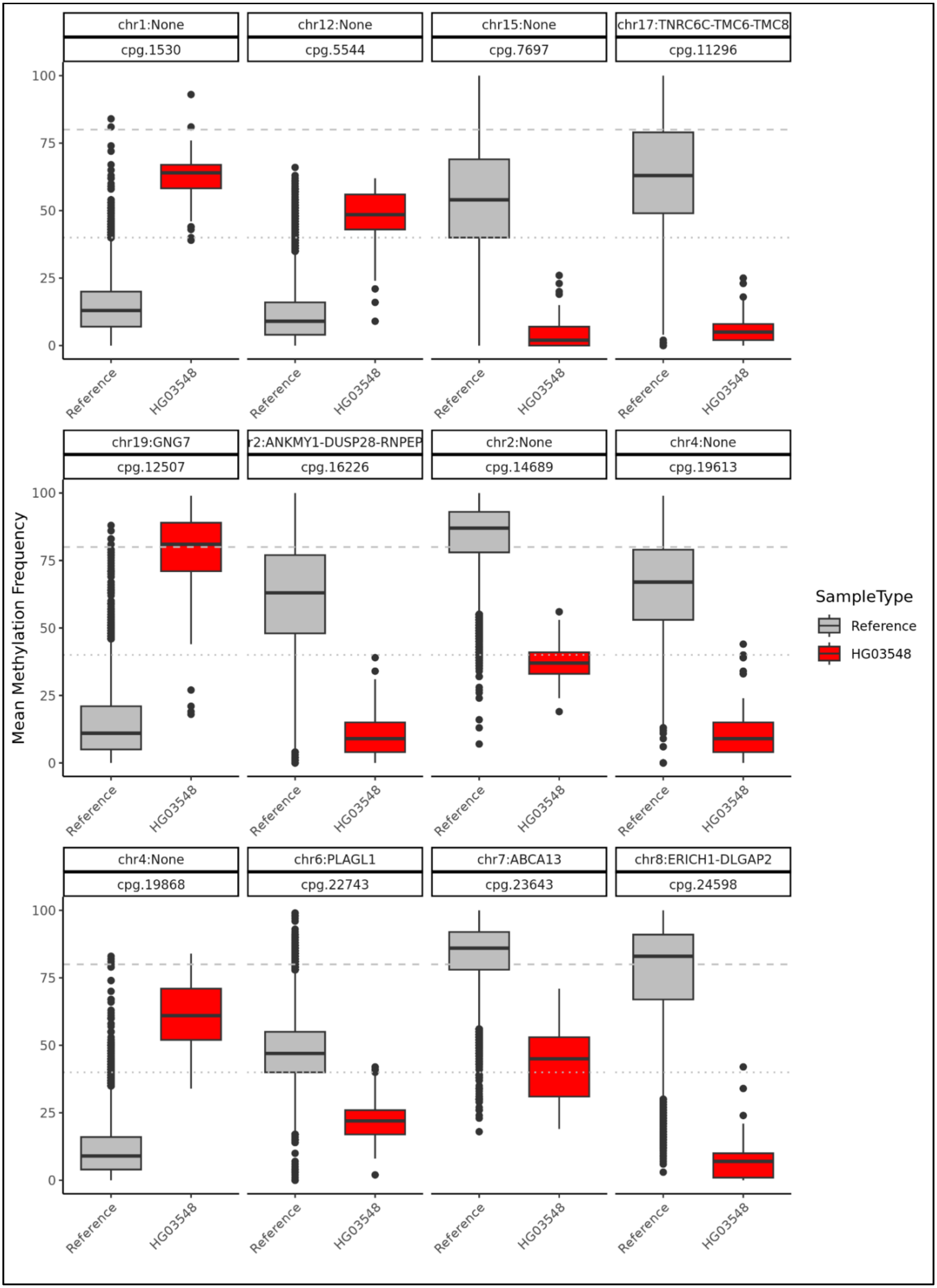
chr1:None
cpg.1530
chr12:None
cpg.5544
chr15:None
cpg.7697
chr17:TNRC6C-TMC6-TMC8
cpg.11296
chr19:GNG7
cpg.12507
r2:ANKMY1-DUSP28-RNPEP
cpg.16226
chr2:None
cpg.14689
chr4:None
cpg.19613
chr4:None
cpg.19868
chr6:PLAGL1
cpg.22743
chr7:ABCA13
cpg.23643
chr8:ERICH1-DLGAP2
cpg.24598
Mean Methylation Frequency
0
25
50
75
100
Reference
HG03548
SampleType
Reference
HG03548

**Figure S23. Unique DMRs are associated with nearby changes in gene expression.**
Expression Z-scores thresholds from RNA-sequencing data for 15 AFGR samples for 85 genes in proximity (10kbp) of DMRs identified with MeOW versus log odds ratios across all sample-gene pairs to identify expression outliers.

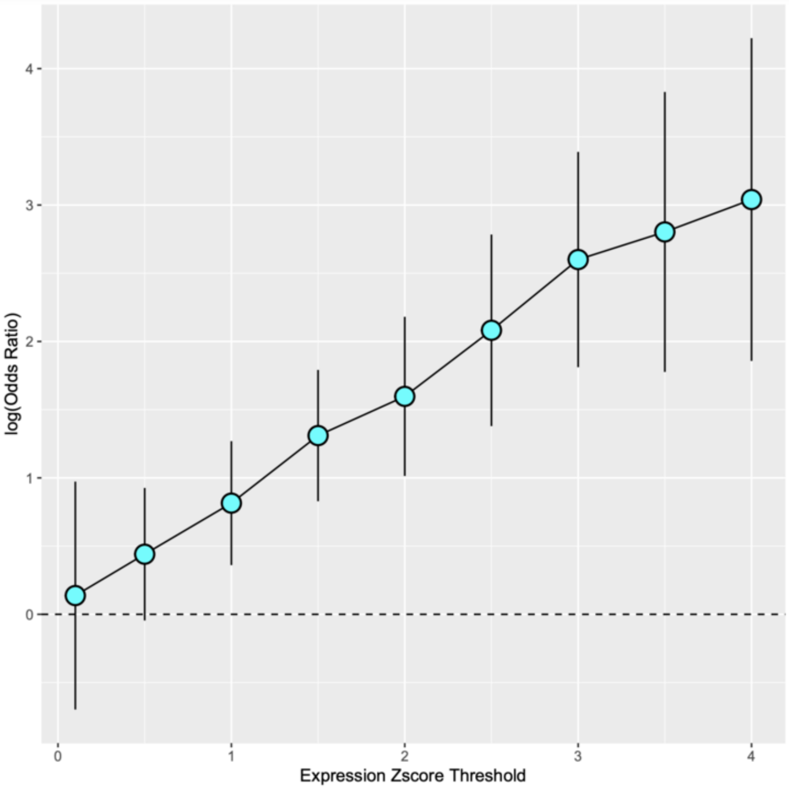
